# Osteoporosis Genetic Risk Prediction Using Bone Mineral Density Polygenic Scores in Japanese: TMM CommCohort Study

**DOI:** 10.64898/2026.01.02.25342989

**Authors:** Yayoi Otsuka-Yamasaki, Yoichi Sutoh, Tsuyoshi Hachiya, Motoki Nakao, Shiori Minabe, Shohei Komaki, Hideki Ohmomo, Makoto Sasaki, Atsushi Shimizu

## Abstract

Osteoporosis and fractures are major health concerns. We developed and validated a polygenic score (PGS) for osteoporosis in a Japanese population using heel quantitative ultrasound–derived T-scores. Genome-wide association study data from 12,371 participants in the Tohoku Medical Megabank Community-Based Cohort identified genome-wide significant loci, including *MBL2, TMEM135,* and *WNT16.* PGS models were constructed and evaluated using independent datasets for model selection (n = 1,419) and validation (n = 8,711). Adding the PGS to age and sex yielded modest improvements in discrimination but supported genetic risk stratification. Compared with the intermediate group, the lowest PGS quintile (bottom 20%, genetically high-risk) had higher odds of osteoporosis (OR = 1.22, 95% confidence interval (CI): 1.07–1.40), whereas the highest PGS quintile (top 20%, genetically low-risk) had lower odds of osteoporosis (OR = 0.85, 95% CI: 0.71–0.98). Prospective follow-up (mean 3.4 years) showed a similar gradient for incident osteoporosis, with higher incidence rate ratios in the high-risk group (1.42, 95% CI: 1.17–1.73) and lower incidence rate ratios in the low-risk group (0.70, 95% CI: 0.54–0.89). Age-stratified analyses revealed no significant age–PGS interaction and no differences in the slope of age-related T-score declines across genetic risk groups. Observed T-scores in young adults (20–44 years) and extrapolation to age 20 suggested lower bone status around peak bone mass among genetically high-risk individuals. These findings indicate that a Japanese-specific PGS can stratify osteoporosis risk and may help identify individuals at elevated genetic risk earlier in adulthood.

## Introduction

Osteoporosis and osteoporotic fractures are significant public health concerns associated with impaired activities of daily living, diminished quality of life, and elevated mortality rates [1]. In older adults, fractures frequently necessitate prolonged care, thereby imposing economic burdens, including elevated expenditures for medical care and long-term care services [2, 3]. The estimated number of patients with osteoporosis in Japan is approximately 12.8 million, with approximately 193,400 hip fractures occurring annually [4].

Bone mineral density (BMD) is widely used as an indicator of bone mass, with heritability (the contribution of genetic factors) estimated at 50–80%, indicating a significant influence of genetic factors [3, 5]. Bone mass follows a characteristic life-course trajectory, with rapid accrual during childhood and puberty, reaching peak levels at 20–30 years of age, remaining relatively stable through midlife, and subsequently declining, particularly after menopause in women [1, 6]. As 40–60% of adult bone mass is formed during puberty, peak bone mass is considered a key determinant of the lifelong risk of developing osteoporosis.^[6]^ Achieving and maintaining a higher peak bone mass during growth has been reported to provide a protective effect against osteoporosis in later life [1, 6, 7].

Advancements in genome-wide association studies (GWASs) have enabled the identification of numerous variants associated with BMD and fracture risk [5, 8]. Polygenic scores (PGS), derived from variants associated with BMD, have been developed to predict the genetic risk of osteoporosis and osteoporotic fractures [5, 9].

Recently, an estimated bone mineral density (eBMD) PGS derived from quantitative ultrasound (QUS) measurements was developed using data from the UK Biobank [5, 9]. The eBMD-based PGS has been evaluated for its predictive accuracy for osteoporosis and fractures in multiple cohorts, as it has shown a negative correlation with osteoporosis and fracture risk [5, 9, 10]. However, studies have consistently revealed that PGS derived from European populations exhibit reduced predictive performance in non-European populations, including East Asians [10–12]. To address this limitation, PGS models tailored to non-Europeans, including East Asians, should be developed [13–17]. Chen et al [17] have tested the multi-ancestry PGS, including Asians in a cohort of Taiwanese people, and showed that it has moderate efficacy, but its performance is not always optimal, and it emphasizes that the development of population-specific PGS is essential in East Asian populations.

Concurrently, it has been shown that incorporating modifiable lifestyle factors into a PGS framework may predict lifetime risk as well as guide targeted prevention strategies [18]. BMD is influenced by both basic and unchangeable environmental factors, such as sex, age, and race, as well as modifiable factors such as calcium intake, smoking, alcohol consumption, physical activity, and sun exposure [6,19]. Even individuals with a high genetic risk for osteoporotic fractures can reduce their risk by adhering to healthy lifestyle habits in European populations [19].

In this study, we developed a PGS for QUS-derived T-score in a Japanese population using data from the Tohoku Medical Megabank (TMM), the largest genome cohort in Japan [20–22], and evaluated its ability to predict osteoporosis risk. We assessed genetic and environmental factors at baseline and followed participants over time to capture incident osteoporosis using a longitudinal design. We stratified participants based on PGS, examined its joint effects with non-modifiable and modifiable risk factors, and evaluated its potential as a tool to inform osteoporosis prevention strategies in Japan.

## Materials and Methods

### Study population

TMM Project Community-Based Cohort (TMM CommCohort) has been described in previous studies [20–22]. Participants who visited the evaluation centers in Miyagi and Iwate Prefectures, were grouped into two primary cohorts: TMM18K and TMM9K, respectively. The baseline survey was conducted from May 2013 to March 2016, and the secondary survey, conducted 4 years after the baseline survey, was conducted from May 2017 to March 2020 as part of the ongoing follow-up.

The two cohorts were further divided into four groups based on genotyping with the Axiom Japonica Array v2 (JPAv2) [23, 24] or NEO (JPANEO) [25] (TMM18K-JPAv2, TMM18K-JPANEO, TMM9K-JPAv2, and TMM9K-JPANEO) (Figure 1 and Supplementary Table S1). The TMM18K-JPAv2 cohort (n = 12,371) was used to perform QUS-derived T-score GWAS, from which the variant weights for the PGS model were derived. The TMM9K-JPANEO cohort (n = 1,419) was used to identify the best model for predicting osteoporosis. The TMM9K-JPAv2 (n = 7,621) and TMM18K-JPANEO (n = 1,090) cohorts were used for subsequent analyses. All four sample subgroups were independent from one another.

**Figure 1.**
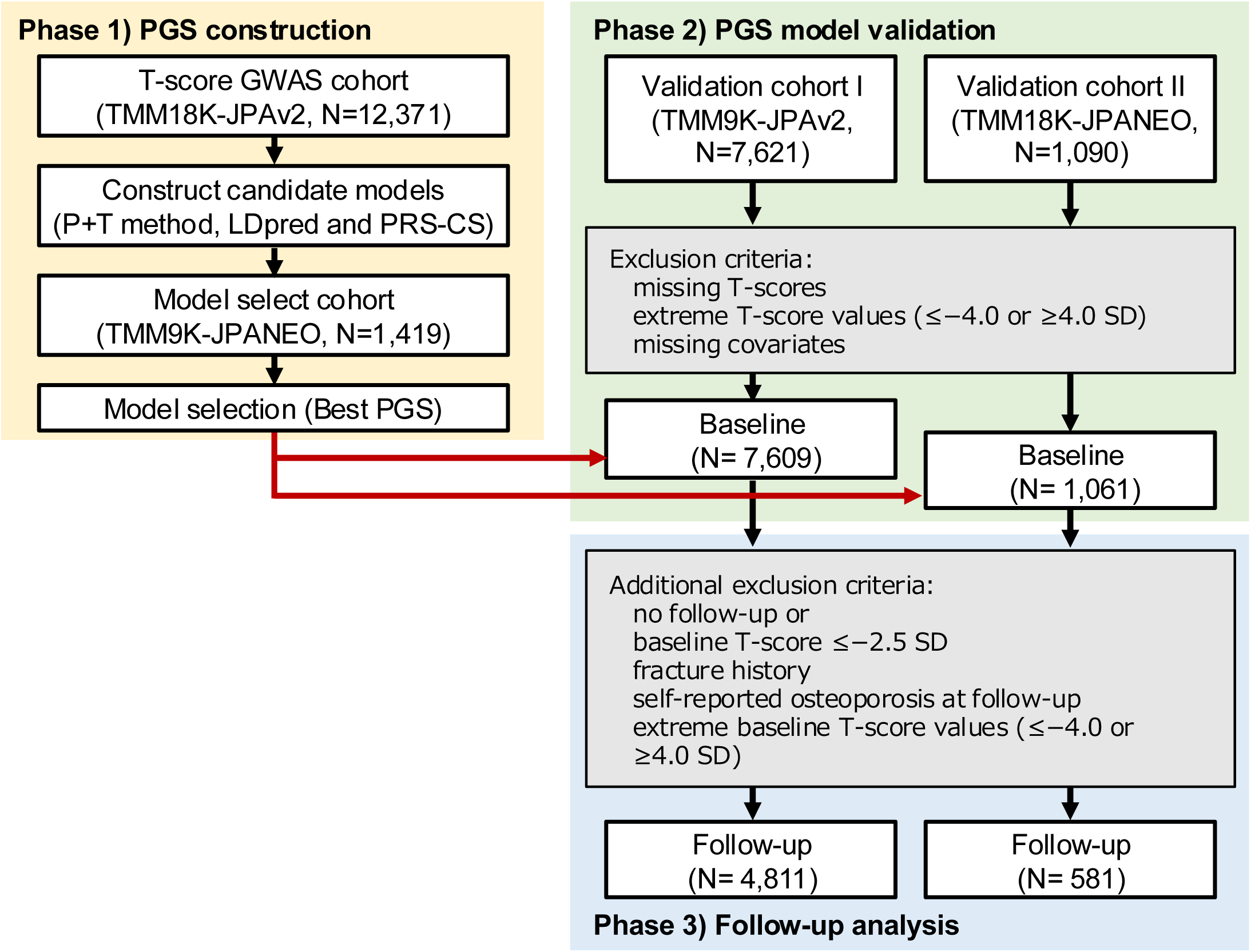
Study design. Two TMM cohorts (9K and 18K) [20–22] were split using genotyping array, Axiom Japonica Array v2 (JPAv2) [23, 24] and NEO (JPANEO) [25], into four independent subgroups (TMM18KlZIJPAv2, TMM18KlZIJPANEO, TMM9KlZIJPAv2, and TMM9KlZIJPANEO). QUS-derived T-score GWAS was conducted in TMM18KlZIJPAv2 to derive variant weights; model selection was performed in TMM9KlZIJPANEO (Phase 1). The final PGS was evaluated in two independent validation cohorts: I (TMM9KlZIJPAv2) and II (TMM18KlZIJPANEO). Each cohort underwent (Phase 2) crosslZIsectional analysis at baseline (excluding participants with missing TlZIscores, extreme baseline T-scores (outside the range −4.0 to +4.0 SD), or missing covariates) and (Phase 3) prospective analysis using followlZIup data with additional exclusions (no followlZIup, baseline TlZIscore ≤ –2.5 SD, fracture history, selflZIreported osteoporosis at followlZIup, or extreme baseline T-scores (outside the range −4.0 to +4.0 SD)). Final sample sizes were 7,609 and 1,061 for crosslZIsectional, and 4,811 and 581 for prospective analyses in validation cohorts I and II, respectively. All four subgroups were independent. Abbreviations: GWAS, genome-wide association study; JPANEO, Japonica array NEO; JPAv2, Japonica array version 2; PGS, polygenic score; PRS-CS, polygenic risk score—continuous shrinkage method; P+T, pruning and thresholding method; SD, standard deviation; TMM, Tohoku Medical Megabank.

### Phenotype definition and measurement

Calcaneal QUS measurements were performed using a Benus device (NIHON KOHDEN, Tokyo, Japan). The device-derived calcaneal T-score was calculated using the manufacturer’s young-adult reference (ages 20–40 years). We classified participants as having osteoporosis when the device-derived QUS T-score was ≤ −2.5. Fracture cases were identified based on the self-reported history of fractures in the lumbar spine, femur, wrist, or humerus as documented by the participants in the questionnaire. Other anthropometric and lifestyle factors, including alcohol intake and smoking history, were obtained from self-reported questionnaires administered at baseline.

Participants who had smoked at least 100 cigarettes in their lifetime were defined as having a “smoking” status. “High alcohol intake” was defined as the consumption of ≥ 30 g of alcohol per day. “Physical inactivity” was defined as engaging in leisure-time physical activity less than three times weekly. “Obesity” was defined as a body mass index (BMI) of ≥ 25 kg/m^2^, calculated from height and weight measured at baseline, in accordance with the local definition in Japan [26].

### Study participants and validation cohort definitions

To evaluate the predictive performance of PGS, we used two independent validation cohorts: TMM9K-JPAv2 (validation cohort I) and TMM18K-JPANEO (validation cohort II). We conducted cross-sectional and prospective analyses based on baseline and follow-up survey data, respectively.

For the cross-sectional analysis, we excluded participants with (1) missing T-score values at baseline, (2) extreme baseline T-scores (outside the range −4.0 to +4.0 SD), or (3) missing essential covariates. The final sample included 7,609 participants in validation cohort I and 1,061 participants in validation cohort II (Supplementary Table S1).

For the prospective analysis, additional exclusion criteria were applied: (1) no participation in the follow-up survey; (2) baseline T-score ≤−2.5 SD; (3) history of fracture (lumbar spine, femur, wrist, or humerus); (4) self-reported history of osteoporosis at follow-up (because of unclear timing); and (5) extreme baseline T-scores (outside the range −4.0 to +4.0 SD). After these exclusions, the prospective subcohorts included 4,811 participants in validation cohort I and 581 participants in validation cohort II (Supplementary Table S1).

### Genotyping and imputation

It was described in previous studies that the genotyping was performed using either JPAv2 [23, 24] or NEO (JPANEO) [25]. Participants with a call rate of < 0.95 and sex discrepancies between genotypic and cohort data were excluded. Genotypes that failed to meet the quality control criteria (call rate of < 0.99, Hardy–Weinberg equilibrium exact test *P* of < 1 × 10^−5^, and minor allele frequency of < 0.01) were also omitted. Pre-phasing and imputation were performed using SHAPEIT2 (r837) and IMPUTE4 (r300.3) with 3.5KJPNv2 haplotype reference panels [23, 28, 29]. Variant positions in the genome were based on the GRCh37 reference genome.

### Genome-wide association analysis and annotations

To generate the PGS model, single nucleotide polymorphism (SNP) weights were estimated using a publicly available GWAS summary of TMM18K-JPAv2 from jMorp (ID: TGA000015; https://jmorp.megabank.tohoku.ac.jp/). Additional methodological details are available on the official website. In brief, GWAS was conducted with BOLT-LMM v2.3.2 [27]. Rank-based inverse normalization was applied to reduce the skewness and kurtosis in the T-score distribution. Age, sex, and the first 11 principal components were included as fixed effect covariates.

Annotation was performed using an in-house pipeline tailored to the Japanese population. The ToMMo 3.5KJPNv2 reference panel [28] was used to assign allele frequency information and dbSNP identifiers (rsIDs) to variants in the GWAS summary statistics. In addition, the gnomAD v2.1 dataset [29] was incorporated to ensure comprehensive rsID annotation and facilitate cross-population comparisons. Regional association plots were generated using LocusZoom (version 0.14.0) [30]. Linkage disequilibrium patterns and proxy variants were examined using LDproxy (LDlink, National Cancer Institute) [31] with the 1LKG EAS reference panel.

### PGS construction and selection

Candidate models were constructed as described previously using three methods: LD pruning and thresholding (P+T), LDpred and polygenic risk score–continuous shrinkage (PRS-CS) [26, 32–34]. For the P+T method, 24 models were generated using different LD-pruning conditions using one of six parameters for p values (1, 0.5, 5 × 10^−2^, 5 × 10^−4^, 5 × 10^−6^, and 5 × 10^−8^) and one of four parameters of *R^2^* values (0.2, 0.4, 0.6, and 0.8). For LDpred, seven models were generated for each Spearman’s rank correlation (ρ) value (1, 0.3, 0.1, 0.03, 0.01, 0.003, and 0.001). For PRS-CS, six models were generated for each φ value (1 × 10^−1^, 1 × 10^−2^, 1 × 10^−4^, 1 × 10^−6^, 1 × 10^−8^, and “auto”). Individual PGSs were calculated using PLINK 2.0 for each model and normalized to Z-scores.

In the selection step, TMM9K-JPANEO cohort was used as the dataset for model selection. The model with the highest Spearman rank correlation coefficient (ρ) between the age- and sex-adjusted T-scores and PGS Z-scores was selected.

Statistical analyses were performed using R (version 4.2.1). The receiver operating characteristic (ROC) curve and area under the curve (AUC) for the PGS model was estimated using the R package “pROC” (version 1.18.0). The AUCs between the models were compared using DeLong’s test.

### PGS-based risk assessment and stratified analysis

The normalized PGS was divided into quintiles. Cross-sectional associations at baseline were tested using linear regression for continuous outcomes and logistic regression for binary outcomes, adjusting for sex and age. In addition, we conducted stratified cross-sectional analyses by baseline age (< 65/≥ 65 years), sex, and major osteoporosis-related risk factors (smoking, alcohol intake, physical activity, and obesity). For these stratified analyses, participants were classified into three genetic risk groups: High (Q1), Intermediate (Q2–Q4), and Low (Q5), using the Intermediate group as the reference category in all comparisons.

For prospective analyses, participants without osteoporosis at baseline were followed up for an average of 3.4 years. Because the exact event dates were unavailable, person-time was approximated using the midpoint method: incident cases contributed half of the follow-up duration, whereas non-cases contributed the full duration. Incidence rates per 1,000 person-years and cumulative incidence were estimated by genetic risk group with exact 95% confidence intervals (CI).

Results from the two validation cohorts were pooled using the R package “meta” (version 6.2-1). Fixed-effect models were used for the primary meta-analyses. Between-cohort heterogeneity was assessed using Cochran’s Q test and the I^2^ statistic.

Random-effects models (DerSimonian–Laird method) were also fitted, and were examined as sensitivity analyses to confirm the robustness of the results in the presence of substantial heterogeneity (Q test *P* < 0.05 or I^2^ > 50%).

### Ethical considerations

This research was conducted in accordance with the ethical principles outlined in the Declaration of Helsinki. All participants provided written informed consent prior to their enrollment. The study protocol was approved by the Institutional Review Board of Iwate Medical University (approval ID: HG H25-2). Analyses using individual-level genomic and phenotypic data were performed on a secure, standalone computational platform operated by the Tohoku Medical Megabank Organization (ToMMo) at Tohoku University (ToMMo Supercomputer System URL; https://sc.megabank.tohoku.ac.jp/).

### Data Availability

The QUS-derived T-score GWAS summary has been published in jMorp (https://jmorp.megabank.tohoku.ac.jp/gwas-studies/TGA000015). 3.5KJPNv2 is also available on the jMorp website with a web interface. Owing to ethical considerations, including the protection of participant privacy and prevention of unintended identification, individual-level data from the TMM CommCohort Study are not publicly available. Access to these cohorts may be granted upon reasonable request and is subject to approval by the Ethics Committee of Iwate Medical University and the Materials and Information Distribution Review Committee of the TMM Project. Researchers interested in accessing these datasets should contact the corresponding author to initiate the request.

## Results

### Heritable factors for QUS-derived T-score

GWAS was conducted to identify variants associated with the T-score (Supplementary Figure S1A). Variants in three independent loci were above the threshold for the genome-wide significance (*P*ϑ*<*ϑ5L×L10^−8^) (Supplementary Table S2). Notably, 792 variants were noted above the suggestive line (*P*L< 1L×L10^−5^) (Supplementary Table S3).

The three loci containing significant variants included genes that had been reported in previous GWASs using QUS-derived T-scores as traits, such as Mannose Binding Lectin 2 (*MBL2*), Transmembrane Protein 135 (*TMEM135*), and *Wnt* Family Member 16 (*WNT16*) [3,5,8] (Supplementary Table S2). The UK Biobank GWAS summary statistics and GWAS catalog study GCST006433 [5] were used for comparison.

As shown in Supplementary Table S2, at the *MBL2* locus, the lead variant in the Japanese GWAS was rs71032654/rs200903639 (del(TA)_9_CA > dup(TA)_9_CA), a structural indel. Notably, neither rs71032654 nor rs7908619 was present in the UK Biobank summary statistics (Supplementary Table S2). LocusZoom plots showed recombination hotspots exceeding 30 cM/Mb on either side of rs71032654 (Supplementary Figure S1C). Based on the LDproxy results, rs71032654/rs200903639 showed only low LD with surrounding variants (maximum *R*^2^ = 0.0687), indicating that it may represent a distinct signal from previously known tagged variants.

Variants in rs2451053 (C>T) in *TMEM135* and rs10254825 (A>G) in *WNT16* were present in both the TMM and UK Biobank cohorts, indicating their similar biological effects on BMD across populations.

### Determination of the optimal PGS model for T-score measured with QUS

Based on the above GWAS summary, we constructed 37 candidate models to calculate the PGS using three distinct methods (Figure 2A and Supplementary Table S4). We determined that the PGS model established by PRS-CS with φ set to “auto” which was composed of 1,012,314 variants, showed the strongest correlation with T-score levels after adjusting for age and sex (Spearman’s ρ = 0.147, *P* = 2.94 × 10^-8^). Therefore, this model was selected and used for subsequent validation analyses as the best model.

**Figure 2.**
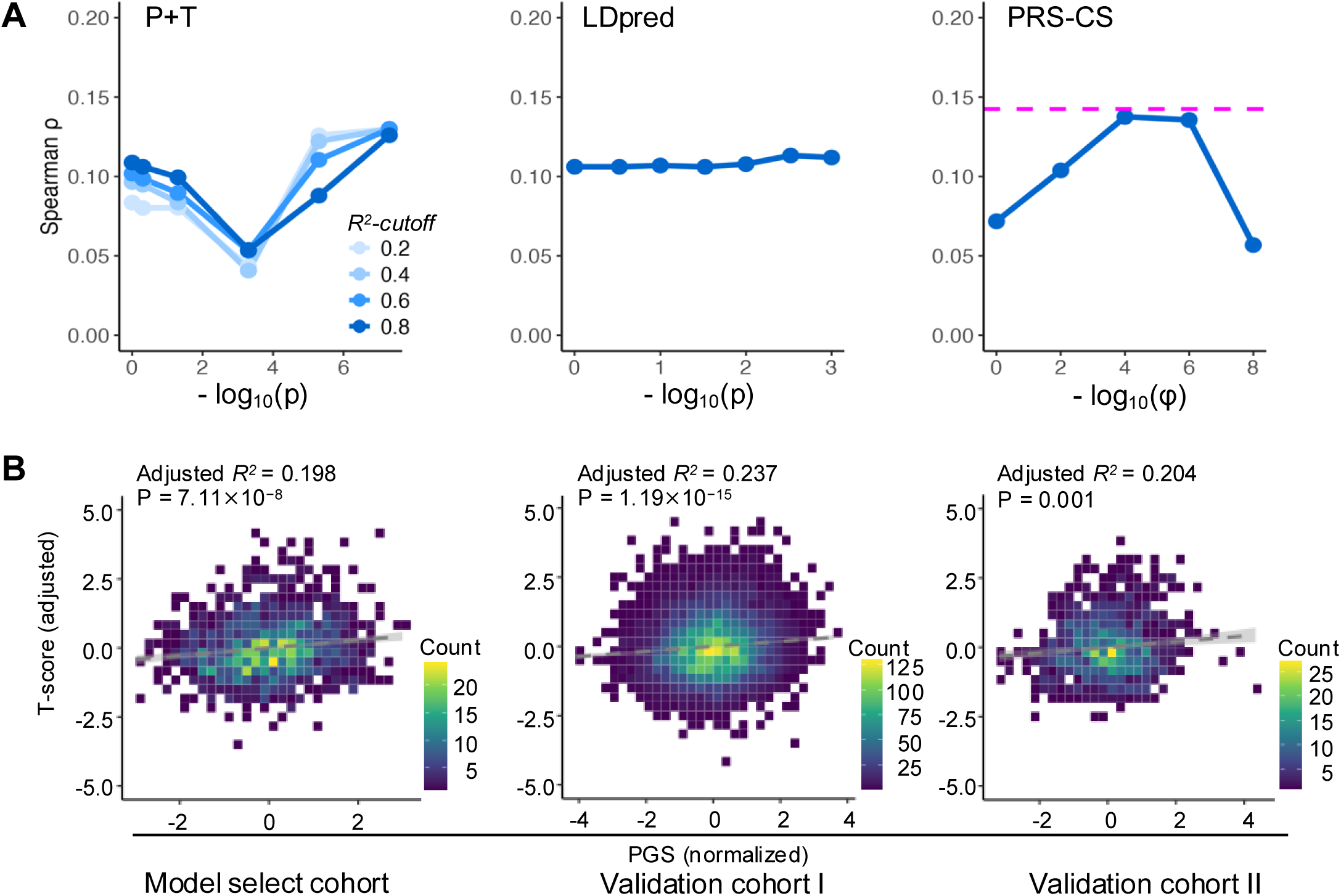
Model selection of T-score PGSs. (A) Discriminative performance (ρ) of 37 candidate models generated using P+T, LDpred, and PRS-CS methods. PRS-CS (φ = “auto”) showed the highest ρ (ρ = 0.147, *P* = 2.94 × 10^-8^). (B) Correlation of PGSs between the model selection cohort (TMM9K-JPANEO) and validation cohorts I and II (TMM9K-JPAv2 and TMM18K-JPANEO). Scatterplots show the relationship between normalized PGSs (x-axis) and age- and sex-adjusted T-scores (y-axis). Dashed lines represent fitted linear regression lines with 95% CI. Color gradients indicate sample density. *P* values are from linear regression analyses. Abbreviations: PGS, polygenic score; PRS-CS, polygenic risk score—continuous shrinkage method; P+T, pruning and thresholding method.

Linear regression analysis of baseline T-scores against normalized PGS, adjusted for sex and age, revealed a significant positive association in validation cohorts I (Estimate = 0.090, *P =* 1.19 × 10^−15^, Adjusted *R^2^* = 0.237), and II (Estimate = 0.098, *P* = 0.001, Adjusted *R^2^* = 0.204) (Figure 2B and Supplementary Table S5).

### Predictive performance and association of T-score PGS with osteoporosis

To investigate the predictive utility of PGS for osteoporosis, we evaluated the performance of the model after including PGS in addition to sex and age. In validation cohort I, there were 1,286 cases of osteoporosis out of 7,609, and in validation cohort II, there were 233 cases of osteoporosis in 1,061 patients (Supplementary Table S6). In validation cohorts I and II, the AUC values of the baseline model, including sex and age were 0.726 (95% CI: 0.711–0.741) and 0.659 (95% CI: 0.622–0.696), respectively (Supplementary Figure S2A and Supplementary Table S7). Adding PGS to the model resulted in slight improvements, with AUC values of 0.728 (95% CI: 0.713–0.743) and 0.664 (95% CI: 0.627–0.701) in validation cohorts I and II, respectively (Supplementary Figure S2A and Table S6).

We assessed the association between the T-score PGS and osteoporosis using multivariable models in the two validation cohorts (Supplementary Figure S2B and Supplementary Table S8). A fixed-effect model indicated minimal intercohort heterogeneity for the PGS effect (I^2^ = 0%, Cochran’s Q = 0.02, P for heterogeneity = 0.878). Accordingly, we report the fixed-effect pooled estimate. Each 1.0-unit increase in the normalized PGS was associated with 12% lower odds of osteoporosis (OR = 0.88, 95% CI: 0.83–0.94, *P* = 2.77 × 10^-5^), indicating an inverse association between the T-score PGS and osteoporosis risk. In addition, the odds of osteoporosis increased by 7% per year of age (OR = 1.07, 95% CI: 1.07–1.08, *P* = 3.03 × 10^-115^), and was 1.84-fold higher in women than in men (OR = 1.84, 95% CI: 1.57–2.17, *P* = 2.02 × 10^-13^).

### Osteoporosis risk prediction using T-score PGS

For further analysis, individuals in the validation cohorts were stratified into quintiles using the PGS. In the fixed-effect meta-analysis of the two validation cohorts, individuals in the lowest PGS quintile (Q1), which also had the lowest mean T-score (Supplementary Table S9), had a significantly increased risk of developing osteoporosis compared to the intermediate PGS quintile (Q3) (Table 1). Moreover, the OR for Q2 was significantly higher than that for Q3 (Table 1). In contrast, the highest PGS quintile (Q5), which had the highest mean T-score (Supplementary Table S9), was associated with a significantly lower risk of osteoporosis than Q3 (Table 1).

**Table 1.**
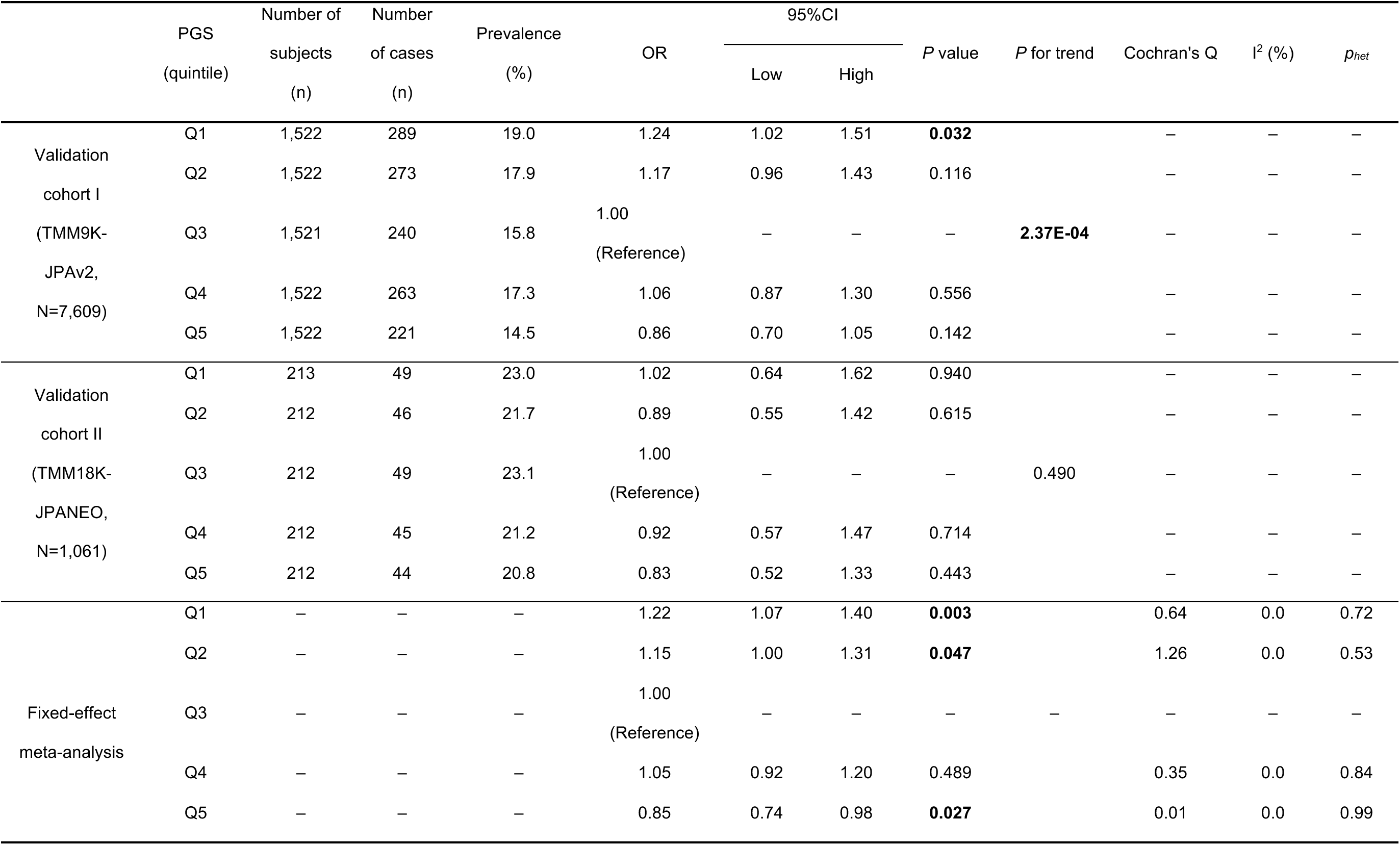
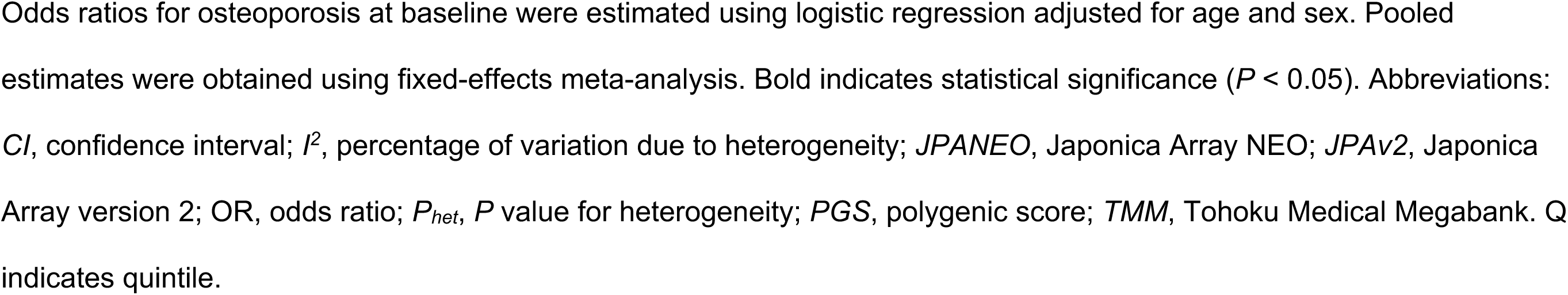
Osteoporosis prevalence and odds ratio stratified by PGS quintile in baseline study.

| | PGS<br>(quintile) | Number of<br>subjects<br>(n) | Number<br>of cases<br>(n) | Prevalence<br>(%) | OR | 95%CI | | P value | P for trend | Cochran's Q | I<sup>2</sup> (%) | $p_{het}$ |
| --- | --- | --- | --- | --- | --- | --- | --- | --- | --- | --- | --- | --- |
|  |  |  |  |  |  | Low | High |  |  |  |  |  |
| Validation<br>cohort I<br>(TMM9K-JPAv2,<br>N=7,609) | Q1 | 1,522 | 289 | 19.0 | 1.24 | 1.02 | 1.51 | **0.032** |  | – | – | – |
|  | Q2 | 1,522 | 273 | 17.9 | 1.17 | 0.96 | 1.43 | 0.116 |  | – | – | – |
|  | Q3 | 1,521 | 240 | 15.8 | 1.00<br>(Reference) | – | – | – | **2.37E-04** | – | – | – |
|  | Q4 | 1,522 | 263 | 17.3 | 1.06 | 0.87 | 1.30 | 0.556 |  | – | – | – |
|  | Q5 | 1,522 | 221 | 14.5 | 0.86 | 0.70 | 1.05 | 0.142 |  | – | – | – |
| Validation<br>cohort II<br>(TMM18K-JPANEO,<br>N=1,061) | Q1 | 213 | 49 | 23.0 | 1.02 | 0.64 | 1.62 | 0.940 |  | – | – | – |
|  | Q2 | 212 | 46 | 21.7 | 0.89 | 0.55 | 1.42 | 0.615 |  | – | – | – |
|  | Q3 | 212 | 49 | 23.1 | 1.00<br>(Reference) | – | – | – | 0.490 | – | – | – |
|  | Q4 | 212 | 45 | 21.2 | 0.92 | 0.57 | 1.47 | 0.714 |  | – | – | – |
|  | Q5 | 212 | 44 | 20.8 | 0.83 | 0.52 | 1.33 | 0.443 |  | – | – | – |
| Fixed-effect<br>meta-analysis | Q1 | – | – | – | 1.22 | 1.07 | 1.40 | **0.003** |  | 0.64 | 0.0 | 0.72 |
|  | Q2 | – | – | – | 1.15 | 1.00 | 1.31 | **0.047** |  | 1.26 | 0.0 | 0.53 |
|  | Q3 | – | – | – | 1.00<br>(Reference) | – | – | – | – | – | – | – |
|  | Q4 | – | – | – | 1.05 | 0.92 | 1.20 | 0.489 |  | 0.35 | 0.0 | 0.84 |
|  | Q5 | – | – | – | 0.85 | 0.74 | 0.98 | **0.027** |  | 0.01 | 0.0 | 0.99 |

Odds ratios for osteoporosis at baseline were estimated using logistic regression adjusted for age and sex. Pooled estimates were obtained using fixed-effects meta-analysis. Bold indicates statistical significance ( $P < 0.05$ ). Abbreviations: *CI*, confidence interval; $I^2$ , percentage of variation due to heterogeneity; *JPANEO*, Japonica Array NEO; *JPAv2*, Japonica Array version 2; OR, odds ratio; $P_{het}$ , $P$ value for heterogeneity; *PGS*, polygenic score; *TMM*, Tohoku Medical Megabank. Q indicates quintile.

### Osteoporosis genetic risk stratification by T-score PGS: Joint effects of age group, sex, and modifiable factors

To evaluate the influence of age and sex on the genetic risk of osteoporosis, we stratified participants at the clinically relevant threshold of 65 years of age, as recommended by osteoporosis guidelines for the Dual-energy X-ray Absorptiometry-based screening [35]. We compared the risk of osteoporosis across the genetic risk categories using individuals with intermediate (Q2–Q4) PGS as the reference group (Figure 3 and Supplementary Table S10). In the fixed-effect meta-analysis, younger individuals (< 65 years of age) with a lower genetic risk (Q5) showed significantly reduced odds of osteoporosis compared with the reference (OR = 0.74, 95% CI: 0.59–0.92, *P* = 0.006). Among older participants (≥ 65 years of age), those with a high genetic risk (Q1) exhibited significantly increased odds of osteoporosis compared to the reference (OR = 1.30, 95% CI: 1.04–1.62, *P* = 0.019). These findings indicate that a low genetic risk is more detectable in younger individuals, whereas a high genetic risk becomes evident in those aged ≥ 65 years.

**Figure 3.**
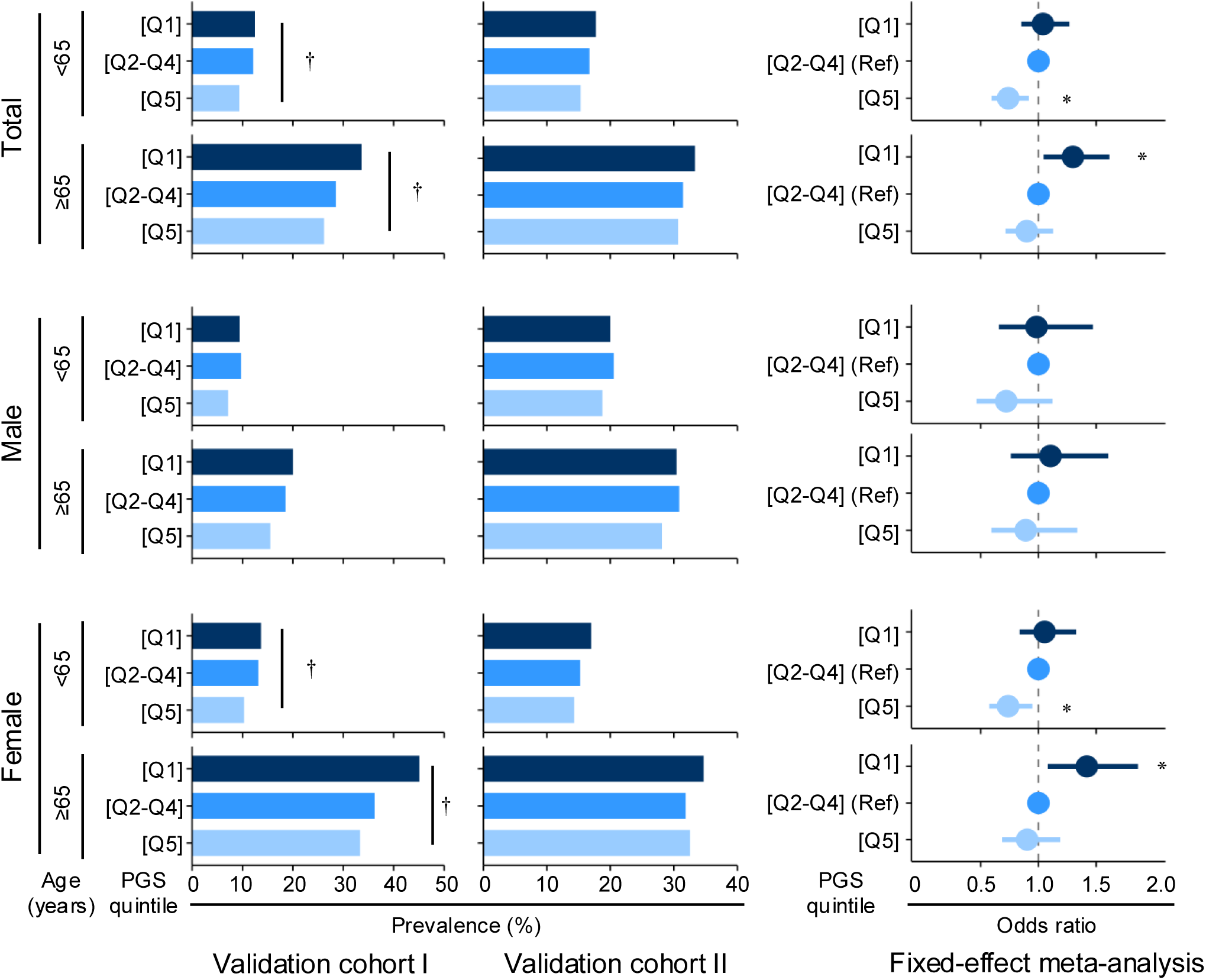
Osteoporosis prevalence and ORs stratified by age-stratified genetic risk groups. Participants were stratified by genetic risk (Q1: high, Q2–Q4: intermediate, Q5: low) and age (< 65 or ≥ 65 years). The individuals with the intermediate [Q2–Q4] PGS group served as reference. Osteoporosis prevalence (left and middle) and ORs (right) at baseline were estimated using logistic regression adjusted for age and sex, with pooled results obtained through fixed-effect meta-analysis. Bars indicate 95% CI. The bar color indicates the status of genetic risk levels. † indicates a significant trend across genetic risk groups (P < 0.05). * indicates a statistically significant pooled ORs from the fixed-effect meta-analysis (P < 0.05). Abbreviations: PGS, polygenic score; Q, quintile.

Next, we performed additional stratification according to sex. In women, the pattern was consistent with the overall findings: those with low genetic risk (Q5) had a significantly reduced odds of osteoporosis in the < 65 years of age group (OR = 0.74, 95% CI: 0.57–0.95, *P* = 0.017), whereas those ≥ 65 years with high genetic risk (Q1) showed increased odds compared with the reference group (OR = 1.42, 95% CI: 1.08–1.86, *P* = 0.012). However, in men, no significant differences in osteoporosis odds were observed across genetic risk categories within age groups.

Subsequently, we assessed the effects of modifiable lifestyle factors. Stratification by established risk factors, including obesity, smoking, high alcohol intake, and physical inactivity, did not show significant modification of genetic risk (Supplementary Figure S3 and Supplementary Table S11).

### Prospective cohort analysis

During the follow-up period, the corresponding person-years of observation for validation cohorts I and II were 16,226.8 and 1,901.6, respectively. During the follow-up period, 422 of 4,811 participants in validation cohort I and 116 of 581 in validation cohort II developed new cases (Supplementary Table S12). In both cohorts, the cumulative incidence of osteoporosis was highest in the genetic high-risk group (Q1) and lowest in the low-risk group (Q5), with the incidence rate showing a similar trend (Table 2). Notably, in the meta-analysis of IRR, the high-risk group (Q1) showed a significantly elevated risk compared with the intermediate group (Q2-Q4), whereas the low-risk group (Q5) showed a significantly reduced risk (Table 2).

**Table 2.**
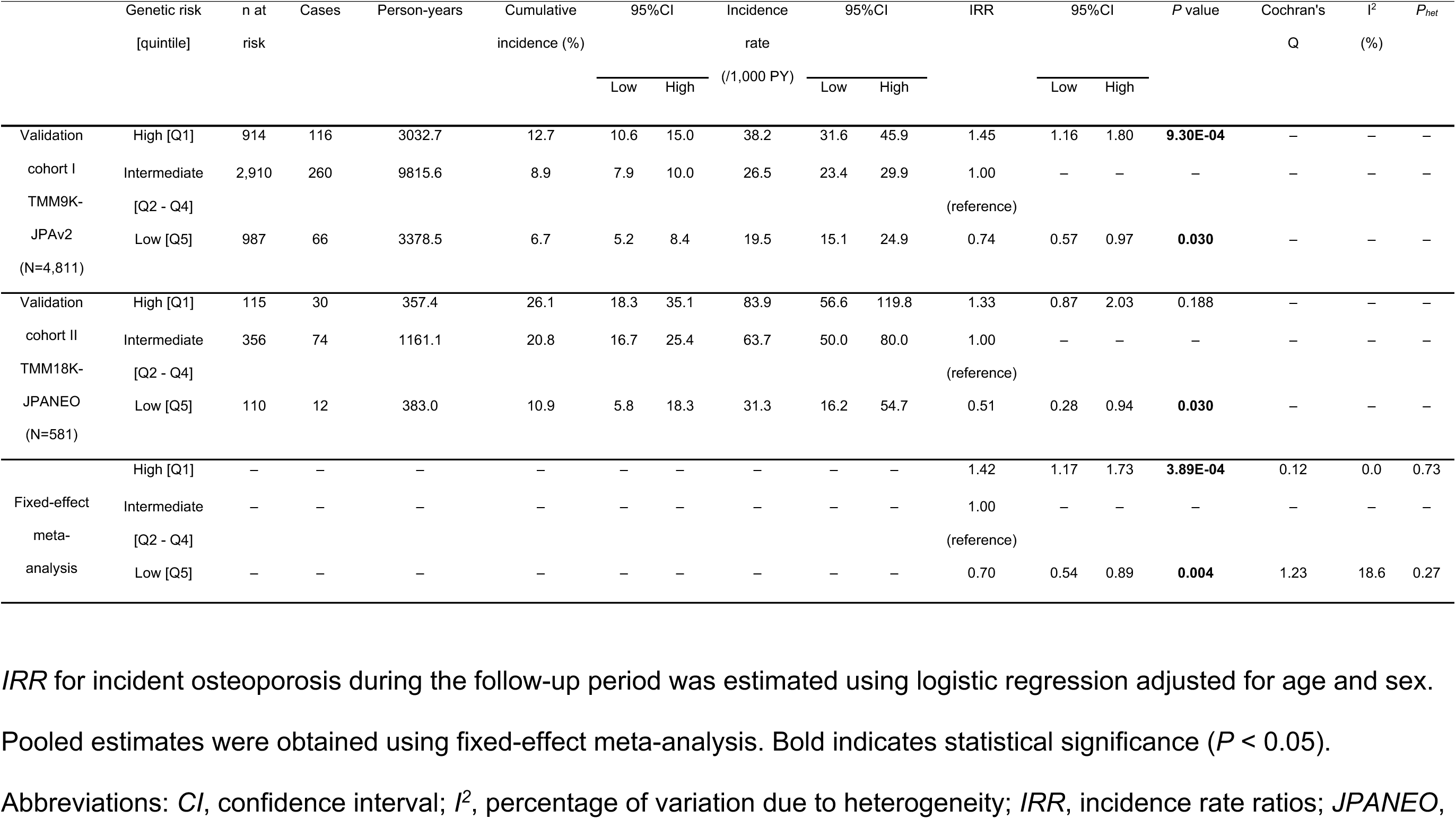
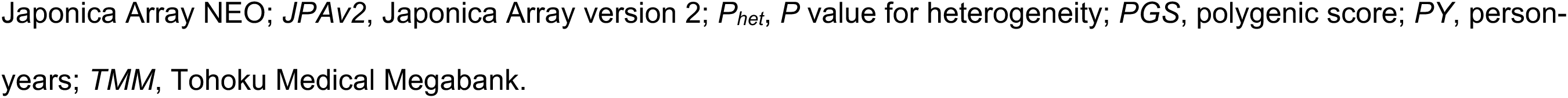
Incidence and risk estimates of osteoporosis by PGS-based genetic risk group in the follow-up study.

|  | Genetic risk | n at risk | Cases | Person-years | Cumulative incidence (%) | 95%CI |  | Incidence rate (/1,000 PY) | 95%CI |  | IRR | 95%CI |  | P value | Cochran's Q | I<sup>2</sup> (%) | P<sub>het</sub> |
| --- | --- | --- | --- | --- | --- | --- | --- | --- | --- | --- | --- | --- | --- | --- | --- | --- | --- |
|  | [quintile] |  |  |  |  | Low | High |  | Low | High |  | Low | High |  |  |  |  |
| Validation cohort I | High [Q1] | 914 | 116 | 3032.7 | 12.7 | 10.6 | 15.0 | 38.2 | 31.6 | 45.9 | 1.45 | 1.16 | 1.80 | **9.30E-04** | — | — | — |
| TMM9K-JPAv2 (N=4,811) | Intermediate [Q2 - Q4] | 2,910 | 260 | 9815.6 | 8.9 | 7.9 | 10.0 | 26.5 | 23.4 | 29.9 | 1.00 | — | — | — | — | — | — |
|  | Low [Q5] | 987 | 66 | 3378.5 | 6.7 | 5.2 | 8.4 | 19.5 | 15.1 | 24.9 | 0.74 | 0.57 | 0.97 | **0.030** | — | — | — |
| Validation cohort II | High [Q1] | 115 | 30 | 357.4 | 26.1 | 18.3 | 35.1 | 83.9 | 56.6 | 119.8 | 1.33 | 0.87 | 2.03 | 0.188 | — | — | — |
| TMM18K-JPANEO (N=581) | Intermediate [Q2 - Q4] | 356 | 74 | 1161.1 | 20.8 | 16.7 | 25.4 | 63.7 | 50.0 | 80.0 | 1.00 | — | — | — | — | — | — |
|  | Low [Q5] | 110 | 12 | 383.0 | 10.9 | 5.8 | 18.3 | 31.3 | 16.2 | 54.7 | 0.51 | 0.28 | 0.94 | **0.030** | — | — | — |
| Fixed-effect meta-analysis | High [Q1] | — | — | — | — | — | — | — | — | — | 1.42 | 1.17 | 1.73 | **3.89E-04** | 0.12 | 0.0 | 0.73 |
|  | Intermediate [Q2 - Q4] | — | — | — | — | — | — | — | — | — | 1.00 | — | — | — | — | — | — |
|  | Low [Q5] | — | — | — | — | — | — | — | — | — | 0.70 | 0.54 | 0.89 | **0.004** | 1.23 | 18.6 | 0.27 |

*IRR* for incident osteoporosis during the follow-up period was estimated using logistic regression adjusted for age and sex.
Pooled estimates were obtained using fixed-effect meta-analysis. Bold indicates statistical significance ( $P < 0.05$ ).
Abbreviations: *CI*, confidence interval; *I*<sup>2</sup>, percentage of variation due to heterogeneity; *IRR*, incidence rate ratios; *JPANEO*,
Japonica Array NEO; *JPAv2*, Japonica Array version 2; $P_{het}$ , $P$ value for heterogeneity; *PGS*, polygenic score; *PY*, person-years; *TMM*, Tohoku Medical Megabank.

### Interaction of age with genetic risk of T-score and osteoporosis

We examined whether the predictive effect of PGS on osteoporosis varied with age. Interaction terms for age × PGS were tested using logistic regression models with osteoporosis as the outcome, including sex as a covariate. Notably, none of the interaction terms reached statistical significance, indicating that the effect of PGS on osteoporosis was consistent across different ages (Supplementary Table S13).

Next, we assessed the correlation between baseline age and T-score separately by genetic risk group, comparing each group with the intermediate-risk group (Supplementary Figure S4). No significant differences were observed in either the high- or low-risk groups, and the slope of the association did not differ significantly between the groups (Supplementary Table S14). These results indicate that the PGS was not associated with the rate of age-related T-score decline.

Next, we used the fitted age–T-score regression lines to estimate group-specific mean T-scores at age 20 years as an approximation of peak bone mass. When extrapolating the linear regression model to age 20 years, the predicted mean T-score differed significantly by genetic risk group: relative to the intermediate-risk group, the low-risk group showed a positive deviation, whereas the high-risk group showed a negative deviation (Supplementary Table S15). Consistent with this pattern, analyses of observed T-scores at baseline among participants aged 20–44 years (young adult range) showed a modest, non-significant trend toward lower values in the high-risk group and significantly higher values in the low-risk group compared with the intermediate-risk group (Table 3). These findings indicate that high-genetic risk primarily lowers the BMD trajectory from early adulthood, instead of accelerating age-related bone loss, so that parallel declines over time cause these individuals to reach the osteoporotic range earlier.

**Table 3.**
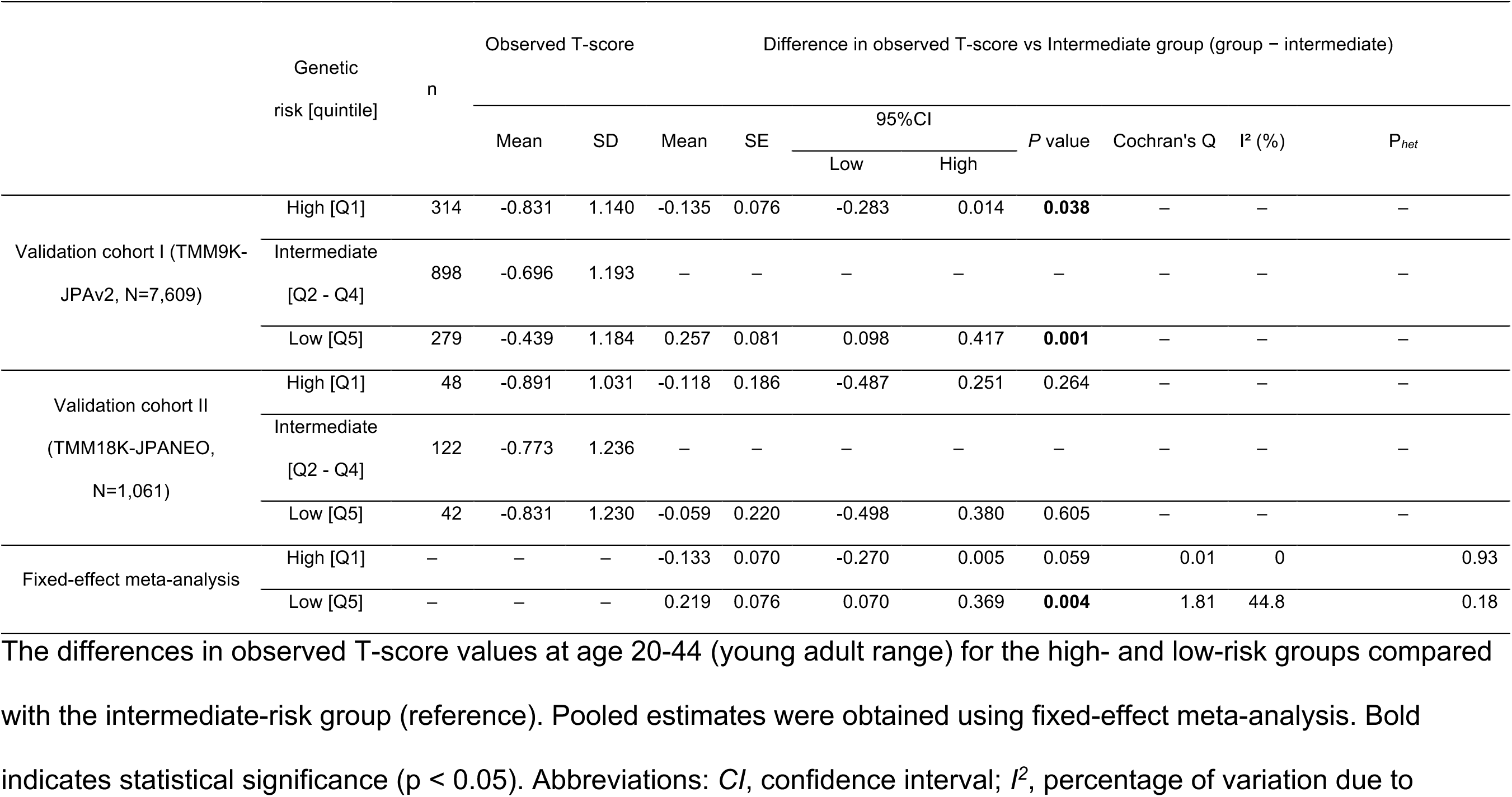
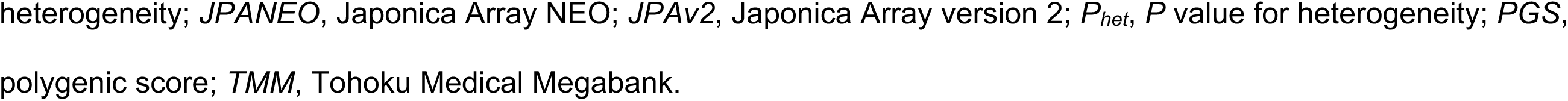
Differences in observed T-score at baseline (age 20-44, young adult range) for high- and low-generic risk groups versus the intermediate-risk group (Q2–Q4).

## Discussion

Most polygenic models for bone traits have been developed using European cohorts [5, 9, 10]. Because of ancestry differences, portability across populations remains a key challenge, despite recent efforts toward cross-ancestry models [36, 37]. Although our Japanese-specific model may not directly integrate into multi-ancestry frameworks, it provides one of the first large-scale polygenic tools for osteoporosis risk in East Asians and may serve as a valuable option in this population.

In the present study, we developed and validated a Japanese-specific PGS model for osteoporosis in a large-scale cohort. Because of known ancestry differences in allele frequencies, linkage disequilibrium structure, and environmental exposures, such population-tailored scores are likely to perform better than PGSs derived exclusively from European cohorts in East Asian populations. Our GWAS identified three loci, *MBL2, TMEM135*, and *WNT16*, which have also been reported in previous GWASs in both European and East Asian populations [5, 9, 38, 39]. The concordance of these loci across populations, together with their involvement in bone-metabolism pathways [8], supports the reliability of our GWAS and its use as a basis for constructing a PGS in East Asians.

The predictive performance of our model (AUC of approximately 0.73) was comparable to that of European-derived models [5, 10]. Our model explained approximately 20–24% of the variance in QUS-derived T-scores, a proportion consistent with previous large-scale studies using similar methods [9]. These results suggest that a population-matched PGS can provide complementary, genetically anchored information for risk stratification in East Asians. However, the incremental gain in discrimination beyond age and sex is modest, indicating that the current model is best viewed as an adjunct for stratification and hypothesis generation rather than a standalone clinical screening tool.

Although no statistically significant interaction between age and PGS was detected in logistic regression models, subgroup analyses revealed clinically meaningful patterns. Age-stratified odds ratios (ORs) indicated that individuals at high genetic risk had an increased likelihood of osteoporosis in older age, even in the absence of a formal interaction term. Together with the lack of differences in the slope of the age–T-score association across genetic risk groups, these findings indicate that the PGS is not strongly associated with the rate of age-related T-score decline.

Collectively, the observed age-stratified ORs and the contrasts in predicted and observed T-scores in young adults support a model in which high-genetic risk primarily lowers the bone mass trajectory from early adulthood, instead of accelerating age-related bone loss. Using this model, parallel age-related declines across PGS groups mean that individuals at high genetic risk are more likely to cross the osteoporotic threshold earlier in later life. This interpretation is in line with previous observations that many BMD-associated genetic variants exert stronger effects during the bone accrual phase in youth [7], and is concordant with pediatric data showing that a BMD-related polygenic score is associated with higher BMD and reduced fracture risk in childhood [40], as also highlighted by Fiscaletti and Manousaki [41].

This interpretation is clinically relevant. Peak bone mass has long-term implications for osteoporosis development, and individuals with a high genetic risk may inherently have a lower bone mass established during early adulthood. Therefore, early identification and intervention before the onset of bone loss may be the most effective. The absence of statistical interaction should not be viewed as evidence against clinical relevance; instead, our findings highlight the importance of age-stratified analyses and large-scale cohorts in detecting subtle yet meaningful gene-environment dynamics.

This study had certain limitations. First, data on prior osteoporosis treatment were unavailable, and participants undergoing therapy at baseline may not have been excluded. Because osteoporosis treatment can increase or stabilize bone measures, inclusion of treated individuals at baseline would be expected to attenuate associations and bias effect estimates toward the null, making our observed gradients conservative. Second, recruitment was limited to participants from health check-ups and community support centers in Miyagi and Iwate Prefectures, potentially introducing a health-conscious selection bias [20–22]. This may limit external generalizability; however, it is less likely to fully explain the monotonic risk gradient observed across PGS strata and its consistency across independent datasets and prospective follow-up. Third, key lifestyle factors (such as alcohol use, smoking, and physical activity) were assessed by self-report, which may have affected accuracy. In addition, most participants were middle-aged or older, and although lifestyle interventions can increase or maintain BMD to some extent even in older adults, their effects are generally considered small [1, 42]. This does not contrast with the importance of lifestyle, but may instead reflect the limited plasticity of BMD once peak bone mass has been established. Therefore, our findings support a prevention strategy that combines early identification of genetically susceptible individuals with lifelong optimization of lifestyle factors.

In conclusion, we established a polygenic score for osteoporosis risk in a Japanese cohort and validated its performance. Our findings indicate that this score primarily reflects genetically determined peak bone mass, such that high-risk individuals enter adulthood with lower BMD levels and, despite similar rates of decline, reach the osteoporotic threshold earlier. These observations support the use of polygenic information to identify susceptible individuals at an early stage and to focus bone-preserving lifestyle strategies particularly before the attainment of peak bone mass. However, further studies are required to determine whether PGS-based interventions initiated in childhood or adolescence can reduce the risk of osteoporosis. Although immediate clinical impact may be limited by phenotype definition and cohort constraints, our findings provide population-specific evidence that polygenic liability is linked to bone status established around peak bone mass, supporting a life-course prevention framework for East Asian populations.

## Data Availability

The QUS-derived T-score GWAS summary statistics are available at jMorp (https://jmorp.megabank.tohoku.ac.jp/gwas-studies/TGA000015). Individual-level data are not publicly available; access may be granted upon request and approval by the Information Distribution Review Committee of the TMM Project.

## Acknowledgements

This study was supported by the Tohoku Medical Megabank (TMM) Project (Special Account for the Reconstruction of the Great East Japan Earthquake) of the Ministry of Education, Culture, Sports, Science and Technology (MEXT) and the Japan Agency for Medical Research and Development (AMED; grant number JP23tm0124006). The supercomputer system in the TMM Project (grant number JP21tm0424601) was used for data analysis. YOY and AS were supported by the Japan Society for the Promotion of Science (JSPS) KAKENHI Grant JP23K09696. During the preparation of this manuscript, the authors used ChatGPT (OpenAI; GPT-4o) to assist with language editing, summarizing relevant literature, and improving code readability and debugging.

## Author contributions

Conceptualization: YOY, YS, TH, AS; Methodology: YOY, YS, TH, MN; Software: YOY, YS, TH; Formal analysis: YOY, YS, TH; Data curation: YOY, YS; Writing—original draft: YOY; Writing—review & editing: YOY, YS, TH, MN, SM, SK, HO, MS, AS.

## Conflict of Interest

TH is a board member of Genome Analytics Japan, Inc. The other authors declare no conflicts of interest associated with this manuscript.

## Supplementary information

The Supplementary information is available at the Journal of Human Genetics website.

## Notes

### Competing Interest Statement

T.H. is a board member of Genome Analytics Japan, Inc. The other authors declare no conflicts of interest associated with this manuscript.

### Funding Statement

This study was supported by the Tohoku Medical Megabank Project of AMED (JP23tm0124006; JP21tm0424601) and JSPS KAKENHI (JP23K09696, YOY and AS).

### Author Declarations

Institutional Review Board of Iwate Medical University gave ethical approval for this work (approval ID: HG H25-2).

### Summary of Updates

Revised the text and moved Table 1 to Supplementary Table 2.

