## Supplemental_information for "Osteoporosis Genetic Risk Prediction Using Bone Mineral Density Polygenic Scores in Japanese: TMM CommCohort Study"

### ReadMe: Supplementary Information

Title：

Contains Supplementary Figures S1–S4 and Supplementary Tables S1–S15 referenced in the main text, including additional model construction details, validation results, sensitivity analyses, and age-stratified analyses.

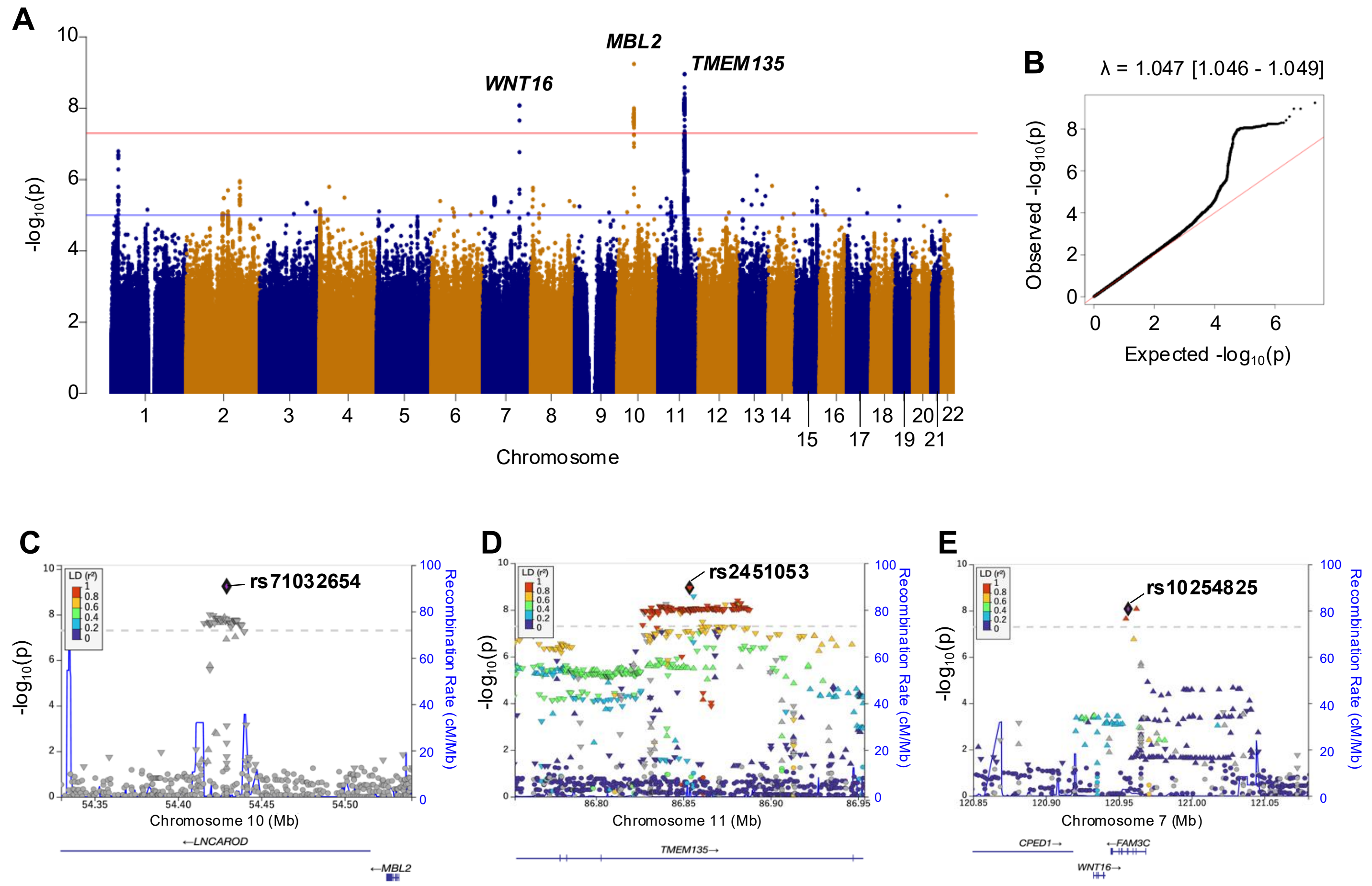


**Supplementary Fig. S1. GWAS results for T-score derived from QUS measurements.**

(A) Manhattan plot showing -$\log_{10}$ (p) for association with T-score across chromosomes. The horizonal red lines indicate the genome-wide significance threshold $P = 5 \times 10^{-8}$, and the blue lines indicate suggestive significance threshold $P = 1 \times 10^{-5}$. (B) Q–Q plot with observed vs. expected p-values. Genomic inflation factor (λ) is shown. (C–E). Regional association plots for lead variants: (C) rs71032654 at *MBL2* in chromosome 10, (D) rs2451053 at *TMEM135* in chromosome 11 and (E) rs10254825 at *WNT16* in chromosome 7. The lead variants within each locus are shown as purple diamonds. The colors indicate the extent of linkage disequilibrium (LD) ($r^2$) with lead variant based on EAS population from 1000 Genomes; recombination rates shown in blue. Plots generated using LocusZoom (v0.14.0) [30].

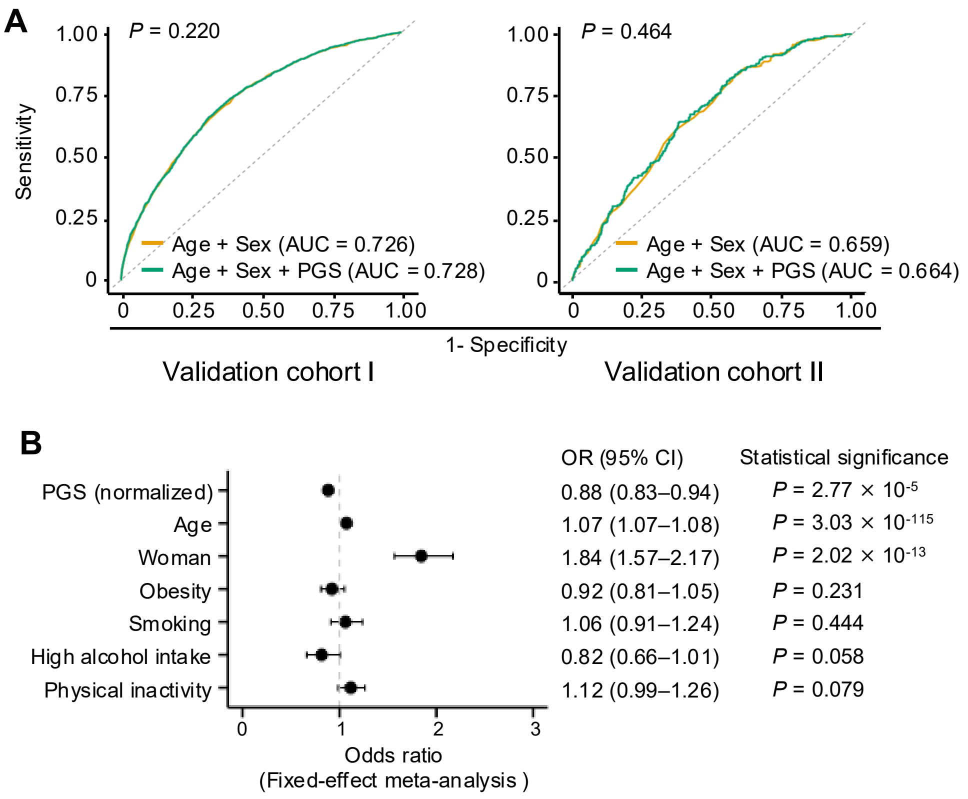


**Supplementary Figure S2. Predictive performance and association of T-score PGS with osteoporosis.** (A) Receiver operating characteristic (ROC) curves and area under the curve (AUC) for basic (age + sex) and PGS-enhanced models. The p value shown in the left corner represents the result of DeLong's test for comparing AUCs between models. (B) Multivariable logistic regression analyses of the association between the T-score PGS and osteoporosis. To address potential confounding, multivariable logistic regression models were used to estimate ORs for prevalent osteoporosis at baseline, including PGS, age, sex, obesity, smoking, high alcohol intake, and physical inactivity as covariates. Obesity was defined as BMI ≥25 kg/m²; smoking status as having smoked more than 100 cigarettes in total; high alcohol intake as ≥30 g/day; and physical inactivity as engaging in leisure-time physical activity fewer than three times per week. Cohort-specific estimates were pooled using a fixed-effect meta-analysis. CI, confidence interval; PGS, polygenic score.

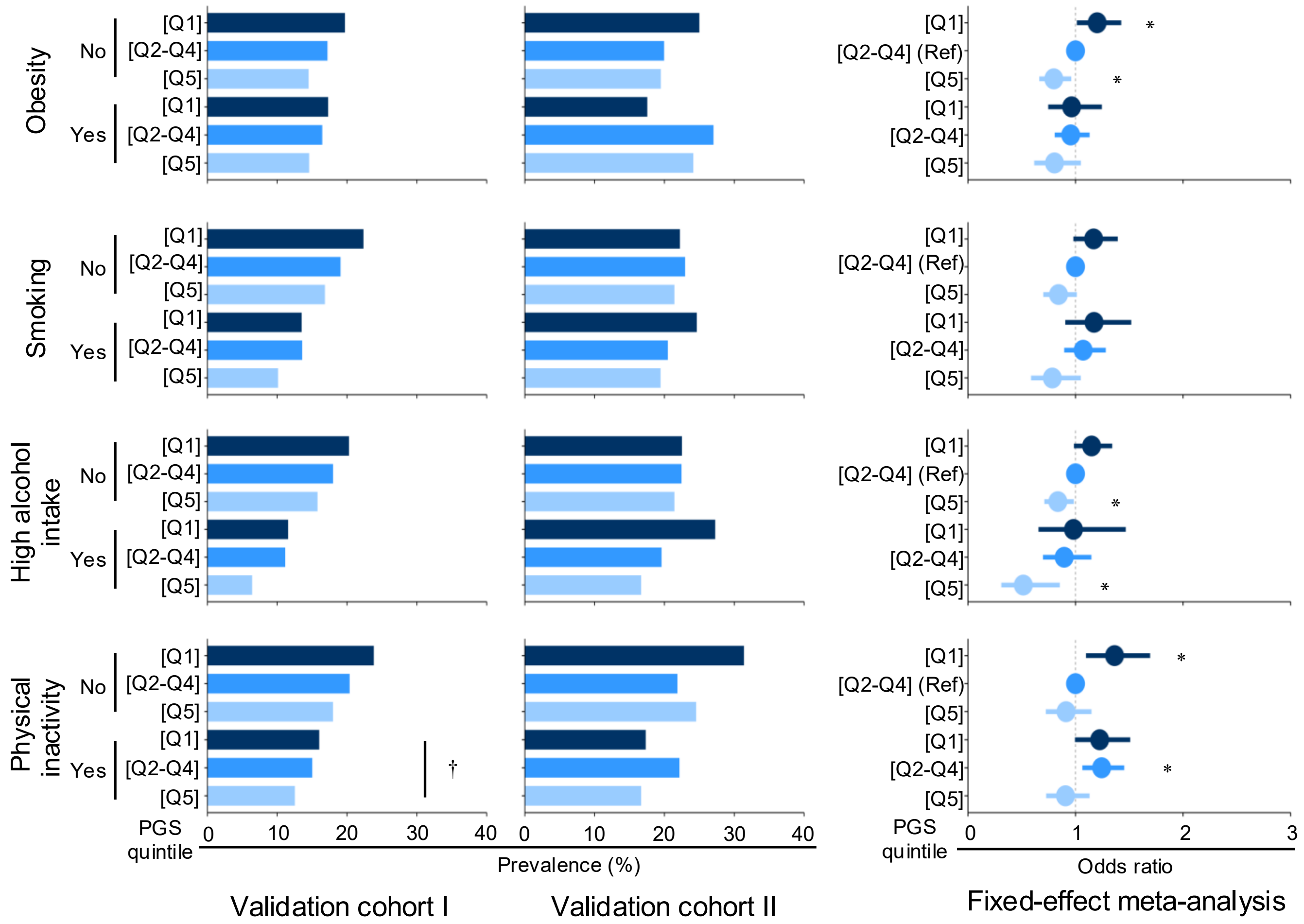


**Supplementary Figure S3. Osteoporosis prevalence and odds ratios by PGS-based genetic risk and modifiable lifestyle factors.**

Participants were stratified into six groups by genetic risk (Q1: high, Q2–Q4: intermediate, Q5: low) and the presence or absence of modifiable lifestyle factors (obesity, smoking, high alcohol intake, physical inactivity). Within each lifestyle category, the intermediate-risk group without the lifestyle risk factor served as the reference. Osteoporosis prevalence (left and middle panels) and odds ratios (right panel) at baseline were estimated using logistic regression adjusted for age and sex, with pooled results via fixed-effect meta-analysis. Bars indicate 95% confidence intervals. The bar color indicates the status of genetic risk levels. † indicates a significant trend across genetic risk groups ($P < 0.05$). * indicates a statistically significant pooled odds ratio from the fixed-effect meta-analysis ($P < 0.05$). PGS, polygenic score; Q, quintile.

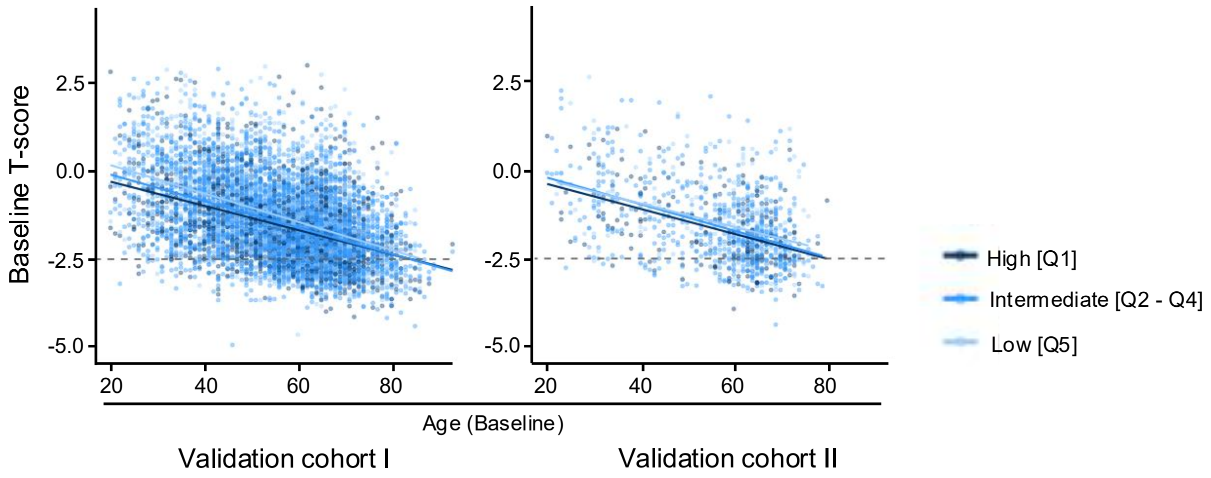


**Supplementary Figure S4. Scatter plots of baseline age and T-score, stratified by genetic risk group.**

Each point represents an individual participant. Linear regression lines are shown separately for the high-risk (dark blue), intermediate-risk (blue), and low-risk (light blue) groups. These models were used to compare the slopes of age–T-score associations across groups.

**Supplementary Table S1. Demographic characteristics of participants in each study cohort.**

| | GWAS cohort | Model-selection cohort | Validation cohort I | | | | | | Validation cohort II | | | | | |
|---|---|---|---|---|---|---|---|---|---|---|---|---|---|---|
| | TMM18K-JPAv2 | TMM9K-JPANEO | TMM9K-JPAv2 | | | | | | TMM18K-JPANEO | | | | | |
| | Baseline study | Baseline study | Baseline study | | | Follow-up study | | | Baseline study | | | Follow-up study | | |
| | | | Total | Male | Female | Total | Male | Female | Total | Male | Female | Total | Male | Female |
| N | 12,371 [c] | 1,419 | 7,609 | – | – | 4,811 | – | – | 1,061 | – | – | 581 | – | – |
| Women, N (%) | 9,345 (75.5) | 975 (68.7) | 5,067 (66.6) | – | – | 3,135 (65.2) | – | – | 733 (69.1) | – | – | 388 (66.8) | – | – |
| Age (years)[a] | 58.0 ± 13.0 | 62.5 ± 9.1 | 56.5 ± 13.2 | 58.6 ± 13.6 | 55.4 ± 12.9 | 59.2 ± 12.3 | 62.7 ± 12.5 | 57.3 ± 11.7 | 57.0 ± 13.4 | 60.3 ± 12.3 | 55.5 ± 13.7 | 60.6 ± 13.2 | 64.3 ± 12.3 | 58.8 ± 13.3 |
| T-score [a] | -1.56 ± 1.12 | -1.70 ± 0.97 | -1.46 ± 1.12 | -1.23 ± 1.15 | -1.58 ± 1.08 | -1.32 ± 1.03 | -1.11 ± 1.05 | -1.42 ± 1.00 | -1.61 ± 1.08 | -1.66 ± 1.07 | -1.59 ± 1.09 | -1.59 ± 1.04 | -1.60 ± 1.05 | -1.58 ± 1.04 |
| Height (cm)[a] | 158.4 ± 7.7 | 157.2 ± 8.2 | 159.1 ± 8.4 | 167.1 ± 6.5 | 155.0 ± 6.0 | 159.4 ± 8.1 | 166.9 ± 6.4 | 155.4 ± 5.8 | 158.7 ± 7.9 | 167.1 ± 6.0 | 155.0 ± 5.4 | 158.8 ± 8.1 | 166.9 ± 6.0 | 154.8 ± 5.6 |
| Weight (kg) [a] | 57.3 ± 10.7 | 57.9 ± 10.6 | 59.2 ± 11.3 | 67.6 ± 10.1 | 55.0 ± 9.4 | 60.3 ± 11.3 | 67.9 ± 10.3 | 56.2 ± 9.6 | 58.7 ± 11.2 | 67.3 ± 10.6 | 54.9 ± 9.1 | 58.6 ± 10.9 | 66.0 ± 9.9 | 54.9 ± 9.5 |
| BMI ($kg/m^2$) [a] | 22.8 ± 3.5 | 23.3 ± 3.4 | 23.3 ± 3.6 | 24.2 ± 3.2 | 22.9 ± 3.7 | 23.6 ± 3.6 | 24.3 ± 3.1 | 23.3 ± 3.7 | 23.3 ± 3.7 | 24.1 ± 3.4 | 22.9 ± 3.7 | 23.2 ± 3.6 | 23.6 ± 3.0 | 22.9 ± 3.8 |
| Follow-up periods (months) [a] | – | – | – | – | – | 42.4 ± 9.5 | 42.5 ± 9.4 | 42.3 ± 9.5 | – | – | – | 43.7 ± 11.0 | 42.9 ± 9.7 | 44.1 ± 11.6 |
| Array | JPAv2 | JPANEO | JPAv2 | | | | | | JPANEO | | | | | |
| Residency [b] | Miyagi | Iwate | Iwate | | | | | | Miyagi | | | | | |
| Recruited | Visited an assessment center in ToMMo | Visited an assessment center in IMM | Visited an assessment center in IMM | | | | | | Visited an assessment center in ToMMo | | | | | |

Abbreviations: *BMI*, body mass index; *GWAS*, genome-wide association study; *IMM*, Iwate Medical Megabank Organization; *JPANEO*, Japonica Array NEO; *JPAv2*, Japonica Array version 2; *TMM*, Tohoku Medical Megabank; *ToMMo*, Tohoku Medical Megabank Organization.

a, Mean ± standard deviation.

b, Prefecture in Japan.

c, Initial cohort size.

**Supplementary Table S2. Lead variants identified in the genome-wide association study of QUS-derived T-score in the Japanese population.**

| rsID (Lead SNP) | CHR | BP | Near Gene | TMM18K-JPAv2 | | | | | | | GCST006433 | | | | | | |
|---|---|---|---|---|---|---|---|---|---|---|---|---|---|---|---|---|---|
| | | | | EA | NEA | EAF | Beta | SE | *P* value | INFO | EA | NEA | EAF | Beta | SE | *P* value | INFO |
| rs71032654; rs200903639 | 10 | 54,429,192 | *MLB2* | TAC | TATATATATATATATATATACAC | 0.649 | 0.091 | 0.015 | **5.70E-10** | 0.935 | n.d. | n.d. | n.d. | n.d. | n.d. | n.d. | n.d. |
| rs2451053 | 11 | 86,853,548 | *TMEM135* | T | C | 0.678 | -0.088 | 0.014 | **1.10E-09** | 0.999 | T | C | 0.471 | -0.043 | 0.002 | **7.70E-97** | 0.998 |
| rs10254825 | 7 | 120,956,440 | *WNT16* | G | A | 0.445 | 0.081 | 0.014 | **8.30E-09** | 0.941 | G | A | 0.395 | 0.182 | 0.002 | **3.0E-1601** | 0.98 |

Variants were filtered based on minor allele frequency (MAF ≥0.01) and imputation INFO score (≥0.4). Effect alleles (EA) and non-effect alleles (NEA), along with corresponding effect sizes (Beta), from the GCST006433 GWAS summary statistics were re-aligned and recalculated using gnomAD v2.1 (GRCh37) to harmonize allele orientation with the TMM18K-JPAv2 dataset. Bold indicates statistical significance ($P < 5 \times 10^{-8}$).

Abbreviations: BP, base position; CHR, chromosome; EAF, effect allele frequency; INFO, imputation quality score; JPAv2, Japonica Array version 2；MBL2, mannose-binding lectin 2; n.d., not detected; SE, standard error; TMEM135, transmembrane protein 135; TMM, Tohoku Medical Megabank;

**Supplementary Table S3. Genome-wide significant ($P < 5 \times 10^{-8}$) and suggestive ($P < 1 \times 10^{-5}$) variants associated with QUS-derived T-scores in this study.**

| rsID | Variant ID | CHR | BP | EA | NEA | EAF | Beta | SE | *P* value | INFO | Annotation | Gene |
|---|---|---|---|---|---|---|---|---|---|---|---|---|
| rs71032654;rs200903639 | 10:54429192_TATATATATATATATATATACAC_TAC | 10 | 54429192 | TAC | TATATATATATATATATATACAC | 0.648691 | 0.0911713 | 0.0147278 | 5.70E-10 | 0.93485 | intragenic_variant | RP11-556E13.1 |
| rs2451053 | 11:86853548_C_T | 11 | 86853548 | T | C | 0.67794 | -0.0878018 | 0.0144546 | 1.10E-09 | 0.998624 | intron_variant | TMEM135 |
| rs578533 | 11:86853615_T_C | 11 | 86853615 | C | T | 0.677879 | -0.0877846 | 0.014453 | 1.10E-09 | 0.998807 | intron_variant | TMEM135 |
| rs57015442;rs796966343 | 11:86855797_GT_G | 11 | 86855797 | G | GT | 0.663604 | -0.0856291 | 0.0143371 | 2.60E-09 | 0.989448 | intron_variant | TMEM135 |
| rs147009946 | 11:86881074_C_CACACGTTATTTTATTAA | 11 | 86881074 | CACACGTTATTTTATTAA | C | 0.668619 | -0.0843409 | 0.0143379 | 3.90E-09 | 0.998455 | intron_variant | TMEM135 |
| rs653519 | 11:86881007_A_G | 11 | 86881007 | G | A | 0.669371 | -0.0837962 | 0.0143507 | 5.10E-09 | 0.998267 | intron_variant | TMEM135 |
| rs555190 | 11:86882874_T_C | 11 | 86882874 | C | T | 0.669307 | -0.0838662 | 0.0143492 | 5.10E-09 | 0.998451 | intron_variant | TMEM135 |
| rs12575986 | 11:86840058_T_<DEL> | 11 | 86840058 | <DEL> | T | 0.661649 | -0.0839693 | 0.0143938 | 5.50E-09 | 0.983476 | intron_variant | TMEM135 |
| rs34718412 | 11:86874059_CTT_CT | 11 | 86874059 | CT | CTT | 0.66645 | -0.0834644 | 0.0143338 | 5.80E-09 | 0.994902 | intron_variant | TMEM135 |
| rs479593 | 11:86878421_T_C | 11 | 86878421 | C | T | 0.669526 | -0.0835222 | 0.0143481 | 5.80E-09 | 0.998742 | intron_variant | TMEM135 |
| rs480660 | 11:86878561_C_G | 11 | 86878561 | G | C | 0.669473 | -0.0834999 | 0.0143485 | 5.80E-09 | 0.998489 | intron_variant | TMEM135 |
| rs537712 | 11:86880164_A_C | 11 | 86880164 | C | A | 0.669527 | -0.0835244 | 0.0143481 | 5.80E-09 | 0.998736 | intron_variant | TMEM135 |
| rs652171 | 11:86880711_T_C | 11 | 86880711 | C | T | 0.669527 | -0.0835254 | 0.0143481 | 5.80E-09 | 0.998735 | intron_variant | TMEM135 |
| rs35152231 | 11:86881050_CT_C | 11 | 86881050 | C | CT | 0.669527 | -0.083526 | 0.0143481 | 5.80E-09 | 0.998734 | intron_variant | TMEM135 |
| rs654405 | 11:86881179_G_C | 11 | 86881179 | C | G | 0.669528 | -0.0835266 | 0.0143481 | 5.80E-09 | 0.998733 | intron_variant | TMEM135 |
| rs654485 | 11:86881246_T_C | 11 | 86881246 | C | T | 0.669528 | -0.0835266 | 0.0143481 | 5.80E-09 | 0.998733 | intron_variant | TMEM135 |
| rs34575204 | 11:86881368_T_TC | 11 | 86881368 | TC | T | 0.669528 | -0.0835266 | 0.0143481 | 5.80E-09 | 0.998733 | intron_variant | TMEM135 |
| rs572433 | 11:86881621_C_T | 11 | 86881621 | T | C | 0.669528 | -0.0835272 | 0.0143481 | 5.80E-09 | 0.998733 | intron_variant | TMEM135 |
| rs34126459 | 11:86882384_T_TTTG | 11 | 86882384 | TTTG | T | 0.666845 | -0.0834754 | 0.0143299 | 5.80E-09 | 0.996733 | intron_variant | TMEM135 |
| rs560624 | 11:86863701_C_A | 11 | 86863701 | A | C | 0.659549 | -0.0825826 | 0.0142347 | 6.50E-09 | 0.995582 | intron_variant | TMEM135 |
| rs564877 | 11:86836625_G_A | 11 | 86836625 | A | G | 0.668418 | -0.0829477 | 0.0143214 | 6.90E-09 | 0.999808 | downstream_gene_variant | RP11-648O15.2 |
| rs2932120 | 11:86836692_G_A | 11 | 86836692 | A | G | 0.668418 | -0.0829477 | 0.0143214 | 6.90E-09 | 0.999808 | downstream_gene_variant | RP11-648O15.2 |
| rs612523 | 11:86836785_G_A | 11 | 86836785 | A | G | 0.668418 | -0.0829477 | 0.0143214 | 6.90E-09 | 0.999808 | downstream_gene_variant | RP11-648O15.2 |
| rs34601982 | 11:86837184_G_GA | 11 | 86837184 | GA | G | 0.668418 | -0.0829477 | 0.0143214 | 6.90E-09 | 0.999808 | downstream_gene_variant | RP11-648O15.2 |
| rs628686 | 11:86838100_T_C | 11 | 86838100 | C | T | 0.668418 | -0.0829477 | 0.0143214 | 6.90E-09 | 0.999808 | intron_variant | TMEM135 |
| rs12574813 | 11:86840057_GT_G | 11 | 86840057 | G | GT | 0.666505 | -0.0831081 | 0.0143455 | 6.90E-09 | 0.995106 | intron_variant | TMEM135 |
| rs563019 | 11:86836408_G_A | 11 | 86836408 | A | G | 0.668354 | -0.0829356 | 0.0143227 | 7.00E-09 | 0.99952 | downstream_gene_variant | RP11-648O15.2 |
| rs487366 | 11:86837863_A_C | 11 | 86837863 | C | A | 0.668407 | -0.0829195 | 0.0143215 | 7.00E-09 | 0.999767 | intron_variant | TMEM135 |
| rs487370 | 11:86837865_A_C | 11 | 86837865 | C | A | 0.668407 | -0.0829195 | 0.0143215 | 7.00E-09 | 0.999767 | intron_variant | TMEM135 |
| rs649693 | 11:86869577_T_C | 11 | 86869577 | C | T | 0.662672 | -0.0833151 | 0.0144063 | 7.30E-09 | 0.979991 | intron_variant | TMEM135 |
| rs60438604;rs796962235 | 11:86840973_TA_T | 11 | 86840973 | T | TA | 0.667608 | -0.0828055 | 0.0143148 | 7.40E-09 | 0.999117 | intron_variant | TMEM135 |
| rs5793239;rs796340568 | 11:86838920_GA_G | 11 | 86838920 | G | GA | 0.668288 | -0.0827022 | 0.0143143 | 7.60E-09 | 0.999998 | intron_variant | TMEM135 |
| rs2048973 | 11:86839097_G_T | 11 | 86839097 | T | G | 0.668288 | -0.0827022 | 0.0143143 | 7.60E-09 | 0.999998 | intron_variant | TMEM135 |
| rs655050 | 11:86839381_T_C | 11 | 86839381 | C | T | 0.668288 | -0.0827022 | 0.0143143 | 7.60E-09 | 0.999998 | intron_variant | TMEM135 |
| rs655907 | 11:86839536_G_T | 11 | 86839536 | T | G | 0.668288 | -0.0827022 | 0.0143143 | 7.60E-09 | 0.999998 | intron_variant | TMEM135 |
| rs35418233;rs71040293 | 11:86839947_TAC_T | 11 | 86839947 | T | TAC | 0.668288 | -0.0827022 | 0.0143143 | 7.60E-09 | 0.999998 | intron_variant | TMEM135 |
| rs491895 | 11:86840098_G_C | 11 | 86840098 | C | G | 0.668288 | -0.0827022 | 0.0143143 | 7.60E-09 | 0.999998 | intron_variant | TMEM135 |
| rs595930 | 11:86841969_C_T | 11 | 86841969 | T | C | 0.668288 | -0.0827022 | 0.0143143 | 7.60E-09 | 0.999998 | intron_variant | TMEM135 |
| rs520755 | 11:86883454_A_G | 11 | 86883454 | G | A | 0.667811 | -0.0827128 | 0.014319 | 7.80E-09 | 0.999742 | intron_variant | TMEM135 |
| rs476597 | 11:86884018_C_T | 11 | 86884018 | T | C | 0.667812 | -0.0827118 | 0.014319 | 7.80E-09 | 0.999739 | intron_variant | TMEM135 |
| rs502580 | 11:86884532_G_T | 11 | 86884532 | T | G | 0.667812 | -0.0827124 | 0.014319 | 7.80E-09 | 0.999739 | intron_variant | TMEM135 |
| rs603140 | 11:86884615_G_A | 11 | 86884615 | A | G | 0.667812 | -0.0827124 | 0.014319 | 7.80E-09 | 0.999739 | intron_variant | TMEM135 |
| rs604035 | 11:86884825_C_G | 11 | 86884825 | G | C | 0.667812 | -0.0827129 | 0.014319 | 7.80E-09 | 0.999739 | intron_variant | TMEM135 |
| rs606299 | 11:86885342_T_C | 11 | 86885342 | C | T | 0.667812 | -0.0827129 | 0.014319 | 7.80E-09 | 0.999739 | intron_variant | TMEM135 |
| rs3133321 | 11:86831635_T_C | 11 | 86831635 | C | T | 0.668154 | -0.0829001 | 0.0143673 | 7.90E-09 | 0.990807 | upstream_gene_variant | RP11-648O15.2 |
| rs552521 | 11:86882599_C_A | 11 | 86882599 | A | C | 0.667739 | -0.0826775 | 0.01432 | 7.90E-09 | 0.99946 | intron_variant | TMEM135 |
| rs604199 | 11:86866260_C_T | 11 | 86866260 | T | C | 0.665656 | -0.0826726 | 0.0143381 | 8.00E-09 | 0.992814 | intron_variant | TMEM135 |
| rs10254825 | 7:120956440_A_G | 7 | 120956440 | G | A | 0.444551 | 0.0812378 | 0.0139542 | 8.30E-09 | 0.941277 | intergenic_region | CPED1:WNT16 |
| rs2908007 | 7:120962164_A_G | 7 | 120962164 | G | A | 0.451038 | 0.078498 | 0.013539 | 8.30E-09 | 1 | intergenic_region | CPED1:WNT16 |
| rs502079 | 11:86842812_T_C | 11 | 86842812 | C | T | 0.66755 | -0.0826015 | 0.0143273 | 8.30E-09 | 0.998313 | intron_variant | TMEM135 |
| rs609914 | 11:86842813_G_A | 11 | 86842813 | A | G | 0.66755 | -0.0826015 | 0.0143273 | 8.30E-09 | 0.998313 | intron_variant | TMEM135 |
| rs500312 | 11:86842992_A_G | 11 | 86842992 | G | A | 0.667936 | -0.0824763 | 0.0143125 | 8.30E-09 | 0.999938 | intron_variant | TMEM135 |
| rs482339 | 11:86857174_C_A | 11 | 86857174 | A | C | 0.667917 | -0.0825508 | 0.01432 | 8.30E-09 | 0.999563 | intron_variant | TMEM135 |
| rs495238 | 11:86882981_A_G | 11 | 86882981 | G | A | 0.66748 | -0.0825996 | 0.0143199 | 8.30E-09 | 0.999196 | intron_variant | TMEM135 |
| rs11424291;rs398045367 | 11:86848712_A_AT | 11 | 86848712 | AT | A | 0.667352 | -0.0824992 | 0.0143371 | 8.40E-09 | 0.996232 | intron_variant | TMEM135 |
| rs597319 | 11:86853997_A_G | 11 | 86853997 | G | A | 0.667919 | -0.0824623 | 0.0143198 | 8.60E-09 | 0.999522 | intron_variant | TMEM135 |
| rs492896 | 11:86882043_G_A | 11 | 86882043 | A | G | 0.668231 | -0.082484 | 0.0143232 | 8.60E-09 | 0.999733 | intron_variant | TMEM135 |
| rs603676 | 11:86866109_T_C | 11 | 86866109 | C | T | 0.660055 | -0.0828553 | 0.014377 | 8.70E-09 | 0.979527 | intron_variant | TMEM135 |
| rs608966 | 11:86854309_G_C | 11 | 86854309 | C | G | 0.667958 | -0.0823858 | 0.0143195 | 8.80E-09 | 0.999617 | intron_variant | TMEM135 |
| rs620303 | 11:86861127_C_T | 11 | 86861127 | T | C | 0.667866 | -0.0824506 | 0.0143302 | 8.80E-09 | 0.998102 | intron_variant | TMEM135 |
| rs2512490 | 11:86886175_T_G | 11 | 86886175 | G | T | 0.667682 | -0.0824084 | 0.0143211 | 8.80E-09 | 0.99936 | intron_variant | TMEM135 |
| rs202144782 | 11:86857361_GATCA_G | 11 | 86857361 | G | GATCA | 0.667983 | -0.0823428 | 0.0143181 | 8.90E-09 | 0.999846 | intron_variant | TMEM135 |
| rs765195417 | 11:86857366_GCCTGAGGGAATGT_G | 11 | 86857366 | G | GCCTGAGGGAATGT | 0.667983 | -0.0823433 | 0.0143181 | 8.90E-09 | 0.999849 | intron_variant | TMEM135 |
| rs670125 | 11:86857447_C_G | 11 | 86857447 | G | C | 0.667976 | -0.0823482 | 0.014318 | 8.90E-09 | 0.999897 | intron_variant | TMEM135 |
| rs568905 | 11:86857498_T_C | 11 | 86857498 | C | T | 0.667971 | -0.082351 | 0.0143179 | 8.90E-09 | 0.99993 | intron_variant | TMEM135 |
| rs670857 | 11:86857564_C_A | 11 | 86857564 | A | C | 0.667015 | -0.0822715 | 0.0143069 | 8.90E-09 | 0.998595 | intron_variant | TMEM135 |
| rs111307209 | 11:86858553_G_GTTACTTT | 11 | 86858553 | GTTACTTT | G | 0.667965 | -0.0823554 | 0.0143177 | 8.90E-09 | 0.999979 | intron_variant | TMEM135 |
| rs687208 | 11:86858868_T_C | 11 | 86858868 | C | T | 0.667964 | -0.082354 | 0.0143178 | 8.90E-09 | 0.999977 | intron_variant | TMEM135 |
| rs534009 | 11:86859033_T_C | 11 | 86859033 | C | T | 0.667965 | -0.0823554 | 0.0143177 | 8.90E-09 | 0.999982 | intron_variant | TMEM135 |
| rs533056 | 11:86859142_T_C | 11 | 86859142 | C | T | 0.667965 | -0.0823554 | 0.0143177 | 8.90E-09 | 0.999982 | intron_variant | TMEM135 |
| rs477944 | 11:86860509_G_A | 11 | 86860509 | A | G | 0.667964 | -0.0823562 | 0.0143177 | 8.90E-09 | 0.999993 | intron_variant | TMEM135 |
| rs607619 | 11:86860551_T_G | 11 | 86860551 | G | T | 0.667963 | -0.0823566 | 0.0143177 | 8.90E-09 | 0.999995 | intron_variant | TMEM135 |
| rs522439 | 11:86861753_T_A | 11 | 86861753 | A | T | 0.667963 | -0.0823554 | 0.0143177 | 8.90E-09 | 0.999995 | intron_variant | TMEM135 |
| rs636159 | 11:86862397_G_T | 11 | 86862397 | T | G | 0.667964 | -0.0823547 | 0.0143178 | 8.90E-09 | 0.999992 | intron_variant | TMEM135 |
| rs576582 | 11:86863374_A_G | 11 | 86863374 | G | A | 0.66797 | -0.0823473 | 0.0143182 | 8.90E-09 | 0.999969 | intron_variant | TMEM135 |
| rs561397 | 11:86863761_A_G | 11 | 86863761 | G | A | 0.66797 | -0.0823473 | 0.0143182 | 8.90E-09 | 0.999969 | intron_variant | TMEM135 |
| rs566046 | 11:86864277_A_G | 11 | 86864277 | G | A | 0.66797 | -0.0823473 | 0.0143182 | 8.90E-09 | 0.999969 | intron_variant | TMEM135 |
| rs482258 | 11:86864892_A_G | 11 | 86864892 | G | A | 0.66797 | -0.0823473 | 0.0143182 | 8.90E-09 | 0.999969 | intron_variant | TMEM135 |
| rs542678 | 11:86866652_A_G | 11 | 86866652 | G | A | 0.66797 | -0.0823473 | 0.0143182 | 8.90E-09 | 0.999969 | intron_variant | TMEM135 |
| rs542697 | 11:86866658_A_T | 11 | 86866658 | T | A | 0.66797 | -0.0823473 | 0.0143182 | 8.90E-09 | 0.999969 | intron_variant | TMEM135 |
| rs543687 | 11:86866802_G_A | 11 | 86866802 | A | G | 0.66797 | -0.0823473 | 0.0143182 | 8.90E-09 | 0.999969 | intron_variant | TMEM135 |
| rs608533 | 11:86854217_C_G | 11 | 86854217 | G | C | 0.668002 | -0.0823295 | 0.0143183 | 9.00E-09 | 0.999746 | intron_variant | TMEM135 |
| rs608870 | 11:86854250_C_T | 11 | 86854250 | T | C | 0.668002 | -0.0823295 | 0.0143183 | 9.00E-09 | 0.999747 | intron_variant | TMEM135 |
| rs609819 | 11:86854505_T_A | 11 | 86854505 | A | T | 0.667933 | -0.0823227 | 0.0143197 | 9.00E-09 | 0.999409 | intron_variant | TMEM135 |
| rs610228 | 11:86854595_G_T | 11 | 86854595 | T | G | 0.668002 | -0.0823295 | 0.0143183 | 9.00E-09 | 0.999748 | intron_variant | TMEM135 |
| rs611523 | 11:86854857_G_T | 11 | 86854857 | T | G | 0.668002 | -0.0823295 | 0.0143183 | 9.00E-09 | 0.999748 | intron_variant | TMEM135 |
| rs611579 | 11:86854903_C_T | 11 | 86854903 | T | C | 0.668002 | -0.0823295 | 0.0143183 | 9.00E-09 | 0.999748 | intron_variant | TMEM135 |
| rs541998 | 11:86855296_G_A | 11 | 86855296 | A | G | 0.668001 | -0.08233 | 0.0143183 | 9.00E-09 | 0.99975 | intron_variant | TMEM135 |
| rs541089 | 11:86855391_A_G | 11 | 86855391 | G | A | 0.668001 | -0.08233 | 0.0143183 | 9.00E-09 | 0.99975 | intron_variant | TMEM135 |
| rs626897 | 11:86856021_G_C | 11 | 86856021 | C | G | 0.668001 | -0.08233 | 0.0143183 | 9.00E-09 | 0.999752 | intron_variant | TMEM135 |
| rs654569 | 11:86856206_C_T | 11 | 86856206 | T | C | 0.668001 | -0.08233 | 0.0143183 | 9.00E-09 | 0.999752 | intron_variant | TMEM135 |
| rs655778 | 11:86856437_G_T | 11 | 86856437 | T | G | 0.668001 | -0.08233 | 0.0143183 | 9.00E-09 | 0.999752 | intron_variant | TMEM135 |
| rs486739 | 11:86856763_C_T | 11 | 86856763 | T | C | 0.668 | -0.0823306 | 0.0143183 | 9.00E-09 | 0.999753 | intron_variant | TMEM135 |
| rs618926 | 11:86867252_C_G | 11 | 86867252 | G | C | 0.667976 | -0.08234 | 0.0143185 | 9.00E-09 | 0.999951 | intron_variant | TMEM135 |
| rs563688 | 11:86867479_C_A | 11 | 86867479 | A | C | 0.667979 | -0.0823362 | 0.0143187 | 9.00E-09 | 0.99994 | intron_variant | TMEM135 |
| rs632708 | 11:86868033_C_T | 11 | 86868033 | T | C | 0.667988 | -0.0823254 | 0.0143192 | 9.00E-09 | 0.999918 | intron_variant | TMEM135 |
| rs576679 | 11:86868071_G_A | 11 | 86868071 | A | G | 0.667995 | -0.0823361 | 0.0143194 | 9.00E-09 | 0.999891 | intron_variant | TMEM135 |
| rs556139 | 11:86868347_C_T | 11 | 86868347 | T | C | 0.667989 | -0.0823234 | 0.0143192 | 9.00E-09 | 0.999915 | intron_variant | TMEM135 |
| rs495668 | 11:86868965_C_T | 11 | 86868965 | T | C | 0.667989 | -0.0823234 | 0.0143192 | 9.00E-09 | 0.999915 | intron_variant | TMEM135 |
| rs35695205;rs796768493 | 11:86869679_AT_A | 11 | 86869679 | A | AT | 0.667989 | -0.0823234 | 0.0143192 | 9.00E-09 | 0.999915 | intron_variant | TMEM135 |
| rs494698 | 11:86870469_G_A | 11 | 86870469 | A | G | 0.667989 | -0.0823234 | 0.0143192 | 9.00E-09 | 0.999915 | intron_variant | TMEM135 |
| rs664398 | 11:86870511_T_C | 11 | 86870511 | C | T | 0.667989 | -0.0823234 | 0.0143192 | 9.00E-09 | 0.999915 | intron_variant | TMEM135 |
| rs580120 | 11:86870848_A_T | 11 | 86870848 | T | A | 0.667989 | -0.0823234 | 0.0143192 | 9.00E-09 | 0.999915 | intron_variant | TMEM135 |
| rs513662 | 11:86872104_A_G | 11 | 86872104 | G | A | 0.667989 | -0.0823234 | 0.0143192 | 9.00E-09 | 0.999915 | intron_variant | TMEM135 |
| rs547867 | 11:86872647_G_T | 11 | 86872647 | T | G | 0.667989 | -0.0823234 | 0.0143192 | 9.00E-09 | 0.999915 | intron_variant | TMEM135 |
| rs577329 | 11:86873570_G_A | 11 | 86873570 | A | G | 0.667989 | -0.0823247 | 0.0143192 | 9.00E-09 | 0.999914 | intron_variant | TMEM135 |
| rs146959550 | 11:86873599_C_CT | 11 | 86873599 | CT | C | 0.667989 | -0.0823247 | 0.0143192 | 9.00E-09 | 0.999914 | intron_variant | TMEM135 |
| rs60027610;rs796641596 | 11:86877799_CT_C | 11 | 86877799 | C | CT | 0.66799 | -0.0823319 | 0.0143193 | 9.00E-09 | 0.999902 | intron_variant | TMEM135 |
| rs484135 | 11:86878332_G_C | 11 | 86878332 | C | G | 0.66799 | -0.0823325 | 0.0143193 | 9.00E-09 | 0.999901 | intron_variant | TMEM135 |
| rs484403 | 11:86878971_T_C | 11 | 86878971 | C | T | 0.667991 | -0.0823337 | 0.0143193 | 9.00E-09 | 0.999899 | intron_variant | TMEM135 |
| rs624004 | 11:86878994_G_C | 11 | 86878994 | C | G | 0.667991 | -0.0823337 | 0.0143193 | 9.00E-09 | 0.999899 | intron_variant | TMEM135 |
| rs511755 | 11:86879011_T_C | 11 | 86879011 | C | T | 0.667991 | -0.0823343 | 0.0143193 | 9.00E-09 | 0.999899 | intron_variant | TMEM135 |
| rs509277 | 11:86879342_C_T | 11 | 86879342 | T | C | 0.667991 | -0.0823349 | 0.0143193 | 9.00E-09 | 0.999898 | intron_variant | TMEM135 |
| rs570716 | 11:86881464_G_T | 11 | 86881464 | T | G | 0.667992 | -0.0823385 | 0.0143193 | 9.00E-09 | 0.999891 | intron_variant | TMEM135 |
| rs651961 | 11:86863609_C_T | 11 | 86863609 | T | C | 0.667892 | -0.0823211 | 0.0143192 | 9.10E-09 | 0.999667 | intron_variant | TMEM135 |
| rs607894 | 11:86877716_T_C | 11 | 86877716 | C | T | 0.667747 | -0.0823422 | 0.0143235 | 9.10E-09 | 0.99921 | intron_variant | TMEM135 |
| rs489247 | 11:86881641_A_G | 11 | 86881641 | G | A | 0.648503 | -0.081926 | 0.0142278 | 9.10E-09 | 0.99085 | intron_variant | TMEM135 |
| rs540403 | 11:86880458_G_A | 11 | 86880458 | A | G | 0.667854 | -0.0822894 | 0.0143214 | 9.20E-09 | 0.999619 | intron_variant | TMEM135 |
| rs2451060 | 11:86886408_C_T | 11 | 86886408 | T | C | 0.667646 | -0.0823125 | 0.0143207 | 9.20E-09 | 0.999379 | intron_variant | TMEM135 |
| rs546229 | 11:86872504_C_T | 11 | 86872504 | T | C | 0.66772 | -0.0822759 | 0.014325 | 9.30E-09 | 0.998752 | intron_variant | TMEM135 |
| rs687244 | 11:86852333_G_A | 11 | 86852333 | A | G | 0.668132 | -0.0821964 | 0.0143164 | 9.40E-09 | 0.999879 | intron_variant | TMEM135 |
| rs687651 | 11:86852427_G_T | 11 | 86852427 | T | G | 0.668128 | -0.0822009 | 0.0143165 | 9.40E-09 | 0.999857 | intron_variant | TMEM135 |
| rs499388 | 11:86852569_T_G | 11 | 86852569 | G | T | 0.668124 | -0.0822069 | 0.0143167 | 9.40E-09 | 0.999827 | intron_variant | TMEM135 |
| rs688542 | 11:86852593_G_A | 11 | 86852593 | A | G | 0.668123 | -0.0822078 | 0.0143167 | 9.40E-09 | 0.999823 | intron_variant | TMEM135 |
| rs497592 | 11:86852758_A_G | 11 | 86852758 | G | A | 0.668121 | -0.0822092 | 0.0143168 | 9.40E-09 | 0.999812 | intron_variant | TMEM135 |
| rs581833 | 11:86852877_A_T | 11 | 86852877 | T | A | 0.668121 | -0.0822092 | 0.0143168 | 9.40E-09 | 0.999811 | intron_variant | TMEM135 |
| rs593243 | 11:86853125_T_G | 11 | 86853125 | G | T | 0.668121 | -0.0822092 | 0.0143168 | 9.40E-09 | 0.999811 | intron_variant | TMEM135 |
| rs492211 | 11:86853282_G_A | 11 | 86853282 | A | G | 0.668121 | -0.0822104 | 0.0143168 | 9.40E-09 | 0.99981 | intron_variant | TMEM135 |
| rs150087331;rs59786778 | 11:86842383_A_AATCCCCTGTGCTCTACCTATTC | 11 | 86842383 | AATCCCCTGTGCTCTACCTATTC | A | 0.668146 | -0.0821772 | 0.0143158 | 9.50E-09 | 0.999985 | intron_variant | TMEM135 |
| rs502165 | 11:86842783_C_A | 11 | 86842783 | A | C | 0.668146 | -0.0821772 | 0.0143158 | 9.50E-09 | 0.999985 | intron_variant | TMEM135 |
| rs34935722;rs397712833 | 11:86843865_GT_G | 11 | 86843865 | G | GT | 0.668146 | -0.0821781 | 0.0143159 | 9.50E-09 | 0.999983 | intron_variant | TMEM135 |
| rs7949278 | 11:86844422_C_A | 11 | 86844422 | A | C | 0.668146 | -0.0821781 | 0.0143159 | 9.50E-09 | 0.999982 | intron_variant | TMEM135 |
| rs7949665 | 11:86844578_G_C | 11 | 86844578 | C | G | 0.668146 | -0.0821781 | 0.0143159 | 9.50E-09 | 0.999982 | intron_variant | TMEM135 |
| rs36070851;rs398045366 | 11:86844841_CT_C | 11 | 86844841 | C | CT | 0.668146 | -0.0821781 | 0.0143159 | 9.50E-09 | 0.999981 | intron_variant | TMEM135 |
| rs550988 | 11:86844886_A_T | 11 | 86844886 | T | A | 0.668146 | -0.0821781 | 0.0143159 | 9.50E-09 | 0.999981 | intron_variant | TMEM135 |
| rs398016991;rs5793240 | 11:86847703_CT_C | 11 | 86847703 | C | CT | 0.668145 | -0.0821794 | 0.0143159 | 9.50E-09 | 0.999978 | intron_variant | TMEM135 |
| rs628447 | 11:86848358_G_T | 11 | 86848358 | T | G | 0.668145 | -0.0821791 | 0.0143159 | 9.50E-09 | 0.999977 | intron_variant | TMEM135 |
| rs543915 | 11:86848478_T_C | 11 | 86848478 | C | T | 0.668145 | -0.0821791 | 0.0143159 | 9.50E-09 | 0.999977 | intron_variant | TMEM135 |
| rs630249 | 11:86848756_G_A | 11 | 86848756 | A | G | 0.668145 | -0.0821791 | 0.0143159 | 9.50E-09 | 0.999977 | intron_variant | TMEM135 |
| rs538494 | 11:86849091_G_A | 11 | 86849091 | A | G | 0.668145 | -0.0821791 | 0.0143159 | 9.50E-09 | 0.999975 | intron_variant | TMEM135 |
| rs535515 | 11:86849451_T_C | 11 | 86849451 | C | T | 0.668145 | -0.0821791 | 0.0143159 | 9.50E-09 | 0.999975 | intron_variant | TMEM135 |
| rs644083 | 11:86849633_T_A | 11 | 86849633 | A | T | 0.668145 | -0.0821791 | 0.0143159 | 9.50E-09 | 0.999975 | intron_variant | TMEM135 |
| rs644457 | 11:86849686_A_G | 11 | 86849686 | G | A | 0.668145 | -0.0821791 | 0.0143159 | 9.50E-09 | 0.999975 | intron_variant | TMEM135 |
| rs483373 | 11:86850483_C_T | 11 | 86850483 | T | C | 0.668145 | -0.0821791 | 0.0143159 | 9.50E-09 | 0.999974 | intron_variant | TMEM135 |
| rs2247537 | 11:86850705_G_A | 11 | 86850705 | A | G | 0.668145 | -0.0821791 | 0.0143159 | 9.50E-09 | 0.999974 | intron_variant | TMEM135 |
| rs2512350 | 11:86850736_A_G | 11 | 86850736 | G | A | 0.668145 | -0.0821791 | 0.0143159 | 9.50E-09 | 0.999974 | intron_variant | TMEM135 |
| rs672194 | 11:86851267_G_A | 11 | 86851267 | A | G | 0.668145 | -0.0821803 | 0.0143159 | 9.50E-09 | 0.999973 | intron_variant | TMEM135 |
| rs672269 | 11:86851316_A_G | 11 | 86851316 | G | A | 0.66791 | -0.08219 | 0.0143182 | 9.50E-09 | 0.999155 | intron_variant | TMEM135 |
| rs565869 | 11:86886904_A_C | 11 | 86886904 | C | A | 0.667604 | -0.0821921 | 0.0143197 | 9.60E-09 | 0.999473 | intron_variant | TMEM135 |
| rs487678 | 11:86847185_T_C | 11 | 86847185 | C | T | 0.668097 | -0.0821474 | 0.0143172 | 9.70E-09 | 0.999769 | intron_variant | TMEM135 |
| rs606284 | 11:86860244_C_T | 11 | 86860244 | T | C | 0.667496 | -0.0821383 | 0.0143151 | 9.70E-09 | 0.999608 | intron_variant | TMEM135 |
| rs531500 | 11:86868727_G_T | 11 | 86868727 | T | G | 0.667825 | -0.0821376 | 0.0143189 | 9.80E-09 | 0.999622 | intron_variant | TMEM135 |
| rs5793242 | 11:86868732_GA_G | 11 | 86868732 | G | GA | 0.667825 | -0.0821376 | 0.0143189 | 9.80E-09 | 0.999622 | intron_variant | TMEM135 |
| rs5028348 | 11:86868742_A_G | 11 | 86868742 | G | A | 0.667825 | -0.0821376 | 0.0143189 | 9.80E-09 | 0.999622 | intron_variant | TMEM135 |
| rs7908619 | 10:54419140_A_G | 10 | 54419140 | G | A | 0.253617 | -0.089713 | 0.0157215 | 1.00E-08 | 0.979041 | intragenic_variant | RP11-556E13.1 |
| rs11234985 | 11:86842411_C_G | 11 | 86842411 | G | C | 0.668071 | -0.0820351 | 0.0143172 | 1.00E-08 | 0.999691 | intron_variant | TMEM135 |
| rs496671 | 11:86843374_A_G | 11 | 86843374 | G | A | 0.668093 | -0.0820365 | 0.0143173 | 1.00E-08 | 0.999847 | intron_variant | TMEM135 |
| rs625554 | 11:86855690_C_T | 11 | 86855690 | T | C | 0.667965 | -0.0820258 | 0.0143208 | 1.00E-08 | 0.999496 | intron_variant | TMEM135 |
| rs35881264;rs566036019 | 11:86856730_A_AT | 11 | 86856730 | AT | A | 0.667821 | -0.0820323 | 0.0143183 | 1.00E-08 | 0.999183 | intron_variant | TMEM135 |
| rs549932 | 11:86872908_C_T | 11 | 86872908 | C | T | 0.668334 | -0.0820891 | 0.0143227 | 1.00E-08 | 1 | intron_variant | TMEM135 |
| rs634277 | 11:86887931_A_G | 11 | 86887931 | G | A | 0.667569 | -0.0820854 | 0.0143184 | 1.00E-08 | 0.999608 | intron_variant | TMEM135 |
| rs7920998 | 10:54421272_T_C | 10 | 54421272 | C | T | 0.252635 | -0.0897705 | 0.0157703 | 1.10E-08 | 0.980806 | intragenic_variant | RP11-556E13.1 |
| rs2512495 | 11:86826934_T_A | 11 | 86826934 | A | T | 0.671934 | -0.0821435 | 0.014361 | 1.10E-08 | 0.999884 | intron_variant | TMEM135 |

| | | | | | | | | | | | | |
|---|---|---|---|---|---|---|---|---|---|---|---|---|
| rs2512496 | 11:86826963_G_A | 11 | 86826963 | A | G | 0.671994 | -0.08207 | 0.0143599 | 1.10E-08 | 0.999954 | intron_variant | TMEM135 |
| rs5793237 | 11:86827617_T_TTGA | 11 | 86827617 | TTGA | T | 0.671994 | -0.0820694 | 0.0143599 | 1.10E-08 | 0.999961 | upstream_gene_variant | RP11-648O15.2 |
| rs2445560 | 11:86827825_C_G | 11 | 86827825 | G | C | 0.671994 | -0.0820694 | 0.0143599 | 1.10E-08 | 0.999961 | upstream_gene_variant | RP11-648O15.2 |
| rs2451065 | 11:86827946_C_T | 11 | 86827946 | T | C | 0.671994 | -0.0820702 | 0.0143599 | 1.10E-08 | 0.999962 | upstream_gene_variant | RP11-648O15.2 |
| rs2451064 | 11:86828153_G_A | 11 | 86828153 | A | G | 0.671903 | -0.0820026 | 0.014362 | 1.10E-08 | 0.999578 | upstream_gene_variant | RP11-648O15.2 |
| rs2451063 | 11:86828228_G_A | 11 | 86828228 | A | G | 0.671994 | -0.0820702 | 0.0143599 | 1.10E-08 | 0.999963 | upstream_gene_variant | RP11-648O15.2 |
| rs2451062 | 11:86828686_T_C | 11 | 86828686 | C | T | 0.671994 | -0.0820697 | 0.0143598 | 1.10E-08 | 0.999966 | upstream_gene_variant | RP11-648O15.2 |
| rs2445561 | 11:86828786_G_T | 11 | 86828786 | T | G | 0.671994 | -0.0820697 | 0.0143598 | 1.10E-08 | 0.999967 | upstream_gene_variant | RP11-648O15.2 |
| rs2451061 | 11:86830485_G_A | 11 | 86830485 | A | G | 0.671994 | -0.0820692 | 0.0143598 | 1.10E-08 | 0.999977 | upstream_gene_variant | RP11-648O15.2 |
| rs3018710 | 11:86830715_C_T | 11 | 86830715 | T | C | 0.671994 | -0.0820632 | 0.0143597 | 1.10E-08 | 0.99998 | upstream_gene_variant | RP11-648O15.2 |
| rs3018709 | 11:86830772_G_C | 11 | 86830772 | C | G | 0.672033 | -0.082073 | 0.014362 | 1.10E-08 | 0.999836 | upstream_gene_variant | RP11-648O15.2 |
| rs3103386 | 11:86830793_A_G | 11 | 86830793 | G | A | 0.671994 | -0.0820618 | 0.0143597 | 1.10E-08 | 0.999981 | upstream_gene_variant | RP11-648O15.2 |
| rs660246 | 11:86832576_A_T | 11 | 86832576 | T | A | 0.671513 | -0.0821669 | 0.014368 | 1.10E-08 | 0.998175 | intron_variant | TMEM135 |
| rs488227 | 11:86833101_G_A | 11 | 86833101 | A | G | 0.671994 | -0.082058 | 0.0143595 | 1.10E-08 | 1 | downstream_gene_variant | RP11-648O15.2 |
| rs1829359 | 11:86833562_T_G | 11 | 86833562 | G | T | 0.671757 | -0.0821323 | 0.0143641 | 1.10E-08 | 0.999264 | downstream_gene_variant | RP11-648O15.2 |
| rs1829360 | 11:86833565_T_C | 11 | 86833565 | C | T | 0.671757 | -0.0821323 | 0.0143641 | 1.10E-08 | 0.999264 | downstream_gene_variant | RP11-648O15.2 |
| rs518489 | 11:86834117_T_C | 11 | 86834117 | C | T | 0.671994 | -0.082058 | 0.0143595 | 1.10E-08 | 1 | downstream_gene_variant | RP11-648O15.2 |
| rs35903615;rs61541613 | 11:86834313_C_CCTT | 11 | 86834313 | CCTT | C | 0.671426 | -0.0822019 | 0.0143652 | 1.10E-08 | 0.998883 | downstream_gene_variant | RP11-648O15.2 |
| rs35728847 | 11:86834319_G_T | 11 | 86834319 | T | G | 0.671426 | -0.0822019 | 0.0143652 | 1.10E-08 | 0.998883 | downstream_gene_variant | RP11-648O15.2 |
| rs34976066;rs527530852 | 11:86834623_TTG_T | 11 | 86834623 | T | TTG | 0.671994 | -0.082058 | 0.0143595 | 1.10E-08 | 1 | downstream_gene_variant | RP11-648O15.2 |
| rs2512343 | 11:86835260_G_T | 11 | 86835260 | T | G | 0.671994 | -0.082058 | 0.0143595 | 1.10E-08 | 1 | downstream_gene_variant | RP11-648O15.2 |
| rs2512344 | 11:86835374_G_A | 11 | 86835374 | G | A | 0.671994 | -0.082058 | 0.0143595 | 1.10E-08 | 1 | downstream_gene_variant | RP11-648O15.2 |
| rs692857 | 11:86835475_G_A | 11 | 86835475 | A | G | 0.671834 | -0.0821479 | 0.0143627 | 1.10E-08 | 0.999271 | downstream_gene_variant | RP11-648O15.2 |
| rs511694 | 11:86872365_G_A | 11 | 86872365 | A | G | 0.667587 | -0.081901 | 0.014327 | 1.10E-08 | 0.998343 | intron_variant | TMEM135 |
| rs3133320 | 11:86830909_T_C | 11 | 86830909 | C | T | 0.671657 | -0.0819094 | 0.0143661 | 1.20E-08 | 0.998704 | upstream_gene_variant | RP11-648O15.2 |
| rs1829362 | 11:86833728_T_C | 11 | 86833728 | C | T | 0.67187 | -0.0819586 | 0.0143614 | 1.20E-08 | 0.9997 | downstream_gene_variant | RP11-648O15.2 |
| rs2512342 | 11:86835175_T_C | 11 | 86835175 | T | C | 0.67204 | -0.0818611 | 0.014362 | 1.20E-08 | 1 | downstream_gene_variant | RP11-648O15.2 |
| rs2512338 | 11:86840718_A_G | 11 | 86840718 | G | A | 0.667453 | -0.0817511 | 0.0143286 | 1.20E-08 | 0.996357 | intron_variant | TMEM135 |
| rs643145 | 11:86849397_C_T | 11 | 86849397 | T | C | 0.667504 | -0.0817612 | 0.0143259 | 1.20E-08 | 0.997878 | intron_variant | TMEM135 |
| rs643175 | 11:86849413_T_G | 11 | 86849413 | G | T | 0.667504 | -0.0817612 | 0.0143259 | 1.20E-08 | 0.997878 | intron_variant | TMEM135 |
| rs111978223;rs55697853 | 11:86853864_C_CTCT | 11 | 86853864 | CTCT | C | 0.671855 | -0.0821474 | 0.0144017 | 1.20E-08 | 0.995313 | intron_variant | TMEM135 |
| rs1373004 | 10:54427825_G_T | 10 | 54427825 | G | T | 0.741708 | 0.0877053 | 0.0155184 | 1.30E-08 | 1 | intragenic_variant | RP11-556E13.1 |
| rs597344 | 11:86835679_A_T | 11 | 86835679 | T | A | 0.671608 | -0.0817418 | 0.0143589 | 1.30E-08 | 0.999667 | downstream_gene_variant | RP11-648O15.2 |
| rs12256387 | 10:54422872_C_A | 10 | 54422872 | A | C | 0.258233 | -0.0874016 | 0.0154825 | 1.40E-08 | 0.999701 | intragenic_variant | RP11-556E13.1 |
| rs7895388 | 10:54423387_A_G | 10 | 54423387 | G | A | 0.258247 | -0.0873812 | 0.0154809 | 1.40E-08 | 0.999749 | intragenic_variant | RP11-556E13.1 |
| rs7898709 | 10:54423398_T_G | 10 | 54423398 | G | T | 0.258247 | -0.0873812 | 0.0154809 | 1.40E-08 | 0.999749 | intragenic_variant | RP11-556E13.1 |
| rs12258451 | 10:54423853_C_G | 10 | 54423853 | G | C | 0.258235 | -0.0874182 | 0.0154813 | 1.40E-08 | 0.999662 | intragenic_variant | RP11-556E13.1 |
| rs12258467 | 10:54423900_C_T | 10 | 54423900 | T | C | 0.258182 | -0.0874222 | 0.0154819 | 1.40E-08 | 0.99964 | intragenic_variant | RP11-556E13.1 |
| rs986104929 | 11:86827682_CACACACAG_<DEL> | 11 | 86827682 | <DEL> | CACACACAG | 0.6369 | -0.0812597 | 0.0143428 | 1.40E-08 | 0.954583 | upstream_gene_variant | RP11-648O15.2 |
| rs11424292 | 11:86848308_TA_TAA | 11 | 86848308 | TAA | TA | 0.666626 | -0.0813792 | 0.0143345 | 1.40E-08 | 0.996158 | intron_variant | TMEM135 |
| rs12266233 | 10:54424184_A_G | 10 | 54424184 | G | A | 0.258294 | -0.0872646 | 0.0154805 | 1.50E-08 | 0.999761 | intragenic_variant | RP11-556E13.1 |
| rs11003015 | 10:54427337_T_G | 10 | 54427337 | G | T | 0.258294 | -0.087167 | 0.0154777 | 1.50E-08 | 0.999931 | intragenic_variant | RP11-556E13.1 |
| rs397751547;rs5784941 | 10:54427519_AT_A | 10 | 54427519 | A | AT | 0.741706 | 0.0871591 | 0.0154776 | 1.50E-08 | 0.999939 | intragenic_variant | RP11-556E13.1 |
| rs11003037 | 10:54428956_G_A | 10 | 54428956 | A | G | 0.258489 | -0.087235 | 0.0154827 | 1.50E-08 | 0.99935 | intragenic_variant | RP11-556E13.1 |
| rs483548 | 11:86832604_T_G | 11 | 86832604 | G | T | 0.670921 | -0.0814244 | 0.0143767 | 1.50E-08 | 0.996799 | intron_variant | TMEM135 |
| rs597406 | 11:86835722_G_A | 11 | 86835722 | A | G | 0.670838 | -0.0812067 | 0.0143569 | 1.50E-08 | 0.99942 | downstream_gene_variant | RP11-648O15.2 |
| rs7099953 | 10:54426489_G_T | 10 | 54426489 | T | G | 0.258246 | -0.08704 | 0.0154854 | 1.60E-08 | 0.999928 | intragenic_variant | RP11-556E13.1 |
| rs201765865;rs149856990;rs771834625;rs869233649 | 10:54429002_AAAATAAATAAAT_A | 10 | 54429002 | A | AAAATAAATAAAT | 0.25801 | -0.0868773 | 0.0154833 | 1.70E-08 | 0.999246 | intragenic_variant | RP11-556E13.1 |
| rs5028563 | 10:54429526_T_A | 10 | 54429526 | A | T | 0.741814 | 0.0868713 | 0.0154808 | 1.70E-08 | 0.999543 | intragenic_variant | RP11-556E13.1 |
| rs11003047 | 10:54434511_T_G | 10 | 54434511 | G | T | 0.258129 | -0.0868994 | 0.0154935 | 1.70E-08 | 0.99943 | intragenic_variant | RP11-556E13.1 |
| rs11003048 | 10:54434529_C_G | 10 | 54434529 | G | C | 0.257916 | -0.0869922 | 0.0155038 | 1.70E-08 | 0.998505 | intragenic_variant | RP11-556E13.1 |
| rs74133652 | 10:54424356_G_C | 10 | 54424356 | C | G | 0.258153 | -0.0867479 | 0.0154802 | 1.80E-08 | 0.999768 | intragenic_variant | RP11-556E13.1 |
| rs73333526 | 10:54424528_A_G | 10 | 54424528 | G | A | 0.258153 | -0.0867446 | 0.01548 | 1.80E-08 | 0.999774 | intragenic_variant | RP11-556E13.1 |
| rs11002987 | 10:54425113_G_A | 10 | 54425113 | A | G | 0.258153 | -0.0867272 | 0.0154795 | 1.80E-08 | 0.999802 | intragenic_variant | RP11-556E13.1 |
| rs11002989 | 10:54425167_C_T | 10 | 54425167 | T | C | 0.258153 | -0.0867261 | 0.0154794 | 1.80E-08 | 0.999806 | intragenic_variant | RP11-556E13.1 |
| rs7095115 | 10:54425442_G_A | 10 | 54425442 | A | G | 0.258153 | -0.0867208 | 0.0154791 | 1.80E-08 | 0.999821 | intragenic_variant | RP11-556E13.1 |
| rs7095383 | 10:54425601_G_T | 10 | 54425601 | T | G | 0.258124 | -0.086721 | 0.0154807 | 1.80E-08 | 0.999664 | intragenic_variant | RP11-556E13.1 |
| rs7095538 | 10:54425675_G_A | 10 | 54425675 | A | G | 0.258153 | -0.086714 | 0.0154789 | 1.80E-08 | 0.999837 | intragenic_variant | RP11-556E13.1 |
| rs11003038 | 10:54433560_T_C | 10 | 54433560 | C | T | 0.258239 | -0.0867807 | 0.0154886 | 1.80E-08 | 0.999508 | intragenic_variant | RP11-556E13.1 |
| rs11003040 | 10:54433608_G_A | 10 | 54433608 | A | G | 0.258239 | -0.0867807 | 0.0154886 | 1.80E-08 | 0.999508 | intragenic_variant | RP11-556E13.1 |
| rs11003044 | 10:54433988_A_G | 10 | 54433988 | G | A | 0.258239 | -0.0867807 | 0.0154886 | 1.80E-08 | 0.999508 | intragenic_variant | RP11-556E13.1 |
| rs11003045 | 10:54433993_A_G | 10 | 54433993 | G | A | 0.258239 | -0.0867807 | 0.0154886 | 1.80E-08 | 0.999508 | intragenic_variant | RP11-556E13.1 |
| rs11003046 | 10:54434345_C_T | 10 | 54434345 | T | C | 0.241281 | -0.0909111 | 0.0162731 | 1.80E-08 | 0.947945 | intragenic_variant | RP11-556E13.1 |
| rs7067581 | 10:54434926_A_G | 10 | 54434926 | G | A | 0.258239 | -0.0867833 | 0.0154886 | 1.80E-08 | 0.999506 | intragenic_variant | RP11-556E13.1 |
| rs7071681 | 10:54435158_T_C | 10 | 54435158 | C | T | 0.258239 | -0.0867851 | 0.0154886 | 1.80E-08 | 0.999506 | intragenic_variant | RP11-556E13.1 |
| rs10824766 | 10:54428297_C_T | 10 | 54428297 | T | C | 0.258154 | -0.0866373 | 0.0154774 | 1.90E-08 | 0.999944 | intragenic_variant | RP11-556E13.1 |
| rs11003035 | 10:54428726_G_A | 10 | 54428726 | A | G | 0.258092 | -0.0866197 | 0.0154805 | 1.90E-08 | 0.999639 | intragenic_variant | RP11-556E13.1 |
| rs10762869 | 10:54429118_C_T | 10 | 54429118 | T | C | 0.741904 | 0.0866679 | 0.0154797 | 1.90E-08 | 0.999701 | intragenic_variant | RP11-556E13.1 |
| rs11003043 | 10:54433914_A_G | 10 | 54433914 | G | A | 0.258182 | -0.0866928 | 0.015489 | 1.90E-08 | 0.999463 | intragenic_variant | RP11-556E13.1 |
| rs6480949 | 10:54435422_C_T | 10 | 54435422 | T | C | 0.256944 | -0.0867808 | 0.0155237 | 1.90E-08 | 0.998985 | intragenic_variant | RP11-556E13.1 |
| rs7070913 | 10:54420223_G_A | 10 | 54420223 | A | G | 0.257375 | -0.0865374 | 0.0155053 | 2.10E-08 | 0.998942 | intragenic_variant | RP11-556E13.1 |
| rs35505248 | 11:86830927_T_A | 11 | 86830927 | A | T | 0.670363 | -0.0807281 | 0.0143998 | 2.10E-08 | 0.992748 | upstream_gene_variant | RP11-648O15.2 |
| rs2707518 | 7:120954908_G_T | 7 | 120954908 | T | G | 0.458901 | 0.0771341 | 0.0136335 | 2.20E-08 | 0.98167 | intergenic_region | CPED1:WNT16 |
| rs12416665 | 10:54430631_T_C | 10 | 54430631 | C | T | 0.258784 | -0.086199 | 0.0154789 | 2.20E-08 | 0.99939 | intragenic_variant | RP11-556E13.1 |
| rs1443131 | 10:54432209_T_A | 10 | 54432209 | T | A | 0.741199 | 0.0861603 | 0.0154742 | 2.20E-08 | 1 | intragenic_variant | RP11-556E13.1 |
| rs7900044 | 10:54418347_T_A | 10 | 54418347 | A | T | 0.258388 | -0.0863755 | 0.0155131 | 2.30E-08 | 0.994866 | intragenic_variant | RP11-556E13.1 |
| rs7924102 | 10:54419383_C_G | 10 | 54419383 | G | C | 0.254798 | -0.0871119 | 0.0156657 | 2.30E-08 | 0.98717 | intragenic_variant | RP11-556E13.1 |
| rs10824760 | 10:54425325_C_T | 10 | 54425325 | T | C | 0.741735 | 0.0861496 | 0.0154804 | 2.30E-08 | 0.999517 | intragenic_variant | RP11-556E13.1 |
| rs12251299 | 10:54415687_C_T | 10 | 54415687 | T | C | 0.257489 | -0.0866295 | 0.0155783 | 2.40E-08 | 0.991404 | intragenic_variant | RP11-556E13.1 |
| rs6480933 | 10:54417411_T_C | 10 | 54417411 | C | T | 0.741797 | 0.0862637 | 0.0155258 | 2.40E-08 | 0.994731 | intragenic_variant | RP11-556E13.1 |
| rs12250150 | 10:54419939_C_T | 10 | 54419939 | T | C | 0.257993 | -0.0860972 | 0.0154996 | 2.40E-08 | 0.998327 | intragenic_variant | RP11-556E13.1 |
| rs11002958 | 10:54420086_G_C | 10 | 54420086 | C | G | 0.257969 | -0.0861525 | 0.0154933 | 2.40E-08 | 0.999173 | intragenic_variant | RP11-556E13.1 |
| rs12268596 | 10:54421876_G_A | 10 | 54421876 | A | G | 0.257921 | -0.0861319 | 0.0155179 | 2.40E-08 | 1 | intragenic_variant | RP11-556E13.1 |
| rs12262178 | 10:54431185_C_T | 10 | 54431185 | T | C | 0.258687 | -0.0859046 | 0.0154752 | 2.40E-08 | 0.999811 | intragenic_variant | RP11-556E13.1 |
| rs12247120 | 10:54431210_G_T | 10 | 54431210 | T | G | 0.258687 | -0.0859057 | 0.0154752 | 2.40E-08 | 0.999814 | intragenic_variant | RP11-556E13.1 |
| rs12262251 | 10:54431330_C_T | 10 | 54431330 | T | C | 0.258686 | -0.0859084 | 0.0154752 | 2.40E-08 | 0.999815 | intragenic_variant | RP11-556E13.1 |
| rs1373003 | 10:54431543_A_G | 10 | 54431543 | G | A | 0.258733 | -0.0858013 | 0.0154753 | 2.50E-08 | 0.999813 | intragenic_variant | RP11-556E13.1 |
| rs7092106 | 10:54432970_A_G | 10 | 54432970 | G | A | 0.258078 | -0.085925 | 0.0154876 | 2.50E-08 | 0.999603 | intragenic_variant | RP11-556E13.1 |
| rs11002954 | 10:54418743_G_T | 10 | 54418743 | T | G | 0.257921 | -0.086122 | 0.0155265 | 2.60E-08 | 0.994666 | intragenic_variant | RP11-556E13.1 |
| rs7923017 | 10:54419315_G_C | 10 | 54419315 | C | G | 0.257897 | -0.0859884 | 0.0155231 | 2.70E-08 | 0.994343 | intragenic_variant | RP11-556E13.1 |
| rs11003049 | 10:54437506_G_A | 10 | 54437506 | A | G | 0.257149 | -0.0858345 | 0.0155221 | 2.70E-08 | 0.998141 | intragenic_variant | RP11-556E13.1 |
| rs11003050 | 10:54437606_G_A | 10 | 54437606 | A | G | 0.256941 | -0.0857826 | 0.0155185 | 2.80E-08 | 0.998635 | intragenic_variant | RP11-556E13.1 |
| rs74611091 | 10:54437068_TG_T | 10 | 54437068 | T | TG | 0.257147 | -0.0855954 | 0.0155135 | 2.90E-08 | 0.998867 | intragenic_variant | RP11-556E13.1 |
| rs11003051 | 10:54437901_G_T | 10 | 54437901 | T | G | 0.257149 | -0.0856016 | 0.0155135 | 2.90E-08 | 0.998856 | intragenic_variant | RP11-556E13.1 |
| rs7097953 | 10:54438292_G_A | 10 | 54438292 | A | G | 0.257148 | -0.085602 | 0.0155136 | 2.90E-08 | 0.998853 | intragenic_variant | RP11-556E13.1 |
| rs473364 | 11:86860984_C_T | 11 | 86860984 | T | C | 0.617168 | -0.0767376 | 0.0139121 | 3.20E-08 | 0.994176 | intron_variant | TMEM135 |
| rs138578420;rs575889421;rs553815040 | 10:54431741_CAAAACAAAAACA_CAAAACA | 10 | 54431741 | CAAAACA | CAAAACAAAAACA | 0.285256 | -0.0825397 | 0.0150438 | 3.50E-08 | 0.985738 | intragenic_variant | RP11-556E13.1 |
| rs2512494 | 11:86826749_T_A | 11 | 86826749 | A | T | 0.678106 | -0.0793056 | 0.0144577 | 4.40E-08 | 0.998646 | intron_variant | TMEM135 |
| rs607616 | 11:86860547_T_C | 11 | 86860547 | C | T | 0.738233 | -0.0834891 | 0.0153351 | 5.10E-08 | 0.999946 | intron_variant | TMEM135 |
| rs573889 | 11:86863077_A_G | 11 | 86863077 | G | A | 0.738241 | -0.0834801 | 0.0153357 | 5.10E-08 | 0.999912 | intron_variant | TMEM135 |
| rs565938 | 11:86864416_G_A | 11 | 86864416 | A | G | 0.738241 | -0.0834801 | 0.0153357 | 5.10E-08 | 0.999912 | intron_variant | TMEM135 |
| rs525139 | 11:86869374_C_T | 11 | 86869374 | T | C | 0.738251 | -0.0834558 | 0.0153372 | 5.10E-08 | 0.999813 | intron_variant | TMEM135 |
| rs473206 | 11:86871694_A_G | 11 | 86871694 | G | A | 0.738259 | -0.083455 | 0.0153372 | 5.10E-08 | 0.99986 | intron_variant | TMEM135 |
| rs477062 | 11:86878190_C_A | 11 | 86878190 | A | C | 0.738261 | -0.0834656 | 0.0153374 | 5.10E-08 | 0.999843 | intron_variant | TMEM135 |
| rs623606 | 11:86878930_G_C | 11 | 86878930 | C | G | 0.738261 | -0.0834671 | 0.0153374 | 5.10E-08 | 0.999841 | intron_variant | TMEM135 |
| rs34430171 | 11:86862641_CT_C | 11 | 86862641 | C | CT | 0.738197 | -0.0833926 | 0.0153363 | 5.30E-08 | 0.999759 | intron_variant | TMEM135 |
| rs77126167 | 11:86873727_G_C | 11 | 86873727 | C | G | 0.61423 | -0.0750758 | 0.01385 | 5.30E-08 | 0.998666 | intron_variant | TMEM135 |
| rs7902708 | 10:54439728_G_C | 10 | 54439728 | C | G | 0.254854 | -0.0844774 | 0.0156023 | 5.50E-08 | 0.994867 | intragenic_variant | RP11-556E13.1 |
| rs7919753 | 10:54439699_A_G | 10 | 54439699 | G | A | 0.255222 | -0.0842814 | 0.0155838 | 5.60E-08 | 0.996941 | intragenic_variant | RP11-556E13.1 |
| rs583334 | 11:86834865_A_C | 11 | 86834865 | C | A | 0.663631 | -0.0776826 | 0.0143466 | 5.80E-08 | 0.988597 | downstream_gene_variant | RP11-648O15.2 |
| rs505429 | 11:86886650_A_G | 11 | 86886650 | G | A | 0.618373 | -0.0752102 | 0.0139097 | 5.80E-08 | 0.999186 | intron_variant | TMEM135 |
| rs1042006841 | 11:86870631_A_ACG | 11 | 86870631 | ACG | A | 0.575055 | -0.0751259 | 0.0139798 | 6.60E-08 | 0.947489 | intron_variant | TMEM135 |
| rs605313 | 11:86885081_C_G | 11 | 86885081 | G | C | 0.612516 | -0.0744447 | 0.0138313 | 6.80E-08 | 0.999589 | intron_variant | TMEM135 |
| rs476832 | 11:86860663_A_G | 11 | 86860663 | A | G | 0.737956 | -0.0823784 | 0.0153139 | 7.30E-08 | 1 | intron_variant | TMEM135 |
| rs522211 | 11:86869565_A_G | 11 | 86869565 | G | A | 0.733109 | -0.0828029 | 0.015388 | 7.40E-08 | 0.981176 | intron_variant | TMEM135 |
| rs563133 | 11:86836450_A_T | 11 | 86836450 | T | A | 0.612925 | -0.0741565 | 0.0138302 | 7.50E-08 | 0.999813 | downstream_gene_variant | RP11-648O15.2 |
| rs503939 | 11:86859980_G_C | 11 | 86859980 | C | G | 0.61264 | -0.0741276 | 0.0138303 | 7.60E-08 | 0.999653 | intron_variant | TMEM135 |
| rs502144 | 11:86860178_A_T | 11 | 86860178 | T | A | 0.61264 | -0.0741278 | 0.0138303 | 7.60E-08 | 0.999655 | intron_variant | TMEM135 |
| rs500109 | 11:86860450_G_A | 11 | 86860450 | A | G | 0.61264 | -0.0741278 | 0.0138303 | 7.60E-08 | 0.999658 | intron_variant | TMEM135 |
| rs575992 | 11:86868037_T_G | 11 | 86868037 | G | T | 0.612693 | -0.0740687 | 0.0138305 | 7.80E-08 | 0.999739 | intron_variant | TMEM135 |
| rs496834 | 11:86870187_G_A | 11 | 86870187 | A | G | 0.612695 | -0.0740654 | 0.0138305 | 7.80E-08 | 0.999751 | intron_variant | TMEM135 |
| rs529564 | 11:86870367_A_G | 11 | 86870367 | G | A | 0.612695 | -0.0740654 | 0.0138305 | 7.80E-08 | 0.999751 | intron_variant | TMEM135 |
| rs521252 | 11:86852466_C_T | 11 | 86852466 | T | C | 0.612778 | -0.0739728 | 0.0138258 | 8.00E-08 | 0.999831 | intron_variant | TMEM135 |
| rs582415 | 11:86853015_G_C | 11 | 86853015 | C | G | 0.612773 | -0.0739799 | 0.0138262 | 8.00E-08 | 0.999781 | intron_variant | TMEM135 |
| rs481278 | 11:86841274_A_G | 11 | 86841274 | A | G | 0.612794 | -0.0739464 | 0.0138245 | 8.10E-08 | 1 | intron_variant | TMEM135 |
| rs534965 | 11:86841518_G_A | 11 | 86841518 | A | G | 0.612794 | -0.0739464 | 0.0138245 | 8.10E-08 | 1 | intron_variant | TMEM135 |
| rs503952 | 11:86842588_T_C | 11 | 86842588 | C | T | 0.612794 | -0.0739464 | 0.0138245 | 8.10E-08 | 0.999999 | intron_variant | TMEM135 |
| rs561012626;rs67409079;rs772508343;rs869059643;rs57192191;rs796565273 | 11:86841208_ATTTT_ATT | 11 | 86841208 | ATT | ATTTT | 0.612728 | -0.0739166 | 0.0138257 | 8.20E-08 | 0.999781 | intron_variant | TMEM135 |
| rs671781 | 11:86851221_C_T | 11 | 86851221 | T | C | 0.612706 | -0.073907 | 0.0138263 | 8.30E-08 | 0.999693 | intron_variant | TMEM135 |
| rs581174 | 11:86870715_C_T | 11 | 86870715 | T | C | 0.612419 | -0.073956 | 0.0138369 | 8.40E-08 | 0.998762 | intron_variant | TMEM135 |
| rs633211 | 11:86868178_C_T | 11 | 86868178 | T | C | 0.613385 | -0.073927 | 0.0138499 | 8.60E-08 | 0.997746 | intron_variant | TMEM135 |
| rs59532014;rs796305564 | 11:86868361_GT_G | 11 | 86868361 | G | GT | 0.612239 | -0.073829 | 0.0138337 | 8.60E-08 | 0.998289 | intron_variant | TMEM135 |
| rs150258235;rs57158448 | 11:86837639_A_AAT | 11 | 86837639 | AAT | A | 0.612529 | -0.0737426 | 0.013832 | 8.80E-08 | 0.999501 | intron_variant | TMEM135 |
| rs637418 | 11:86862648_C_T | 11 | 86862648 | T | C | 0.612502 | -0.0736817 | 0.0138328 | 9.20E-08 | 0.999293 | intron_variant | TMEM135 |
| rs2451052 | 11:86840665_C_T | 11 | 86840665 | T | C | 0.61319 | -0.0739801 | 0.0138905 | 9.30E-08 | 0.991205 | intron_variant | TMEM135 |
| rs11003046 | 10:54434345_C_CT | 10 | 54434345 | CT | C | 0.720829 | 0.0836691 | 0.0157257 | 9.50E-08 | 0.922461 | intragenic_variant | RP11-556E13.1 |
| rs533931 | 11:86859066_G_A | 11 | 86859066 | A | G | 0.524919 | -0.0723704 | 0.0136217 | 9.70E-08 | 0.979819 | intron_variant | TMEM135 |
| rs301574 | 11:86891259_A_G | 11 | 86891259 | G | A | 0.738535 | -0.0818687 | 0.015368 | 9.80E-08 | 0.996904 | intron_variant | TMEM135 |
| rs139522735;rs3046714 | 11:86833399_AAAT_A | 11 | 86833399 | A | AAAT | 0.607587 | -0.073959 | 0.0139069 | 9.90E-08 | 0.984519 | downstream_gene_variant | RP11-648O15.2 |
| rs5793241 | 11:86855182_TA_T | 11 | 86855182 | T | TA | 0.617029 | -0.0737802 | 0.0138922 | 1.00E-07 | 0.998771 | intron_variant | TMEM135 |
| rs676384 | 11:86833909_A_G | 11 | 86833909 | G | A | 0.616428 | -0.0731988 | 0.0138456 | 1.10E-07 | 0.999694 | downstream_gene_variant | RP11-648O15.2 |
| rs541293 | 11:86834258_C_G | 11 | 86834258 | G | C | 0.615929 | -0.073238 | 0.0138464 | 1.10E-07 | 0.99857 | downstream_gene_variant | RP11-648O15.2 |
| rs2511095 | 11:86903463_A_G | 11 | 86903463 | G | A | 0.737504 | -0.0816522 | 0.0153578 | 1.10E-07 | 0.996681 | intron_variant | TMEM135 |
| rs301576 | 11:86904896_T_G | 11 | 86904896 | G | T | 0.737285 | -0.0816906 | 0.015362 | 1.10E-07 | 0.995929 | intron_variant | TMEM135 |
| rs201765865;rs149856990;rs771834625;rs869233649 | 10:54429002_AAAATAAATAAAT_AAAATAAAT | 10 | 54429002 | AAAATAAAT | AAAATAAATAAAT | 0.723532 | 0.0806423 | 0.0153334 | 1.20E-07 | 0.976697 | intragenic_variant | RP11-556E13.1 |
| rs140985529 | 11:86827680_CACACACACAG_C | 11 | 86827680 | C | CACACACACAG | 0.622781 | -0.0737789 | 0.0139626 | 1.20E-07 | 0.987577 | upstream_gene_variant | RP11-648O15.2 |
| rs2445562 | 11:86829389_C_T | 11 | 86829389 | T | C | 0.616502 | -0.0730781 | 0.0138445 | 1.20E-07 | 0.999961 | upstream_gene_variant | RP11-648O15.2 |
| rs7395163 | 11:86764138_A_T | 11 | 86764138 | T | A | 0.649843 | -0.0752177 | 0.0142009 | 1.30E-07 | 0.989368 | intron_variant | TMEM135 |
| rs3103383 | 11:86831787_C_T | 11 | 86831787 | T | C | 0.616477 | -0.0727386 | 0.0138469 | 1.30E-07 | 0.999187 | upstream_gene_variant | RP11-648O15.2 |
| rs613715 | 11:86847430_T_C | 11 | 86847430 | C | T | 0.619522 | -0.0731626 | 0.0138517 | 1.30E-07 | 0.999556 | intron_variant | TMEM135 |
| rs301595 | 11:86916847_G_A | 11 | 86916847 | A | G | 0.735282 | -0.0810164 | 0.0153154 | 1.30E-07 | 0.997352 | intron_variant | TMEM135 |
| rs301593 | 11:86918850_C_G | 11 | 86918850 | G | C | 0.735281 | -0.0809964 | 0.0153147 | 1.30E-07 | 0.997663 | intron_variant | TMEM135 |
| rs301589 | 11:86924934_A_G | 11 | 86924934 | G | A | 0.264644 | 0.0810573 | 0.0153191 | 1.30E-07 | 0.99768 | intron_variant | TMEM135 |
| rs169972 | 11:86925053_G_C | 11 | 86925053 | C | G | 0.264644 | 0.0810573 | 0.0153191 | 1.30E-07 | 0.99768 | intron_variant | TMEM135 |
| rs673634 | 11:86833304_G_A | 11 | 86833304 | A | G | 0.615709 | -0.0726313 | 0.0138606 | 1.40E-07 | 0.997304 | downstream_gene_variant | RP11-648O15.2 |
| rs12576009 | 11:86840226_T_C | 11 | 86840226 | C | T | 0.613316 | -0.0727236 | 0.0138556 | 1.40E-07 | 0.99505 | intron_variant | TMEM135 |
| rs397964354;rs60730766 | 11:86869506_A_ATT | 11 | 86869506 | ATT | A | 0.594588 | -0.0741076 | 0.0140926 | 1.40E-07 | 0.946074 | intron_variant | TMEM135 |
| rs301575 | 11:86899288_G_A | 11 | 86899288 | A | G | 0.737174 | -0.0807149 | 0.0153544 | 1.50E-07 | 0.996482 | intron_variant | TMEM135 |
| rs34309213;rs398076109;rs796568876 | 11:86899568_AC_A | 11 | 86899568 | A | AC | 0.737025 | -0.0807843 | 0.0153623 | 1.50E-07 | 0.994924 | intron_variant | TMEM135 |
| rs145723159;rs200078866 | 1:22673892_AAAG_A | 1 | 22673892 | A | AAAG | 0.38348 | -0.0743104 | 0.0139831 | 1.60E-07 | 0.986578 | intergenic_region | RP11-415K20.1:RP11-415K20.2 |
| rs301596 | 11:86916490_T_A | 11 | 86916490 | A | T | 0.735625 | -0.080493 | 0.015326 | 1.60E-07 | 0.996785 | intron_variant | TMEM135 |
| rs62476338 | 7:120960271_G_C | 7 | 120960271 | C | G | 0.432291 | 0.0720499 | 0.013691 | 1.70E-07 | 0.993617 | intergenic_region | CPED1:WNT16 |
| rs921872 | 11:86724086_C_A | 11 | 86724086 | A | C | 0.307548 | 0.0764401 | 0.014597 | 1.70E-07 | 0.993376 | intergenic_region | RP11-736K20.6:RP11-736K20.1 |
| rs57322949 | 11:86854407_TC_T | 11 | 86854407 | T | TC | 0.593491 | -0.0727987 | 0.0139898 | 1.70E-07 | 0.960232 | intron_variant | TMEM135 |
| rs35493055 | 11:86830981_T_C | 11 | 86830981 | C | T | 0.610108 | -0.0721661 | 0.0138443 | 1.80E-07 | 0.99601 | upstream_gene_variant | RP11-648O15.2 |

| | | | | | | | | | | | | |
|---|---|---|---|---|---|---|---|---|---|---|---|---|
| rs142297718;rs34592561;rs763813915 | 11:86934891_A_AAAAG | 11 | 86934891 | AAAAG | A | 0.263266 | 0.0804122 | 0.0153557 | 1.80E-07 | 0.996908 | intron_variant | TMEM135 |
| rs4655051 | 1:22674007_G_A | 1 | 22674007 | A | G | 0.389518 | -0.073244 | 0.0138678 | 2.00E-07 | 0.999114 | intergenic_region | RP11-415K20.1:RP11-415K20.2 |
| rs1031584 | 11:86726523_T_C | 11 | 86726523 | C | T | 0.314281 | 0.075041 | 0.0144513 | 2.00E-07 | 0.998948 | intergenic_region | RP11-736K20.6:RP11-736K20.1 |
| rs3133315 | 11:86756205_T_G | 11 | 86756205 | G | T | 0.653802 | -0.0740658 | 0.0141848 | 2.00E-07 | 0.997505 | intron_variant | TMEM135 |
| rs1343987 | 1:22679309_C_T | 1 | 22679309 | T | C | 0.397617 | -0.0731988 | 0.0138701 | 2.20E-07 | 0.986955 | intergenic_region | RP11-415K20.1:RP11-415K20.2 |
| rs34946498 | 11:86829123_A_AT | 11 | 86829123 | AT | A | 0.625018 | -0.0719521 | 0.0139539 | 2.20E-07 | 0.996301 | upstream_gene_variant | RP11-648O15.2 |
| rs2512375 | 11:86752832_C_G | 11 | 86752832 | G | C | 0.653755 | -0.0736993 | 0.0141837 | 2.30E-07 | 0.997207 | intron_variant | TMEM135 |
| rs36029875 | 11:86830925_T_G | 11 | 86830925 | G | T | 0.624029 | -0.0719837 | 0.0139772 | 2.30E-07 | 0.987422 | upstream_gene_variant | RP11-648O15.2 |
| rs2512374 | 11:86753511_T_C | 11 | 86753511 | C | T | 0.653821 | -0.073573 | 0.014186 | 2.40E-07 | 0.997208 | intron_variant | TMEM135 |
| rs140743221;rs4014711 | 11:86855751_A_AGTGTGT | 11 | 86855751 | AGTGTGT | A | 0.558049 | -0.0720621 | 0.0139865 | 2.40E-07 | 0.950416 | intron_variant | TMEM135 |
| rs12048660 | 1:22672363_C_G | 1 | 22672363 | G | C | 0.389661 | -0.0726095 | 0.0138602 | 2.50E-07 | 1 | intergenic_region | RP11-415K20.1:RP11-415K20.2 |
| rs2374995 | 11:86779772_A_G | 11 | 86779772 | G | A | 0.654438 | -0.0734225 | 0.0142041 | 2.70E-07 | 0.997035 | intron_variant | TMEM135 |
| rs2512377 | 11:86752002_T_G | 11 | 86752002 | G | T | 0.653454 | -0.072931 | 0.0141669 | 2.80E-07 | 0.996922 | intron_variant | TMEM135 |
| rs2445536 | 11:86767388_G_A | 11 | 86767388 | G | A | 0.653696 | -0.0729866 | 0.0141748 | 2.90E-07 | 1 | intron_variant | TMEM135 |
| rs2513238 | 11:86781005_T_G | 11 | 86781005 | G | T | 0.654698 | -0.0731189 | 0.0141909 | 2.90E-07 | 0.999104 | intron_variant | TMEM135 |
| rs482288 | 11:86946470_A_G | 11 | 86946470 | G | A | 0.262472 | 0.0788644 | 0.0153478 | 3.00E-07 | 0.998497 | intron_variant | TMEM135 |
| rs301586 | 11:86929219_A_G | 11 | 86929219 | G | A | 0.260975 | 0.0791478 | 0.0153999 | 3.10E-07 | 0.99821 | intron_variant | TMEM135 |
| rs1545600 | 11:86780449_G_A | 11 | 86780449 | A | G | 0.654312 | -0.0729642 | 0.0142002 | 3.20E-07 | 0.99721 | intron_variant | TMEM135 |
| rs1531145 | 11:86781362_A_T | 11 | 86781362 | T | A | 0.654968 | -0.0727496 | 0.0141882 | 3.30E-07 | 0.999739 | intron_variant | TMEM135 |
| rs2513237 | 11:86780864_A_G | 11 | 86780864 | G | A | 0.654876 | -0.0726815 | 0.0141905 | 3.40E-07 | 0.999363 | intron_variant | TMEM135 |
| rs717723 | 11:86783049_G_C | 11 | 86783049 | C | G | 0.65456 | -0.0727252 | 0.0141955 | 3.40E-07 | 0.998385 | intron_variant | TMEM135 |
| rs717722 | 11:86783175_A_G | 11 | 86783175 | G | A | 0.655038 | -0.0726749 | 0.0141916 | 3.40E-07 | 0.999394 | intron_variant | TMEM135 |
| rs604960 | 11:86885033_A_C | 11 | 86885033 | C | A | 0.562575 | -0.0688514 | 0.0135927 | 3.40E-07 | 0.999253 | intron_variant | TMEM135 |
| rs2512398 | 11:86781071_C_T | 11 | 86781071 | T | C | 0.654483 | -0.0727138 | 0.0141967 | 3.50E-07 | 0.99793 | intron_variant | TMEM135 |
| rs10898622 | 11:86937482_T_C | 11 | 86937482 | C | T | 0.262413 | 0.0783256 | 0.0153441 | 3.50E-07 | 0.998952 | intron_variant | TMEM135 |
| rs2512376 | 11:86752388_A_G | 11 | 86752388 | G | A | 0.654079 | -0.0724387 | 0.0141854 | 3.60E-07 | 0.997323 | intron_variant | TMEM135 |
| rs2513211 | 11:86778437_A_G | 11 | 86778437 | G | A | 0.655448 | -0.072424 | 0.0141937 | 3.80E-07 | 0.999796 | intron_variant | TMEM135 |
| rs11234951 | 11:86784866_C_T | 11 | 86784866 | T | C | 0.655006 | -0.0723627 | 0.0141918 | 3.90E-07 | 0.999125 | intron_variant | TMEM135 |
| rs2513235 | 11:86786080_T_C | 11 | 86786080 | C | T | 0.65478 | -0.0723604 | 0.0141905 | 3.90E-07 | 0.999 | intron_variant | TMEM135 |
| rs301573 | 11:86889990_A_G | 11 | 86889990 | G | A | 0.563636 | -0.0685027 | 0.0136079 | 3.90E-07 | 0.998316 | intron_variant | TMEM135 |
| rs576528 | 11:86863357_G_T | 11 | 86863357 | T | G | 0.562175 | -0.0684299 | 0.0135877 | 4.00E-07 | 0.998827 | intron_variant | TMEM135 |
| rs398076108;rs60110548;rs796217581 | 11:86896551_GT_G | 11 | 86896551 | G | GT | 0.562233 | -0.0685539 | 0.0136081 | 4.00E-07 | 0.997586 | intron_variant | TMEM135 |
| rs476943 | 11:86867188_A_T | 11 | 86867188 | T | A | 0.562785 | -0.0683972 | 0.0135936 | 4.10E-07 | 0.999174 | intron_variant | TMEM135 |
| rs7930231 | 11:86894442_C_G | 11 | 86894442 | G | C | 0.56207 | -0.0684263 | 0.0136102 | 4.20E-07 | 0.997018 | intron_variant | TMEM135 |
| rs2445571 | 11:86773293_A_G | 11 | 86773293 | G | A | 0.653904 | -0.0721445 | 0.0142029 | 4.40E-07 | 0.996501 | intron_variant | TMEM135 |
| rs471471 | 11:86884115_A_G | 11 | 86884115 | G | A | 0.561664 | -0.0681977 | 0.0135864 | 4.40E-07 | 0.999445 | intron_variant | TMEM135 |
| rs2445534 | 11:86770266_A_G | 11 | 86770266 | G | A | 0.655468 | -0.0720529 | 0.0142036 | 4.50E-07 | 0.998596 | intron_variant | TMEM135 |
| rs3100849 | 11:86780293_T_C | 11 | 86780293 | C | T | 0.648478 | -0.0725212 | 0.0143186 | 4.50E-07 | 0.973955 | intron_variant | TMEM135 |
| rs556160 | 11:86883000_C_T | 11 | 86883000 | T | C | 0.561493 | -0.0682204 | 0.0135926 | 4.50E-07 | 0.998606 | intron_variant | TMEM135 |
| rs11234939 | 11:86773562_A_G | 11 | 86773562 | G | A | 0.650927 | -0.072164 | 0.0142346 | 4.60E-07 | 0.987319 | intron_variant | TMEM135 |
| rs2445572 | 11:86775628_T_C | 11 | 86775628 | C | T | 0.655586 | -0.0719192 | 0.0141936 | 4.60E-07 | 0.999906 | intron_variant | TMEM135 |
| rs2512360 | 11:86776397_A_G | 11 | 86776397 | G | A | 0.655586 | -0.0719004 | 0.0141942 | 4.60E-07 | 0.999842 | intron_variant | TMEM135 |
| rs965466 | 11:86777067_C_T | 11 | 86777067 | T | C | 0.655587 | -0.0718814 | 0.0141946 | 4.70E-07 | 0.999806 | intron_variant | TMEM135 |
| rs2512362 | 11:86777114_A_G | 11 | 86777114 | G | A | 0.655556 | -0.071863 | 0.0141953 | 4.70E-07 | 0.999673 | intron_variant | TMEM135 |
| rs3100851 | 11:86779589_A_C | 11 | 86779589 | C | A | 0.654635 | -0.0718619 | 0.014198 | 4.70E-07 | 0.997959 | intron_variant | TMEM135 |
| rs634343 | 11:86887886_G_T | 11 | 86887886 | T | G | 0.561418 | -0.0678025 | 0.0135852 | 5.10E-07 | 0.999902 | intron_variant | TMEM135 |
| rs4655048 | 1:22668771_C_T | 1 | 22668771 | T | C | 0.390788 | -0.0713332 | 0.0139464 | 5.20E-07 | 0.986764 | intergenic_region | RP11-415K20.1:RP11-415K20.2 |
| rs11314221 | 11:86744934_CG_C | 11 | 86744934 | C | CG | 0.347775 | 0.0709174 | 0.0141242 | 5.40E-07 | 0.999646 | upstream_gene_variant | TMEM135 |
| rs622921 | 11:86888168_A_G | 11 | 86888168 | A | G | 0.56133 | -0.0676968 | 0.0135862 | 5.40E-07 | 1 | intron_variant | TMEM135 |
| rs672224 | 11:86857850_A_G | 11 | 86857850 | G | A | 0.561131 | -0.0675185 | 0.0135931 | 5.60E-07 | 0.996945 | intron_variant | TMEM135 |
| rs2512503 | 11:86747785_T_A | 11 | 86747785 | A | T | 0.652029 | -0.0707793 | 0.0141227 | 5.70E-07 | 0.999127 | upstream_gene_variant | TMEM135 |
| rs214740 | 11:86927589_C_T | 11 | 86927589 | T | C | 0.258836 | 0.0774821 | 0.0154393 | 5.70E-07 | 0.997152 | intron_variant | TMEM135 |
| rs202015642 | 11:97459491_CTATCTATA_C | 11 | 97459491 | C | CTATCTATA | 0.12584 | 0.108673 | 0.0216608 | 5.70E-07 | 0.88301 | intergenic_region | RP11-882G5.1:RNA5SP347 |
| rs58350802;rs61643428 | 11:86822708_A_AAAAG | 11 | 86822708 | AAAAG | A | 0.613971 | -0.0716747 | 0.0142939 | 5.90E-07 | 0.939873 | intron_variant | TMEM135 |
| rs199619942 | 11:86780102_T_TTGGGAGGTCGAGGCAGA | 11 | 86780102 | TTGGGAGGTCGAGGCAGA | T | 0.652995 | -0.0709146 | 0.0142081 | 6.70E-07 | 0.995404 | intron_variant | TMEM135 |
| rs201987191 | 13:76986625_T_TGC | 13 | 76986625 | TGC | T | 0.0124314 | -0.307644 | 0.0620505 | 7.70E-07 | 0.979498 | intergenic_region | RN7SL571P:RP11-275D4.1 |
| rs199977904;rs571202663;rs201227415;rs202111775 | 11:86868116_GTT_GT | 11 | 86868116 | GT | GTT | 0.598436 | -0.0681805 | 0.013934 | 8.90E-07 | 0.9729 | intron_variant | TMEM135 |
| rs10671410;rs386374399;rs527683380 | 11:86849130_CTTTT_CTTTTTTT | 11 | 86849130 | CTTTTTTT | CTTTT | 0.59737 | -0.0690994 | 0.0142199 | 1.00E-06 | 0.931803 | intron_variant | TMEM135 |
| rs12233083 | 2:176929764_T_G | 2 | 176929764 | G | T | 0.279856 | 0.073558 | 0.0150409 | 1.10E-06 | 0.996769 | intergenic_region | KIAA1715:EVX2 |
| rs301577 | 11:86906491_G_A | 11 | 86906491 | A | G | 0.552447 | -0.0664694 | 0.0136911 | 1.10E-06 | 0.986559 | intron_variant | TMEM135 |
| rs113182670 | 11:86907817_C_CA | 11 | 86907817 | CA | C | 0.557314 | -0.0660096 | 0.0136213 | 1.10E-06 | 0.997155 | intron_variant | TMEM135 |
| rs74357740 | 2:176933551_C_G | 2 | 176933551 | G | C | 0.276769 | 0.073379 | 0.0151168 | 1.20E-06 | 0.994899 | intergenic_region | KIAA1715:EVX2 |
| rs516120 | 11:86846373_A_G | 11 | 86846373 | G | A | 0.569098 | -0.0653279 | 0.0135948 | 1.20E-06 | 0.998989 | intron_variant | TMEM135 |
| rs596912 | 11:86835594_T_C | 11 | 86835594 | C | T | 0.56812 | -0.065249 | 0.0135967 | 1.30E-06 | 0.998737 | downstream_gene_variant | RP11-648O15.2 |
| rs611538 | 11:86836533_T_G | 11 | 86836533 | G | T | 0.568273 | -0.0649559 | 0.0135967 | 1.40E-06 | 0.998758 | downstream_gene_variant | RP11-648O15.2 |
| rs35121629 | 11:86841163_TAA_AAA | 11 | 86841163 | AAA | TAA | 0.596351 | -0.0661685 | 0.0138087 | 1.40E-06 | 0.985291 | intron_variant | TMEM135 |
| rs28620001 | 11:86903029_C_CTT | 11 | 86903029 | CTT | C | 0.364869 | -0.0881571 | 0.0181689 | 1.40E-06 | 0.589081 | intron_variant | TMEM135 |
| rs755721733;rs34204350;rs768904197;rs202228550;rs571906738;rs761952192;rs869041217;rs869106091;rs869196555 | 11:86777640_CAAAAA_CA | 11 | 86777640 | CA | CAAAAA | 0.646758 | -0.0691717 | 0.0142919 | 1.50E-06 | 0.97399 | intron_variant | TMEM135 |
| rs367677158 | 14:31390275_TATACACACACACACACAC_TACACACACACACATACACACACACACACAC | 14 | 31390275 | TACACACACACACATACACACACACACACAC | TATACACACACACACACAC | 0.0732295 | 0.140208 | 0.028648 | 1.50E-06 | 0.805179 | intron_variant | STRN3 |
| rs74229190 | 1:22673895_G_<DEL> | 1 | 22673895 | <DEL> | G | 0.378883 | -0.0684912 | 0.0140737 | 1.60E-06 | 0.976546 | intergenic_region | RP11-415K20.1:RP11-415K20.2 |
| rs35126461;rs568712778;rs753310900;rs778283115;rs775469282 | 4:32011441_TAA_TA | 4 | 32011441 | TA | TAA | 0.41602 | -0.0733607 | 0.0154246 | 1.60E-06 | 0.804567 | intron_variant | RP11-734I18.1 |
| rs847152 | 2:176963127_C_A | 2 | 176963127 | A | C | 0.33666 | 0.0701294 | 0.0145642 | 1.70E-06 | 0.969541 | upstream_gene_variant | HOXD12 |
| rs28599706 | 8:6558522_G_C | 8 | 6558522 | C | G | 0.037545 | 0.170746 | 0.0358113 | 1.70E-06 | 0.977984 | intragenic_variant | CTD-2541M15.1 |
| rs1188669802 | 10:54419148_CTT_CT | 10 | 54419148 | CT | CTT | 0.702041 | 0.0792916 | 0.0166935 | 1.70E-06 | 0.789197 | intragenic_variant | RP11-556E13.1 |
| rs146145887 | 15:93327719_G_T | 15 | 93327719 | T | G | 0.0773242 | -0.123132 | 0.0260429 | 1.70E-06 | 0.95827 | upstream_gene_variant | CTD-2313J17.5 |
| rs35591739;rs796727700 | 11:86837712_TA_T | 11 | 86837712 | T | TA | 0.567346 | -0.0642772 | 0.01359 | 1.80E-06 | 0.998786 | intron_variant | TMEM135 |
| rs60128459 | 7:120965188_A_<DEL> | 7 | 120965188 | <DEL> | A | 0.0915783 | 0.126827 | 0.0267644 | 1.90E-06 | 0.767319 | upstream_gene_variant | WNT16 |
| rs398039283 | 17:38432772_CAAAAAAAAAAAAAA_CAAAAAAAAAAAAAAA | 17 | 38432772 | CAAAAAAAAAAAAAAA | CAAAAAAAAAAAAAA | 0.295197 | -0.0938493 | 0.0198414 | 1.90E-06 | 0.566946 | intron_variant | WIPF2 |
| rs58099920;rs147924485;rs34291756;rs58099920 | 2:137442867_CAAAA_CAAAAA | 2 | 137442867 | CAAAAA | CAAAA | 0.652078 | -0.071807 | 0.0153803 | 2.00E-06 | 0.853079 | intergenic_region | AC092786.1:THSD7B |
| rs28621953 | 8:6558148_C_G | 8 | 6558148 | G | C | 0.0374338 | 0.17031 | 0.0359034 | 2.00E-06 | 0.975913 | intragenic_variant | CTD-2541M15.1 |
| rs11234973 | 11:86830928_A_T | 11 | 86830928 | T | A | 0.16338 | 0.0888493 | 0.018494 | 2.00E-06 | 0.984425 | upstream_gene_variant | RP11-648O15.2 |
| rs11234982 | 11:86839600_A_G | 11 | 86839600 | G | A | 0.159345 | 0.0885098 | 0.0184799 | 2.00E-06 | 0.999573 | intron_variant | TMEM135 |
| rs622517 | 11:86843252_C_T | 11 | 86843252 | T | C | 0.567218 | -0.0640912 | 0.0135853 | 2.00E-06 | 0.998972 | intron_variant | TMEM135 |
| rs397724820;rs59105849 | 11:86843715_TAAC_T | 11 | 86843715 | T | TAAC | 0.567217 | -0.0640923 | 0.0135853 | 2.00E-06 | 0.998971 | intron_variant | TMEM135 |
| rs67967010 | 13:76992263_T_C | 13 | 76992263 | C | T | 0.0126717 | -0.291827 | 0.0610391 | 2.00E-06 | 0.993083 | intergenic_region | RN7SL571P:RP11-275D4.1 |
| rs1319443 | 11:86821502_TAA_TA | 11 | 86821502 | TA | TAA | 0.152748 | 0.0902609 | 0.0189245 | 2.10E-06 | 0.991472 | intron_variant | TMEM135 |
| rs2445535 | 11:86768330_T_A | 11 | 86768330 | A | T | 0.597888 | -0.0647783 | 0.0137073 | 2.30E-06 | 0.998813 | intron_variant | TMEM135 |
| rs11234977 | 11:86835749_A_G | 11 | 86835749 | G | A | 0.155922 | 0.0887929 | 0.0186762 | 2.30E-06 | 0.99893 | downstream_gene_variant | RP11-648O15.2 |
| rs35121629 | 11:86841163_TAA_TA | 11 | 86841163 | TA | TAA | 0.159976 | 0.0878699 | 0.0184765 | 2.30E-06 | 0.997341 | intron_variant | TMEM135 |
| rs5793233;rs111987507;rs543112646 | 11:86782716_AT_A | 11 | 86782716 | A | AT | 0.596315 | -0.0649599 | 0.0137396 | 2.40E-06 | 0.994202 | intron_variant | TMEM135 |
| rs35216554 | 11:86820870_C_CT | 11 | 86820870 | CT | C | 0.710995 | -0.0705845 | 0.0149159 | 2.40E-06 | 0.994081 | intron_variant | TMEM135 |
| rs301590 | 11:86921532_G_A | 11 | 86921532 | A | G | 0.440526 | 0.0639336 | 0.0136303 | 2.40E-06 | 0.997073 | intron_variant | TMEM135 |
| | 7:120965189_CACACACA_<DEL> | 7 | 120965189 | <DEL> | CACACACA | 0.0897151 | 0.126864 | 0.027051 | 2.50E-06 | 0.767948 | upstream_gene_variant | WNT16 |
| rs11820010 | 11:86754784_C_A | 11 | 86754784 | A | C | 0.150763 | 0.0902505 | 0.0190914 | 2.50E-06 | 0.98903 | intron_variant | TMEM135 |
| rs2513233 | 11:86767279_C_T | 11 | 86767279 | T | C | 0.598031 | -0.0645306 | 0.0137072 | 2.50E-06 | 0.999231 | intron_variant | TMEM135 |
| rs9667639 | 11:86829678_G_A | 11 | 86829678 | A | G | 0.155641 | 0.0884585 | 0.0186791 | 2.50E-06 | 0.999539 | upstream_gene_variant | RP11-648O15.2 |
| rs9666468 | 11:86829679_A_T | 11 | 86829679 | T | A | 0.155641 | 0.0884585 | 0.0186791 | 2.50E-06 | 0.999539 | upstream_gene_variant | RP11-648O15.2 |
| rs11234972 | 11:86830181_C_T | 11 | 86830181 | T | C | 0.155641 | 0.0884579 | 0.0186791 | 2.50E-06 | 0.999541 | upstream_gene_variant | RP11-648O15.2 |
| rs11234972 | 11:86850175_A_G | 11 | 86850175 | G | A | 0.158801 | 0.0877988 | 0.0185258 | 2.50E-06 | 0.998755 | intron_variant | TMEM135 |
| rs1188669802 | 10:54419148_CTT_CTTTTTTT | 10 | 54419148 | CTTTTTTT | CTT | 0.0843226 | -0.140958 | 0.0302761 | 2.60E-06 | 0.639369 | intragenic_variant | RP11-556E13.1 |
| rs11234976 | 11:86833654_C_A | 11 | 86833654 | A | C | 0.155639 | 0.0884533 | 0.0186789 | 2.60E-06 | 0.999561 | downstream_gene_variant | RP11-648O15.2 |
| rs10898603 | 11:86834507_A_T | 11 | 86834507 | T | A | 0.155639 | 0.0884545 | 0.0186789 | 2.60E-06 | 0.999562 | downstream_gene_variant | RP11-648O15.2 |
| rs1316342 | 1:22680517_A_C | 1 | 22680517 | C | A | 0.40172 | -0.0657732 | 0.0137696 | 2.70E-06 | 0.99573 | intergenic_region | RP11-415K20.1:RP11-415K20.2 |
| rs75955114 | 2:176955312_G_A | 2 | 176955312 | A | G | 0.278597 | 0.0710302 | 0.0150878 | 2.70E-06 | 0.992854 | upstream_gene_variant | HOXD13 |
| rs138043776 | 11:86286818_C_G | 11 | 86286818 | G | C | 0.0123772 | 0.29265 | 0.0622742 | 2.70E-06 | 0.96901 | intron_variant | RP11-317J19.1 |
| rs4943971 | 11:86764312_G_A | 11 | 86764312 | A | G | 0.597991 | -0.0643949 | 0.0137133 | 2.70E-06 | 0.998047 | intron_variant | TMEM135 |
| rs4531265 | 1:22701924_T_C | 1 | 22701924 | T | C | 0.567815 | -0.0638526 | 0.0136719 | 2.80E-06 | 1 | intergenic_region | RP11-415K20.2:ZBTB40 |
| rs79835123 | 2:176953563_G_A | 2 | 176953563 | A | G | 0.278917 | 0.0707436 | 0.0150425 | 2.80E-06 | 0.998773 | upstream_gene_variant | EVX2 |
| rs1319443 | 11:86821502_TAA_AAA | 11 | 86821502 | AAA | TAA | 0.721293 | -0.0709311 | 0.0150847 | 2.80E-06 | 0.990984 | intron_variant | TMEM135 |
| rs10898604 | 11:86841950_T_C | 11 | 86841950 | C | T | 0.160136 | 0.0870331 | 0.0184535 | 2.80E-06 | 0.999641 | intron_variant | TMEM135 |
| rs199500975 | 11:86882380_GTTT_GTT | 11 | 86882380 | GTT | GTTT | 0.15885 | 0.0875686 | 0.0185588 | 2.80E-06 | 0.994053 | intron_variant | TMEM135 |
| rs301591 | 11:86921301_G_A | 11 | 86921301 | A | G | 0.440368 | 0.0635419 | 0.0136312 | 2.80E-06 | 0.996744 | intron_variant | TMEM135 |
| rs190152553 | 22:32516850_C_A | 22 | 32516850 | A | C | 0.0135934 | -0.294438 | 0.0628825 | 2.80E-06 | 0.872298 | upstream_gene_variant | ADTB1L1 |
| rs2513228 | 11:86755539_A_G | 11 | 86755539 | G | A | 0.597592 | -0.0642551 | 0.0137171 | 2.90E-06 | 0.997421 | intron_variant | TMEM135 |
| rs658684 | 13:105274851_C_T | 13 | 105274851 | T | C | 0.462809 | 0.0657411 | 0.013917 | 2.90E-06 | 0.952589 | intergenic_region | RP11-357L1.1:RP11-217F17.1 |
| rs7555834 | 1:22710551_G_C | 1 | 22710551 | C | G | 0.401669 | -0.0652011 | 0.013769 | 3.00E-06 | 0.9949 | intergenic_region | RP11-415K20.2:ZBTB40 |
| rs10898600 | 11:86814547_T_G | 11 | 86814547 | G | T | 0.157691 | 0.0875858 | 0.0186204 | 3.00E-06 | 0.996386 | intron_variant | TMEM135 |
| rs11234989 | 11:86843062_C_T | 11 | 86843062 | T | C | 0.159088 | 0.0869125 | 0.0184995 | 3.00E-06 | 0.999531 | intron_variant | TMEM135 |
| rs10898605 | 11:86845497_A_C | 11 | 86845497 | C | A | 0.159088 | 0.0869134 | 0.0184995 | 3.00E-06 | 0.999532 | intron_variant | TMEM135 |
| rs10898606 | 11:86845501_C_T | 11 | 86845501 | T | C | 0.159088 | 0.0869134 | 0.0184995 | 3.00E-06 | 0.999532 | intron_variant | TMEM135 |
| rs10898607 | 11:86847306_T_A | 11 | 86847306 | A | T | 0.159088 | 0.0869151 | 0.0184995 | 3.00E-06 | 0.999532 | intron_variant | TMEM135 |
| rs10792909 | 11:86849115_A_C | 11 | 86849115 | C | A | 0.159088 | 0.0869151 | 0.0184995 | 3.00E-06 | 0.999531 | intron_variant | TMEM135 |
| rs10898608 | 11:86851496_C_G | 11 | 86851496 | G | C | 0.159088 | 0.0869158 | 0.0184996 | 3.00E-06 | 0.99953 | intron_variant | TMEM135 |
| rs10898609 | 11:86851499_A_G | 11 | 86851499 | G | A | 0.159088 | 0.0869158 | 0.0184996 | 3.00E-06 | 0.99953 | intron_variant | TMEM135 |
| rs10898610 | 11:86852648_G_A | 11 | 86852648 | A | G | 0.159087 | 0.0869216 | 0.0184999 | 3.00E-06 | 0.999504 | intron_variant | TMEM135 |
| rs10898613 | 11:86862889_C_T | 11 | 86862889 | T | C | 0.159054 | 0.0868943 | 0.0185033 | 3.00E-06 | 0.999317 | intron_variant | TMEM135 |
| rs11234996 | 11:86866895_G_A | 11 | 86866895 | A | G | 0.159054 | 0.0868965 | 0.0185034 | 3.00E-06 | 0.999315 | intron_variant | TMEM135 |
| rs17212655 | 11:86901237_C_T | 11 | 86901237 | T | C | 0.554782 | -0.0630494 | 0.0135989 | 3.00E-06 | 0.997189 | intron_variant | TMEM135 |
| rs11972702 | 7:38147661_C_T | 7 | 38147661 | T | C | 0.287823 | -0.0703625 | 0.0149733 | 3.10E-06 | 0.999245 | intergenic_region | EPDR1:STARD3NL |
| rs2512485 | 11:86757921_G_C | 11 | 86757921 | C | G | 0.597937 | -0.064078 | 0.013719 | 3.10E-06 | 0.997176 | intron_variant | TMEM135 |
| rs11234945 | 11:86780937_G_A | 11 | 86780937 | A | G | 0.155923 | 0.0878143 | 0.01871 | 3.10E-06 | 0.999536 | intron_variant | TMEM135 |
| rs11234946 | 11:86780958_T_G | 11 | 86780958 | G | T | 0.155923 | 0.0878143 | 0.01871 | 3.10E-06 | 0.999536 | intron_variant | TMEM135 |
| rs1968741 | 11:86781499_A_G | 11 | 86781499 | G | A | 0.155923 | 0.0878143 | 0.01871 | 3.10E-06 | 0.99953 | intron_variant | TMEM135 |
| rs1531144 | 11:86781808_A_G | 11 | 86781808 | G | A | 0.155926 | 0.0878202 | 0.01871 | 3.10E-06 | 0.999524 | intron_variant | TMEM135 |
| rs2512397 | 11:86783668_C_G | 11 | 86783668 | G | C | 0.60084 | -0.0641971 | 0.013746 | 3.10E-06 | 0.997978 | intron_variant | TMEM135 |
| rs10792908 | 11:86847176_T_A | 11 | 86847176 | A | T | 0.159137 | 0.0868481 | 0.0185009 | 3.10E-06 | 0.999185 | intron_variant | TMEM135 |
| rs60580053 | 4:82709501_GTATATATATGTATATGTATATATGTA_ATATATATATGTATATGTATATATGTA | 4 | 82709501 | ATATATATATGTATATGTATATATGTA | GTATATATATGTATATGTATATATGTA | 0.11478 | -0.13089 | 0.0274625 | 3.20E-06 | 0.598089 | intron_variant | RASGEF1B |
| rs3055240;rs397694502;rs797009344 | 7:38146624_AAT_A | 7 | 38146624 | A | AAT | 0.711018 | 0.0702017 | 0.0149615 | 3.20E-06 | 0.99935 | intergenic_region | EPDR1:STARD3NL |
| rs2512499 | 11:86753948_A_G | 11 | 86753948 | G | A | 0.597452 | -0.0639429 | 0.013715 | 3.20E-06 | 0.997345 | intron_variant | TMEM135 |
| rs35104865 | 11:86768858_T_TG | 11 | 86768858 | TG | T | 0.58941 | -0.0639938 | 0.013737 | 3.20E-06 | 0.986425 | intron_variant | TMEM135 |
| rs11234991 | 11:86855808_T_A | 11 | 86855808 | A | T | 0.159018 | 0.0866836 | 0.0185034 | 3.20E-06 | 0.999368 | intron_variant | TMEM135 |
| rs1042237573 | 2:123732214_C_T | 2 | 123732214 | T | C | 0.0103442 | -0.333428 | 0.0718938 | 3.30E-06 | 0.872654 | intergenic_region | AC018737.3:AC062020.2 |
| rs576205488 | 2:123775849_T_G | 2 | 123775849 | G | T | 0.0118601 | -0.307415 | 0.0660501 | 3.30E-06 | 0.890742 | intergenic_region | AC018737.3:AC062020.2 |
| rs10898586 | 11:86755466_A_T | 11 | 86755466 | T | A | 0.155534 | 0.0876273 | 0.0187494 | 3.30E-06 | 0.997965 | intron_variant | TMEM135 |
| rs11234965 | 11:86812299_C_T | 11 | 86812299 | T | C | 0.157157 | 0.0870892 | 0.0186144 | 3.30E-06 | 0.999314 | upstream_gene_variant | RP11-648O15.1 |
| rs11234966 | 11:86813178_A_T | 11 | 86813178 | T | A | 0.157155 | 0.0870877 | 0.0186143 | 3.30E-06 | 0.99933 | intron_variant | TMEM135 |
| rs67946471 | 11:86813716_T_C | 11 | 86813716 | C | T | 0.157154 | 0.0870911 | 0.0186143 | 3.30E-06 | 0.999341 | intron_variant | TMEM135 |
| rs11234968 | 11:86815453_G_T | 11 | 86815453 | T | G | 0.15715 | 0.087092 | 0.018614 | 3.30E-06 | 0.999375 | intron_variant | TMEM135 |
| rs11234969 | 11:86820258_C_G | 11 | 86820258 | G | C | 0.15714 | 0.0870788 | 0.0186132 | 3.30E-06 | 0.99947 | intron_variant | TMEM135 |
| rs11226844 | 11:97375179_A_G | 11 | 97375179 | G | A | 0.160178 | 0.0855558 | 0.0183494 | 3.30E-06 | 0.999115 | intergenic_region | RP11-882G5.1:RNA5SP347 |
| rs1403988 | 7:38109936_A_G | 7 | 38109936 | G | A | 0.712762 | 0.070039 | 0.0149729 | 3.40E-06 | 0.999966 | intergenic_region | EPDR1:STARD3NL |
| rs6956946 | 7:38110032_G_T | 7 | 38110032 | G | T | 0.712757 | 0.0700515 | 0.0149727 | 3.40E-06 | 1 | intergenic_region | EPDR1:STARD3NL |
| | 11:27254011_CA_CAA | 11 | 27254011 | CAA | CA | 0.30709 | 0.0734817 | 0.0157733 | 3.40E-06 | 0.847064 | intergenic_region | RP11-1L12.3:CCDC34 |
| rs10898587 | 11:86756502_C_T | 11 | 86756502 | T | C | 0.155536 | 0.0876167 | 0.0187492 | 3.40E-06 | 0.997973 | intron_variant | TMEM135 |
| rs11234941 | 11:86778113_C_T | 11 | 86778113 | T | C | 0.155503 | 0.0875651 | 0.0187344 | 3.40E-06 | 0.999561 | intron_variant | TMEM135 |
| rs11820668 | 11:86780044_C_G | 11 | 86780044 | G | C | 0.155827 | 0.0875586 | 0.0187193 | 3.40E-06 | 0.998977 | intron_variant | TMEM135 |
| rs7127557 | 11:86782274_T_A | 11 | 86782274 | A | T | 0.155851 | 0.0875084 | 0.0187133 | 3.40E-06 | 0.999389 | intron_variant | TMEM135 |
| rs73029923 | 2:176945667_T_C | 2 | 176945667 | C | T | 0.279229 | 0.0699788 | 0.0150426 | 3.50E-06 | 0.999053 | intron_variant | EVX2 |
| rs76494921 | 2:176946509_C_A | 2 | 176946509 | A | C | 0.279218 | 0.0699836 | 0.0150437 | 3.50E-06 | 0.998994 | intron_variant | EVX2 |
| rs6974574 | 7:38110073_A_T | 7 | 38110073 | T | A | 0.712758 | 0.0699908 | 0.0149727 | 3.50E-06 | 0.999978 | intergenic_region | EPDR1:STARD3NL |
| rs2512357 | 11:86775119_G_A | 11 | 86775119 | G | A | 0.599963 | -0.0637695 | 0.0137182 | 3.50E-06 | 1 | intron_variant | TMEM135 |
| rs11234958 | 11:86788716_T_G | 11 | 86788716 | G | T | 0.156705 | 0.0870874 | 0.0186606 | 3.50E-06 | 0.998194 | intron_variant | TMEM135 |
| rs35028039;rs796197116 | 11:86792726_GTT_GT | 11 | 86792726 | GT | GTT | 0.721796 | -0.0702595 | 0.015108 | 3.50E-06 | 0.99243 | intron_variant | TMEM135 |
| rs7117731 | 11:86824250_A_G | 11 | 86824250 | G | A | 0.156859 | 0.0870963 | 0.0186517 | 3.50E-06 | 0.996602 | intron_variant | TMEM135 |
| rs911232988 | 7:38099909_C_CT | 7 | 38099909 | CT | C | 0.712453 | 0.0699795 | 0.0149805 | 3.60E-06 | 0.998691 | intergenic_region | EPDR1:STARD3NL |

| | | | | | | | | | | | | |
|---|---|---|---|---|---|---|---|---|---|---|---|---|
| rs3502 | 8:6561058_G_A | 8 | 6561058 | A | G | 0.0377988 | 0.163089 | 0.0353381 | 3.60E-06 | 1 | upstream_gene_variant | AGPAT5 |
| rs3045939;rs72042001;rs773722597;rs869041270 | 11:86779850_CAAA_CA | 11 | 86779850 | CA | CAAA | 0.57996 | -0.0650299 | 0.0139884 | 3.60E-06 | 0.947686 | intron_variant | TMEM135 |
| rs1531146 | 11:86781279_G_A | 11 | 86781279 | A | G | 0.599441 | -0.0636856 | 0.0137162 | 3.60E-06 | 0.999774 | intron_variant | TMEM135 |
| rs1968742 | 11:86781426_C_T | 11 | 86781426 | T | C | 0.599441 | -0.0636856 | 0.0137162 | 3.60E-06 | 0.999773 | intron_variant | TMEM135 |
| rs2513239 | 11:86782728_T_G | 11 | 86782728 | G | T | 0.599437 | -0.0636729 | 0.0137164 | 3.60E-06 | 0.999739 | intron_variant | TMEM135 |
| rs12048810 | 1:22690547_G_A | 1 | 22690547 | A | G | 0.397949 | -0.0647405 | 0.0137445 | 3.70E-06 | 0.998655 | downstream_gene_variant | RP11-415K20.2 |
| rs112997284 | 2:176941272_G_A | 2 | 176941272 | A | G | 0.279229 | 0.0697606 | 0.0150356 | 3.70E-06 | 1 | downstream_gene_variant | EVX2 |
| rs143313442;rs3045965;rs35041310;rs771402309 | 11:86781679_AAACAAC_AAACAACAAC | 11 | 86781679 | AAACAACAAC | AAACAAC | 0.599094 | -0.0635936 | 0.0137206 | 3.70E-06 | 0.998987 | intron_variant | TMEM135 |
| rs12363760 | 11:86797750_G_C | 11 | 86797750 | C | G | 0.157154 | 0.0866861 | 0.0186169 | 3.70E-06 | 0.9991 | intron_variant | TMEM135 |
| rs113236993;rs398115596 | 11:86798786_G_GT | 11 | 86798786 | GT | G | 0.157154 | 0.0866869 | 0.0186169 | 3.70E-06 | 0.999101 | intron_variant | TMEM135 |
| rs10898594 | 11:86799055_C_T | 11 | 86799055 | T | C | 0.157154 | 0.0866869 | 0.0186169 | 3.70E-06 | 0.999101 | intron_variant | TMEM135 |
| rs10792906 | 11:86800527_G_A | 11 | 86800527 | A | G | 0.157154 | 0.0866916 | 0.0186169 | 3.70E-06 | 0.9991 | intron_variant | TMEM135 |
| rs145528154;rs200220386 | 13:30223920_T_TTA | 13 | 30223920 | TTA | T | 0.145331 | -0.0900508 | 0.0190931 | 3.70E-06 | 0.993613 | intergenic_region | SLC7A1:UBL3 |
| rs4655052 | 1:22688484_A_G | 1 | 22688484 | G | A | 0.398334 | -0.0646264 | 0.0137406 | 3.80E-06 | 0.999616 | downstream_gene_variant | RP11-415K20.2 |
| rs56009537 | 1:22708910_T_C | 1 | 22708910 | C | T | 0.403043 | -0.0643846 | 0.0137391 | 3.80E-06 | 0.99764 | intergenic_region | RP11-415K20.2:ZBTB40 |
| rs11888176 | 2:176951393_A_G | 2 | 176951393 | G | A | 0.279517 | 0.0697735 | 0.0150413 | 3.80E-06 | 0.999025 | upstream_gene_variant | EVX2 |
| rs10898584 | 11:86753088_C_T | 11 | 86753088 | T | C | 0.155661 | 0.0871723 | 0.0187467 | 3.80E-06 | 0.997901 | intron_variant | TMEM135 |
| rs10898593 | 11:86789285_G_A | 11 | 86789285 | A | G | 0.157045 | 0.0866427 | 0.0186371 | 3.80E-06 | 0.998247 | intron_variant | TMEM135 |
| rs56723130;rs796517410 | 11:86817151_CT_C | 11 | 86817151 | C | CT | 0.158134 | 0.0866228 | 0.0186066 | 3.80E-06 | 0.994981 | intron_variant | TMEM135 |
| rs55890170 | 13:30199555_T_C | 13 | 30199555 | C | T | 0.146294 | -0.089328 | 0.0189806 | 3.80E-06 | 0.998886 | intergenic_region | SLC7A1:UBL3 |
| rs35134156 | 15:77315432_A_G | 15 | 77315432 | G | A | 0.325038 | -0.0671937 | 0.0144822 | 3.80E-06 | 0.99185 | intron_variant | PSTPIP1 |
| rs72649450 | 1:22689344_G_T | 1 | 22689344 | T | G | 0.398369 | -0.0646003 | 0.0137386 | 3.90E-06 | 1 | downstream_gene_variant | RP11-415K20.2 |
| rs12048775 | 1:22690383_G_A | 1 | 22690383 | A | G | 0.397839 | -0.0646136 | 0.0137546 | 3.90E-06 | 0.99797 | downstream_gene_variant | RP11-415K20.2 |
| rs940347 | 7:38110425_A_G | 7 | 38110425 | G | A | 0.712766 | 0.0696582 | 0.0149725 | 3.90E-06 | 0.999909 | intergenic_region | EPDR1:STARD3NL |
| rs1524068 | 7:38110626_T_C | 7 | 38110626 | C | T | 0.712767 | 0.069644 | 0.0149725 | 3.90E-06 | 0.999907 | intergenic_region | EPDR1:STARD3NL |
| rs112699728;rs61007149 | 7:38113064_T_TTTC | 7 | 38113064 | TTTC | T | 0.712635 | 0.0696399 | 0.014972 | 3.90E-06 | 0.999801 | intergenic_region | EPDR1:STARD3NL |
| rs10716170;rs869042266 | 11:86740417_GA_G | 11 | 86740417 | G | GA | 0.368899 | 0.0649351 | 0.0140282 | 3.90E-06 | 0.995496 | downstream_gene_variant | RP11-736K20.1 |
| rs2001411 | 11:86771583_A_G | 11 | 86771583 | G | A | 0.59979 | -0.0633892 | 0.0137168 | 3.90E-06 | 0.999899 | intron_variant | TMEM135 |
| rs2512358 | 11:86775294_C_T | 11 | 86775294 | T | C | 0.59939 | -0.0635451 | 0.0137377 | 3.90E-06 | 0.995551 | intron_variant | TMEM135 |
| rs11226865 | 11:97382880_G_A | 11 | 97382880 | A | G | 0.160321 | 0.0848984 | 0.0183429 | 3.90E-06 | 1 | intergenic_region | RP11-882G5.1:RNA5SP347 |
| rs12029258 | 1:22686493_C_A | 1 | 22686493 | A | C | 0.398072 | -0.0644644 | 0.0137439 | 4.00E-06 | 0.998711 | downstream_gene_variant | RP11-415K20.2 |
| rs16826981 | 1:22691397_C_T | 1 | 22691397 | T | C | 0.397818 | -0.0645113 | 0.0137585 | 4.00E-06 | 0.997904 | non_coding_transcript_exon_variant | RP11-415K20.2 |
| rs936709393 | 1:22701274_T_TTTTA | 1 | 22701274 | TTTTA | T | 0.40017 | -0.0642847 | 0.0137613 | 4.00E-06 | 0.996471 | intergenic_region | RP11-415K20.2:ZBTB40 |
| | 6:28680686_ATAT_A | 6 | 28680686 | A | ATAT | 0.0430704 | 0.21815 | 0.0469514 | 4.00E-06 | 0.503039 | intergenic_region | C6orf38:RPSAP2 |
| rs1618737 | 7:38101469_C_A | 7 | 38101469 | A | C | 0.712305 | 0.0695676 | 0.014971 | 4.00E-06 | 0.999623 | intergenic_region | EPDR1:STARD3NL |
| rs1721382 | 7:38104983_C_A | 7 | 38104983 | A | C | 0.712288 | 0.0696318 | 0.0149702 | 4.00E-06 | 0.999747 | intergenic_region | EPDR1:STARD3NL |
| rs1717721 | 7:38111340_C_T | 7 | 38111340 | T | C | 0.712767 | 0.0695862 | 0.0149725 | 4.00E-06 | 0.999894 | intergenic_region | EPDR1:STARD3NL |
| rs1403987 | 7:38111941_A_G | 7 | 38111941 | G | A | 0.71277 | 0.069546 | 0.0149724 | 4.00E-06 | 0.999899 | intergenic_region | EPDR1:STARD3NL |
| rs12546164 | 8:127903238_C_T | 8 | 127903238 | T | C | 0.185506 | -0.0814048 | 0.0174798 | 4.00E-06 | 0.987966 | intragenic_variant | PCAT1 |
| rs4514445 | 11:86739604_A_G | 11 | 86739604 | G | A | 0.375436 | 0.0641517 | 0.0139366 | 4.00E-06 | 0.998496 | downstream_gene_variant | RP11-736K20.1 |
| rs2077594 | 11:86772166_G_A | 11 | 86772166 | A | G | 0.599789 | -0.0633864 | 0.0137167 | 4.00E-06 | 0.999918 | intron_variant | TMEM135 |
| rs2512395 | 11:86786129_C_G | 11 | 86786129 | G | C | 0.598923 | -0.0633514 | 0.0137207 | 4.00E-06 | 0.998401 | intron_variant | TMEM135 |
| rs11533276 | 11:86817219_C_A | 11 | 86817219 | A | C | 0.156995 | 0.0864082 | 0.0186254 | 4.00E-06 | 0.998702 | intron_variant | TMEM135 |
| rs111538746 | 15:93330538_C_A | 15 | 93330538 | A | C | 0.0795315 | -0.116392 | 0.0254149 | 4.00E-06 | 0.987414 | intergenic_region | CTD-2313J17.5:CTD-2313J17.4 |
| rs55936687 | 1:22687882_G_T | 1 | 22687882 | T | G | 0.397936 | -0.0644556 | 0.0137463 | 4.10E-06 | 0.998689 | downstream_gene_variant | RP11-415K20.2 |
| rs1524064 | 7:38141380_T_A | 7 | 38141380 | A | T | 0.712733 | 0.0694743 | 0.0149756 | 4.10E-06 | 0.999909 | intergenic_region | EPDR1:STARD3NL |
| rs11459506;rs527285245;rs961110254 | 11:86771689_CT_CTT | 11 | 86771689 | CTT | CT | 0.58851 | -0.0638617 | 0.0138577 | 4.10E-06 | 0.970507 | intron_variant | TMEM135 |
| rs11234952 | 11:86785038_C_T | 11 | 86785038 | T | C | 0.156323 | 0.0866257 | 0.0186945 | 4.10E-06 | 0.998493 | intron_variant | TMEM135 |
| rs72065188;rs72456605 | 11:86785471_CTG_C | 11 | 86785471 | C | CTG | 0.156348 | 0.0866631 | 0.0186935 | 4.10E-06 | 0.998351 | intron_variant | TMEM135 |
| rs1531147 | 11:86785473_G_A | 11 | 86785473 | A | G | 0.599291 | -0.063301 | 0.0137154 | 4.10E-06 | 0.999247 | intron_variant | TMEM135 |
| rs552253 | 11:86903085_G_A | 11 | 86903085 | A | G | 0.538234 | -0.0647358 | 0.0140942 | 4.10E-06 | 0.924685 | intron_variant | TMEM135 |
| rs12029320 | 1:22686692_C_T | 1 | 22686692 | T | C | 0.397585 | -0.0644416 | 0.0137556 | 4.20E-06 | 0.997652 | downstream_gene_variant | RP11-415K20.2 |
| rs55739621 | 1:22688018_A_G | 1 | 22688018 | G | A | 0.412139 | -0.0637162 | 0.0136697 | 4.20E-06 | 0.998583 | downstream_gene_variant | RP11-415K20.2 |
| rs12047557 | 1:22699450_G_GTGGAA | 1 | 22699450 | GTGGAA | G | 0.404606 | -0.0643426 | 0.0137846 | 4.20E-06 | 0.988077 | intergenic_region | RP11-415K20.2:ZBTB40 |
| rs200896539 | 2:176941730_G_GA | 2 | 176941730 | GA | G | 0.278679 | 0.0694998 | 0.0150613 | 4.20E-06 | 0.997628 | downstream_gene_variant | EVX2 |
| rs1357651 | 7:38097862_T_G | 7 | 38097862 | G | T | 0.712336 | 0.0694603 | 0.0149724 | 4.20E-06 | 0.99939 | intergenic_region | EPDR1:STARD3NL |
| rs2462184 | 7:38115495_G_C | 7 | 38115495 | C | G | 0.712662 | 0.0694406 | 0.0149713 | 4.20E-06 | 0.999918 | intergenic_region | EPDR1:STARD3NL |
| rs3956749 | 7:38142832_G_A | 7 | 38142832 | A | G | 0.712739 | 0.0694469 | 0.0149758 | 4.20E-06 | 0.999928 | intergenic_region | EPDR1:STARD3NL |
| rs11234950 | 11:86784785_C_T | 11 | 86784785 | T | C | 0.156266 | 0.0865429 | 0.0186999 | 4.20E-06 | 0.998284 | intron_variant | TMEM135 |
| rs746598843 | 2:176941733_C_CGGTACT | 2 | 176941733 | CGGTACT | C | 0.278642 | 0.0694474 | 0.0150637 | 4.30E-06 | 0.997451 | downstream_gene_variant | EVX2 |
| rs2285504 | 7:92987321_G_A | 7 | 92987321 | A | G | 0.195866 | -0.0792745 | 0.0172693 | 4.30E-06 | 0.987136 | intron_variant | VPS50 |
| rs200533649 | 11:42682543_A_AT | 11 | 42682543 | AT | A | 0.138969 | -0.0902181 | 0.0195532 | 4.30E-06 | 0.997799 | intergenic_region | CTD-2572N17.1:CTD-2537L20.1 |
| rs3045939;rs72042001;rs773722597;rs869041270 | 11:86779850_CAAA_CAA | 11 | 86779850 | CAA | CAAA | 0.180026 | 0.0856531 | 0.0185116 | 4.30E-06 | 0.905278 | intron_variant | TMEM135 |
| rs4944666 | 11:86790944_C_T | 11 | 86790944 | T | C | 0.717879 | -0.0709733 | 0.0153566 | 4.30E-06 | 0.950652 | intron_variant | TMEM135 |
| rs1717726 | 7:38104907_T_A | 7 | 38104907 | A | T | 0.712157 | 0.0693182 | 0.0149734 | 4.40E-06 | 0.999272 | intergenic_region | EPDR1:STARD3NL |
| rs10280939 | 7:38120073_G_A | 7 | 38120073 | A | G | 0.712667 | 0.0692932 | 0.0149708 | 4.40E-06 | 0.999945 | intergenic_region | EPDR1:STARD3NL |
| rs7793606 | 7:38132369_C_T | 7 | 38132369 | T | C | 0.712652 | 0.0692379 | 0.0149712 | 4.40E-06 | 0.999898 | intergenic_region | EPDR1:STARD3NL |
| rs6963134 | 7:38144898_G_A | 7 | 38144898 | G | A | 0.712803 | 0.0693174 | 0.0149792 | 4.40E-06 | 1 | intergenic_region | EPDR1:STARD3NL |
| rs397800921;rs5793236 | 11:86824781_C_CTTTG | 11 | 86824781 | CTTTG | C | 0.724689 | -0.0697372 | 0.0151231 | 4.40E-06 | 0.994882 | intron_variant | TMEM135 |
| rs2512336 | 11:86825512_G_A | 11 | 86825512 | A | G | 0.72563 | -0.0697907 | 0.0151296 | 4.40E-06 | 0.99624 | intron_variant | TMEM135 |
| rs182398571 | 3:156306198_C_G | 3 | 156306198 | G | C | 0.0240319 | -0.208172 | 0.0451111 | 4.50E-06 | 0.950646 | intergenic_region | SSR3:RP11-791I24.4 |
| rs1524067 | 7:38122969_C_A | 7 | 38122969 | A | C | 0.71267 | 0.0691995 | 0.0149705 | 4.50E-06 | 0.999969 | intergenic_region | EPDR1:STARD3NL |
| rs1464712 | 7:38125584_A_G | 7 | 38125584 | G | A | 0.712671 | 0.0691682 | 0.0149705 | 4.50E-06 | 0.999969 | intergenic_region | EPDR1:STARD3NL |
| rs1524065 | 7:38126589_C_A | 7 | 38126589 | A | C | 0.712672 | 0.0691652 | 0.0149706 | 4.50E-06 | 0.999963 | intergenic_region | EPDR1:STARD3NL |
| rs6959212 | 7:38128326_T_C | 7 | 38128326 | C | T | 0.712668 | 0.0691824 | 0.0149708 | 4.50E-06 | 0.999943 | intergenic_region | EPDR1:STARD3NL |
| rs11234940 | 11:86773929_G_A | 11 | 86773929 | A | G | 0.155364 | 0.08657 | 0.0187383 | 4.50E-06 | 0.99955 | intron_variant | TMEM135 |
| rs10898590 | 11:86775026_G_A | 11 | 86775026 | A | G | 0.155365 | 0.0865603 | 0.0187382 | 4.50E-06 | 0.999562 | intron_variant | TMEM135 |
| rs12360980 | 11:86775998_T_C | 11 | 86775998 | C | T | 0.155365 | 0.0865569 | 0.0187381 | 4.50E-06 | 0.999564 | intron_variant | TMEM135 |
| rs11234906 | 11:86723138_T_A | 11 | 86723138 | A | T | 0.149124 | 0.0887704 | 0.0192085 | 4.60E-06 | 0.983559 | intergenic_region | RP11-736K20.6:RP11-736K20.1 |
| rs11374133;rs373022667;rs66545611 | 11:86899746_C_CA | 11 | 86899746 | CA | C | 0.310038 | 0.0673083 | 0.0147926 | 4.60E-06 | 0.97425 | intron_variant | TMEM135 |
| rs2512394 | 11:86786626_T_C | 11 | 86786626 | C | T | 0.725011 | -0.069395 | 0.0150952 | 4.70E-06 | 0.998914 | intron_variant | TMEM135 |
| rs2512393 | 11:86786787_T_C | 11 | 86786787 | C | T | 0.725001 | -0.069376 | 0.0150952 | 4.70E-06 | 0.998909 | intron_variant | TMEM135 |
| rs11374133;rs373022667;rs66545611 | 11:86899746_C_CAA | 11 | 86899746 | CAA | C | 0.686026 | -0.0671744 | 0.0147846 | 4.70E-06 | 0.967642 | intron_variant | TMEM135 |
| rs182075513 | 3:156663378_A_C | 3 | 156663378 | C | A | 0.0242052 | -0.203263 | 0.0445184 | 4.80E-06 | 0.977674 | intron_variant | LEKR1 |
| rs2124581 | 11:86788064_A_G | 11 | 86788064 | G | A | 0.724875 | -0.06932 | 0.0150947 | 4.80E-06 | 0.998826 | intron_variant | TMEM135 |
| rs2130153 | 11:86824568_A_T | 11 | 86824568 | T | A | 0.725156 | -0.0694279 | 0.0151195 | 4.80E-06 | 0.996159 | intron_variant | TMEM135 |
| rs2445556 | 11:86825360_C_T | 11 | 86825360 | T | C | 0.725346 | -0.0694584 | 0.0151198 | 4.80E-06 | 0.996702 | intron_variant | TMEM135 |
| rs847201 | 2:176954111_A_G | 2 | 176954111 | G | A | 0.457661 | 0.0623116 | 0.0135234 | 4.90E-06 | 0.998565 | upstream_gene_variant | HOXD13 |
| rs5793233;rs111987507;rs543112646 | 11:86782716_AT_ATT | 11 | 86782716 | ATT | AT | 0.155104 | 0.0863451 | 0.0187809 | 4.90E-06 | 0.998583 | intron_variant | TMEM135 |
| rs2445579 | 11:86787597_T_A | 11 | 86787597 | A | T | 0.724955 | -0.069273 | 0.0150943 | 4.90E-06 | 0.998988 | intron_variant | TMEM135 |
| rs2512345 | 11:86811858_T_C | 11 | 86811858 | C | T | 0.724379 | -0.0692029 | 0.0150858 | 4.90E-06 | 0.999218 | upstream_gene_variant | RP11-648O15.1 |
| rs2124578 | 11:86823377_C_T | 11 | 86823377 | T | C | 0.724873 | -0.0693018 | 0.0151082 | 4.90E-06 | 0.996882 | intron_variant | TMEM135 |
| rs35243492;rs397709897 | 11:86823435_G_GT | 11 | 86823435 | GT | G | 0.724885 | -0.0693081 | 0.015109 | 4.90E-06 | 0.996807 | intron_variant | TMEM135 |
| rs2513229 | 11:86823562_G_T | 11 | 86823562 | T | G | 0.724917 | -0.0693265 | 0.0151107 | 4.90E-06 | 0.996648 | intron_variant | TMEM135 |
| rs2512333 | 11:86823808_T_A | 11 | 86823808 | A | T | 0.724974 | -0.0693635 | 0.0151138 | 4.90E-06 | 0.996406 | intron_variant | TMEM135 |
| rs2512334 | 11:86824297_G_A | 11 | 86824297 | A | G | 0.725094 | -0.0694124 | 0.0151181 | 4.90E-06 | 0.99616 | intron_variant | TMEM135 |
| rs1320596 | 1:22689804_G_A | 1 | 22689804 | A | G | 0.397505 | -0.0638693 | 0.0137466 | 5.00E-06 | 0.998806 | downstream_gene_variant | RP11-415K20.2 |
| rs4944661 | 11:86738825_T_C | 11 | 86738825 | C | T | 0.376166 | 0.0633881 | 0.0139247 | 5.00E-06 | 0.999315 | downstream_gene_variant | RP11-736K20.1 |
| rs2445577 | 11:86788565_G_A | 11 | 86788565 | A | G | 0.724927 | -0.0692095 | 0.0150933 | 5.00E-06 | 0.999131 | intron_variant | TMEM135 |
| rs2445576 | 11:86788952_G_A | 11 | 86788952 | A | G | 0.724923 | -0.0691965 | 0.0150931 | 5.00E-06 | 0.999154 | intron_variant | TMEM135 |
| rs11234961 | 11:86804081_C_G | 11 | 86804081 | G | C | 0.159847 | 0.0849653 | 0.0184968 | 5.00E-06 | 0.997107 | downstream_gene_variant | RP11-648O15.1 |
| rs6592334 | 11:86805789_C_A | 11 | 86805789 | A | C | 0.159846 | 0.0849661 | 0.0184968 | 5.00E-06 | 0.997108 | intron_variant | TMEM135 |
| rs11234962 | 11:86806583_T_C | 11 | 86806583 | C | T | 0.159825 | 0.0849587 | 0.018496 | 5.00E-06 | 0.99721 | intron_variant | TMEM135 |
| rs11234964 | 11:86807959_A_G | 11 | 86807959 | G | A | 0.159889 | 0.0849397 | 0.0184961 | 5.00E-06 | 0.996825 | upstream_gene_variant | RP11-648O15.1 |
| rs2445554 | 11:86822774_A_G | 11 | 86822774 | G | A | 0.724756 | -0.0692191 | 0.0150972 | 5.00E-06 | 0.998082 | intron_variant | TMEM135 |
| rs2445555 | 11:86823051_A_G | 11 | 86823051 | G | A | 0.724796 | -0.069247 | 0.0151027 | 5.00E-06 | 0.997402 | intron_variant | TMEM135 |
| rs142526455 | 13:94941406_C_T | 13 | 94941406 | T | C | 0.0210031 | -0.222759 | 0.0488433 | 5.00E-06 | 0.93721 | intron_variant | GPC6 |
| rs1717720 | 7:38112615_A_G | 7 | 38112615 | G | A | 0.712319 | 0.0688432 | 0.0149742 | 5.10E-06 | 0.999396 | intergenic_region | EPDR1:STARD3NL |
| rs1717719 | 7:38112622_G_A | 7 | 38112622 | A | G | 0.712319 | 0.0688432 | 0.0149742 | 5.10E-06 | 0.999396 | intergenic_region | EPDR1:STARD3NL |
| rs989689 | 11:86821886_A_G | 11 | 86821886 | G | A | 0.724585 | -0.0690435 | 0.0150807 | 5.10E-06 | 0.999935 | intron_variant | TMEM135 |
| rs2168467 | 11:86822290_G_A | 11 | 86822290 | A | G | 0.72464 | -0.0691053 | 0.0150863 | 5.10E-06 | 0.999293 | intron_variant | TMEM135 |
| rs200088578;rs759589811 | 13:59266344_A_AAAAT | 13 | 59266344 | AAAAT | A | 0.101431 | -0.105254 | 0.0232723 | 5.10E-06 | 0.935153 | intergenic_region | RP11-339I24.1:RP11-522F22.1 |
| rs6758117 | 2:176947237_C_T | 2 | 176947237 | T | C | 0.458021 | 0.061983 | 0.0135347 | 5.20E-06 | 0.996811 | intron_variant | EVX2 |
| | 8:41331299_AAAAAAATATATATATATATATATATAT_AATATATAT | 8 | 41331299 | AATATATAT | AAAAAAATATATATATATATATATATAT | 0.133035 | 0.0950082 | 0.0210639 | 5.20E-06 | 0.887879 | intergenic_region | RP11-15G16.1:GOLGA7 |
| rs11234930 | 11:86764925_T_G | 11 | 86764925 | G | T | 0.154776 | 0.0861609 | 0.0187908 | 5.20E-06 | 0.997937 | intron_variant | TMEM135 |
| rs2445542 | 11:86811014_A_C | 11 | 86811014 | C | A | 0.72452 | -0.0690336 | 0.0150819 | 5.20E-06 | 0.999785 | upstream_gene_variant | RP11-648O15.1 |
| rs2513207 | 11:86811337_T_G | 11 | 86811337 | G | T | 0.724519 | -0.0690327 | 0.0150819 | 5.20E-06 | 0.999789 | upstream_gene_variant | RP11-648O15.1 |
| rs2445544 | 11:86814395_C_A | 11 | 86814395 | A | C | 0.72452 | -0.0690195 | 0.0150814 | 5.20E-06 | 0.999854 | intron_variant | TMEM135 |
| rs2445545 | 11:86814889_G_C | 11 | 86814889 | C | G | 0.72452 | -0.0690161 | 0.0150813 | 5.20E-06 | 0.999865 | intron_variant | TMEM135 |
| rs2445547 | 11:86815037_C_A | 11 | 86815037 | A | C | 0.72452 | -0.0690161 | 0.0150813 | 5.20E-06 | 0.999868 | intron_variant | TMEM135 |
| rs2513230 | 11:86815320_G_A | 11 | 86815320 | A | G | 0.72452 | -0.0690146 | 0.0150812 | 5.20E-06 | 0.999875 | intron_variant | TMEM135 |
| rs2445548 | 11:86815590_G_A | 11 | 86815590 | A | G | 0.724521 | -0.0690107 | 0.0150812 | 5.20E-06 | 0.999881 | intron_variant | TMEM135 |
| rs2512353 | 11:86817495_T_C | 11 | 86817495 | C | T | 0.72452 | -0.0689985 | 0.0150808 | 5.20E-06 | 0.999929 | intron_variant | TMEM135 |
| rs2513231 | 11:86819246_C_T | 11 | 86819246 | T | C | 0.724521 | -0.0689902 | 0.0150804 | 5.20E-06 | 0.999977 | intron_variant | TMEM135 |
| rs2512352 | 11:86819979_T_C | 11 | 86819979 | C | T | 0.72453 | -0.0689993 | 0.0150805 | 5.20E-06 | 0.999958 | intron_variant | TMEM135 |
| rs2512351 | 11:86820971_G_A | 11 | 86820971 | A | G | 0.72453 | -0.0690187 | 0.0150815 | 5.20E-06 | 0.999781 | intron_variant | TMEM135 |
| rs2451067 | 11:86821081_C_T | 11 | 86821081 | T | C | 0.724566 | -0.0690286 | 0.0150807 | 5.20E-06 | 0.999925 | intron_variant | TMEM135 |
| rs2445553 | 11:86821489_C_T | 11 | 86821489 | T | C | 0.724579 | -0.0690379 | 0.0150804 | 5.20E-06 | 0.999954 | intron_variant | TMEM135 |
| rs12223784 | 11:86765802_G_T | 11 | 86765802 | T | G | 0.154785 | 0.0860922 | 0.018789 | 5.30E-06 | 0.998108 | intron_variant | TMEM135 |
| rs2512390 | 11:86790003_A_G | 11 | 86790003 | G | A | 0.724589 | -0.0689687 | 0.0150871 | 5.30E-06 | 0.999188 | intron_variant | TMEM135 |
| rs2512388 | 11:86790855_A_G | 11 | 86790855 | G | A | 0.722829 | -0.0690421 | 0.0151018 | 5.30E-06 | 0.994312 | intron_variant | TMEM135 |
| rs2512363 | 11:86791973_A_C | 11 | 86791973 | C | A | 0.72452 | -0.0689424 | 0.0150881 | 5.30E-06 | 0.998966 | intron_variant | TMEM135 |
| rs4396314 | 11:86822412_T_G | 11 | 86822412 | G | T | 0.724617 | -0.0689796 | 0.0150905 | 5.30E-06 | 0.998791 | intron_variant | TMEM135 |
| rs35666829 | 11:86822418_T_G | 11 | 86822418 | G | T | 0.724622 | -0.068986 | 0.0150908 | 5.30E-06 | 0.998764 | intron_variant | TMEM135 |
| rs79411098 | 15:93326726_C_G | 15 | 93326726 | G | C | 0.0902978 | -0.108727 | 0.0240206 | 5.30E-06 | 0.975243 | upstream_gene_variant | CTD-2313J17.5 |
| rs11234909 | 11:86733470_G_T | 11 | 86733470 | T | G | 0.154109 | 0.0857779 | 0.0187869 | 5.40E-06 | 0.998438 | upstream_gene_variant | RP11-736K20.1 |
| rs11234910 | 11:86733474_A_G | 11 | 86733474 | G | A | 0.154109 | 0.0857779 | 0.0187869 | 5.40E-06 | 0.998438 | upstream_gene_variant | RP11-736K20.1 |
| rs577953435 | 11:86743300_ATTATTTAT_ATTATTTATTTATTTATTTAT | 11 | 86743300 | ATTATTTATTTATTTATTTAT | ATTATTTAT | 0.163314 | 0.0847693 | 0.0186583 | 5.40E-06 | 0.955629 | intergenic_region | RP11-736K20.1:TMEM135 |
| rs10898580 | 11:86745532_T_G | 11 | 86745532 | G | T | 0.154782 | 0.0854888 | 0.018751 | 5.40E-06 | 0.998866 | upstream_gene_variant | TMEM135 |
| rs11234932 | 11:86766666_C_T | 11 | 86766666 | T | C | 0.154799 | 0.0859699 | 0.0187855 | 5.40E-06 | 0.998441 | intron_variant | TMEM135 |
| rs10898588 | 11:86769676_A_G | 11 | 86769676 | G | A | 0.155084 | 0.0859806 | 0.0187658 | 5.40E-06 | 0.998422 | intron_variant | TMEM135 |
| rs2445550 | 11:86816943_T_C | 11 | 86816943 | C | T | 0.724256 | -0.0689425 | 0.0150874 | 5.40E-06 | 0.998701 | intron_variant | TMEM135 |
| rs76511445 | 15:93324443_A_G | 15 | 93324443 | G | A | 0.0902355 | -0.107152 | 0.0237151 | 5.40E-06 | 0.99915 | non_coding_transcript_exon_variant | CTD-2313J17.5 |
| rs549530064;rs140968772 | 15:93324875_CA_C | 15 | 93324875 | C | CA | 0.0896491 | -0.108927 | 0.0241443 | 5.40E-06 | 0.971309 | upstream_gene_variant | CTD-2313J17.5 |
| rs140047050;rs773617508;rs535977836 | 8:143298664_AAAAGAAAGAAAGAAAGAAAGAAAGAAAGAAAGAAAGAAAG_AAAAGAAAGAAAGAAAG | 8 | 143298664 | AAAAGAAAGAAAGAAAG | AAAAGAAAGAAAGAAAGAAAGAAAGAAAGAAAGAAAGAAAG | 0.0480236 | 0.158281 | 0.0345349 | 5.50E-06 | 0.826161 | intron_variant | TSNARE1 |
| rs75067134 | 11:42680945_T_C | 11 | 42680945 | C | T | 0.138241 | -0.089327 | 0.0195762 | 5.50E-06 | 0.999778 | intergenic_region | CTD-2572N17.1:CTD-2537L20.1 |
| rs79938891 | 11:42681531_T_G | 11 | 42681531 | G | T | 0.138243 | -0.0893309 | 0.019576 | 5.50E-06 | 0.999784 | intergenic_region | CTD-2572N17.1:CTD-2537L20.1 |
| rs76073944 | 11:42682531_T_C | 11 | 42682531 | C | T | 0.138249 | -0.0893316 | 0.0195765 | 5.50E-06 | 0.99973 | intergenic_region | CTD-2572N17.1:CTD-2537L20.1 |
| rs11234911 | 11:86733797_T_C | 11 | 86733797 | C | T | 0.15411 | 0.0857724 | 0.0187867 | 5.50E-06 | 0.998465 | upstream_gene_variant | RP11-736K20.1 |
| rs11234933 | 11:86766753_C_T | 11 | 86766753 | T | C | 0.1548 | 0.085959 | 0.0187852 | 5.50E-06 | 0.998477 | intron_variant | TMEM135 |
| rs2445582 | 11:86795619_A_G | 11 | 86795619 | G | A | 0.724546 | -0.0688609 | 0.0150846 | 5.50E-06 | 0.999572 | intron_variant | TMEM135 |
| rs2513204 | 11:86796104_T_A | 11 | 86796104 | A | T | 0.724545 | -0.0688527 | 0.0150845 | 5.50E-06 | 0.999588 | intron_variant | TMEM135 |
| rs2512381 | 11:86797999_C_T | 11 | 86797999 | T | C | 0.724658 | -0.0689072 | 0.0150953 | 5.50E-06 | 0.998372 | intron_variant | TMEM135 |
| rs2445569 | 11:86798422_G_A | 11 | 86798422 | A | G | 0.724566 | -0.0688594 | 0.015086 | 5.50E-06 | 0.999479 | intron_variant | TMEM135 |
| rs2513221 | 11:86798940_A_G | 11 | 86798940 | G | A | 0.724566 | -0.0688595 | 0.015086 | 5.50E-06 | 0.999479 | intron_variant | TMEM135 |
| rs2445568 | 11:86799769_C_T | 11 | 86799769 | T | C | 0.724566 | -0.0688595 | 0.015086 | 5.50E-06 | 0.999479 | intron_variant | TMEM135 |
| rs2445567 | 11:86799830_T_C | 11 | 86799830 | C | T | 0.724566 | -0.0688595 | 0.015086 | 5.50E-06 | 0.99948 | intron_variant | TMEM135 |
| rs2512382 | 11:86800848_G_A | 11 | 86800848 | A | G | 0.724566 | -0.0688621 | 0.015086 | 5.50E-06 | 0.999479 | downstream_gene_variant | RP11-648O15.1 |
| rs2445566 | 11:86802000_A_C | 11 | 86802000 | C | A | 0.724547 | -0.0688214 | 0.0150845 | 5.50E-06 | 0.999597 | downstream_gene_variant | RP11-648O15.1 |
| rs10898595 | 11:86802260_G_A | 11 | 86802260 | A | G | 0.159815 | 0.0845623 | 0.0184981 | 5.50E-06 | 0.997054 | downstream_gene_variant | RP11-648O15.1 |
| rs199728646 | 10:69610832_A_C | 10 | 69610832 | C | A | 0.279305 | 0.069117 | 0.0151563 | 5.60E-06 | 0.990293 | downstream_gene_variant | RP11-57G10.8 |
| rs2513218 | 11:86754210_T_C | 11 | 86754210 | C | T | 0.594231 | -0.0625315 | 0.0137556 | 5.60E-06 | 0.990255 | intron_variant | TMEM135 |
| rs4451755 | 11:86822471_C_T | 11 | 86822471 | T | C | 0.72277 | -0.0689739 | 0.015111 | 5.60E-06 | 0.991859 | intron_variant | TMEM135 |
| rs35171130;rs372707663 | 9:14777786_AC_A | 9 | 14777786 | A | AC | 0.0132892 | -0.267015 | 0.0596047 | 5.70E-06 | 0.978901 | intron_variant | FREM1 |
| rs1243846892 | 10:69610831_C_CT | 10 | 69610831 | CT | C | 0.279086 | 0.0691534 | 0.0151646 | 5.70E-06 | 0.990181 | downstream_gene_variant | RP11-57G10.8 |

| | | | | | | | | | | | | |
|---|---|---|---|---|---|---|---|---|---|---|---|---|
| rs2512380 | 11:86797536_A_G | 11 | 86797536 | G | A | 0.724526 | -0.0687534 | 0.0150856 | 5.70E-06 | 0.999331 | intron_variant | TMEM135 |
| rs771965258 | 19:14729658_CAAAAAAA_CAAAAAAAAAAA | 19 | 14729658 | CAAAAAAAAAAA | CAAAAAAA | 0.0284236 | 0.248664 | 0.0551129 | 5.70E-06 | 0.528367 | downstream_gene_variant | ITGB1P1 |
| rs79082223 | 8:4835778_G_C | 8 | 4835778 | C | G | 0.196044 | -0.0790018 | 0.0175051 | 5.80E-06 | 0.949725 | intron_variant | CSMD1 |
| rs761522335;rs71844975;rs766162061;rs71470030 | 11:6825930_TACACACACACAC_TACACAC | 11 | 6825930 | TACACAC | TACACACACACAC | 0.157333 | -0.0909075 | 0.0202834 | 5.80E-06 | 0.848613 | intergenic_region | OR6A2:RP11-413N10.3 |
| rs78608900 | 11:42683589_C_T | 11 | 42683589 | T | C | 0.138675 | -0.0889279 | 0.0195506 | 6.00E-06 | 0.997854 | intergenic_region | CTD-2572N17.1:CTD-2537L20.1 |
| rs10898589 | 11:86771207_G_A | 11 | 86771207 | A | G | 0.15512 | 0.0855818 | 0.0187594 | 6.00E-06 | 0.998861 | intron_variant | TMEM135 |
| rs11234937 | 11:86771317_T_A | 11 | 86771317 | A | T | 0.15512 | 0.0855702 | 0.0187592 | 6.00E-06 | 0.998879 | intron_variant | TMEM135 |
| rs2445584 | 11:86791877_T_C | 11 | 86791877 | C | T | 0.724477 | -0.068569 | 0.0150914 | 6.10E-06 | 0.99814 | intron_variant | TMEM135 |
| rs2130154 | 11:86821534_G_A | 11 | 86821534 | G | A | 0.724569 | -0.0685259 | 0.0150835 | 6.10E-06 | 1 | intron_variant | TMEM135 |
| rs140942370 | 11:86742354_CT_C | 11 | 86742354 | C | CT | 0.154143 | 0.0852536 | 0.0187803 | 6.20E-06 | 0.999115 | intergenic_region | RP11-736K20.1:TMEM135 |
| rs11234923 | 11:86744756_G_A | 11 | 86744756 | A | G | 0.154029 | 0.085299 | 0.0187889 | 6.20E-06 | 0.998584 | upstream_gene_variant | TMEM135 |
| rs10792897 | 11:86722402_C_G | 11 | 86722402 | G | C | 0.14445 | 0.0887207 | 0.0194562 | 6.30E-06 | 0.985283 | intergenic_region | RP11-736K20.6:RP11-736K20.1 |
| rs397821680;rs57086268;rs376153742;rs796287066 | 11:86775304_CTT_CTTT | 11 | 86775304 | CTTT | CTT | 0.580265 | -0.0636674 | 0.0140302 | 6.30E-06 | 0.942499 | intron_variant | TMEM135 |
| rs34801765;rs533348054;rs148372387 | 11:86841158_ATT_AT | 11 | 86841158 | AT | ATT | 0.540404 | -0.0606106 | 0.0135794 | 6.30E-06 | 0.984977 | intron_variant | TMEM135 |
| rs200810002;rs34161815;rs775373176 | 11:86769368_T_TTTA | 11 | 86769368 | TTTA | T | 0.572518 | -0.062662 | 0.0137945 | 6.40E-06 | 0.973737 | intron_variant | TMEM135 |
| rs111920036 | 6:70277040_CAT_CATATATATATCATATATATGATATATATATGATATTATATATGATATGAT | 6 | 70277040 | CATATATATATCATATATATGATATATATATGATATTATATATGATATGAT | CAT | 0.064324 | -0.16548 | 0.0356372 | 6.50E-06 | 0.602996 | intergenic_region | ADGRB3:AL358133.1 |
| rs11234925 | 11:86748051_G_T | 11 | 86748051 | T | G | 0.154205 | 0.0850496 | 0.018779 | 6.50E-06 | 0.998905 | upstream_gene_variant | TMEM135 |
| rs2512385 | 11:86802688_G_A | 11 | 86802688 | A | G | 0.722125 | -0.0681946 | 0.0150602 | 6.60E-06 | 0.99667 | downstream_gene_variant | RP11-648O15.1 |
| rs4351839 | 11:86822514_C_G | 11 | 86822514 | G | C | 0.721956 | -0.0684909 | 0.01513 | 6.60E-06 | 0.989633 | intron_variant | TMEM135 |
| rs7694049 | 4:2242921_A_C | 4 | 2242921 | C | A | 0.35913 | 0.0635241 | 0.0140406 | 6.70E-06 | 0.99785 | intron_variant | HAUS3 |
| rs2512501 | 11:86802707_T_A | 11 | 86802707 | A | T | 0.721625 | -0.0680533 | 0.0150438 | 6.70E-06 | 0.998048 | downstream_gene_variant | RP11-648O15.1 |
| rs2512386 | 11:86803834_G_A | 11 | 86803834 | A | G | 0.721854 | -0.0680541 | 0.0150448 | 6.70E-06 | 0.998262 | downstream_gene_variant | RP11-648O15.1 |
| rs2445563 | 11:86804794_G_T | 11 | 86804794 | T | G | 0.721854 | -0.0680541 | 0.0150448 | 6.70E-06 | 0.998263 | downstream_gene_variant | RP11-648O15.1 |
| rs2513226 | 11:86805562_G_A | 11 | 86805562 | A | G | 0.721854 | -0.0680545 | 0.0150448 | 6.70E-06 | 0.998263 | downstream_gene_variant | RP11-648O15.1 |
| rs2513227 | 11:86805605_A_G | 11 | 86805605 | G | A | 0.721854 | -0.0680545 | 0.0150448 | 6.70E-06 | 0.998263 | downstream_gene_variant | RP11-648O15.1 |
| rs2512488 | 11:86805885_A_C | 11 | 86805885 | C | A | 0.721854 | -0.0680541 | 0.0150448 | 6.70E-06 | 0.998264 | intron_variant | TMEM135 |
| rs2445541 | 11:86806071_CA_GA | 11 | 86806071 | GA | CA | 0.721854 | -0.0680542 | 0.0150448 | 6.70E-06 | 0.998265 | intron_variant | TMEM135 |
| rs11325734;rs796101885 | 11:86806279_CA_C | 11 | 86806279 | C | CA | 0.721855 | -0.068037 | 0.0150445 | 6.70E-06 | 0.998298 | intron_variant | TMEM135 |
| rs2445559 | 11:86807676_A_G | 11 | 86807676 | G | A | 0.721857 | -0.0680147 | 0.0150439 | 6.70E-06 | 0.998377 | upstream_gene_variant | RP11-648O15.1 |
| rs2512327 | 11:86808484_G_A | 11 | 86808484 | A | G | 0.721857 | -0.0680147 | 0.0150439 | 6.70E-06 | 0.998379 | upstream_gene_variant | RP11-648O15.1 |
| rs539689774 | 15:93322621_T_A | 15 | 93322621 | A | T | 0.0898784 | -0.106423 | 0.02383 | 6.70E-06 | 0.990238 | downstream_gene_variant | CTD-2313J17.5 |
| rs78071142 | 15:93326769_T_C | 15 | 93326769 | C | T | 0.0907538 | -0.107476 | 0.0239885 | 6.70E-06 | 0.974458 | upstream_gene_variant | CTD-2313J17.5 |
| rs35627094 | 11:86807217_C_CA | 11 | 86807217 | CA | C | 0.721857 | -0.0680118 | 0.0150439 | 6.80E-06 | 0.998375 | intron_variant | TMEM135 |
| rs2512325 | 11:86807228_T_A | 11 | 86807228 | A | T | 0.721857 | -0.0680118 | 0.0150439 | 6.80E-06 | 0.998375 | intron_variant | TMEM135 |
| rs34493003;rs758470014 | 11:86809445_TA_T | 11 | 86809445 | T | TA | 0.721857 | -0.0680061 | 0.0150436 | 6.80E-06 | 0.998406 | upstream_gene_variant | RP11-648O15.1 |
| rs2513209 | 11:86809600_A_G | 11 | 86809600 | G | A | 0.721857 | -0.0680061 | 0.0150436 | 6.80E-06 | 0.998406 | upstream_gene_variant | RP11-648O15.1 |
| rs2512326 | 11:86808094_G_A | 11 | 86808094 | A | G | 0.721622 | -0.0679482 | 0.0150443 | 6.90E-06 | 0.997588 | upstream_gene_variant | RP11-648O15.1 |
| rs117786845 | 1:119300903_G_A | 1 | 119300903 | A | G | 0.0216031 | 0.214375 | 0.0479912 | 7.00E-06 | 0.941991 | intergenic_region | RP4-630J13.1:TBX15 |
| rs2168469 | 11:86793864_A_C | 11 | 86793864 | C | A | 0.721909 | -0.0678892 | 0.0150477 | 7.00E-06 | 0.997976 | intron_variant | TMEM135 |
| rs2513236 | 11:86787372_G_A | 11 | 86787372 | A | G | 0.50042 | -0.0614673 | 0.0136832 | 7.10E-06 | 0.973642 | intron_variant | TMEM135 |
| rs142603759 | 11:86807151_C_CATATATATAAAGAAAT | 11 | 86807151 | CATATATATAAAGAAAT | C | 0.721774 | -0.0677934 | 0.0150419 | 7.20E-06 | 0.998231 | intron_variant | TMEM135 |
| rs12045139 | 1:22681615_G_A | 1 | 22681615 | A | G | 0.407265 | -0.0628838 | 0.0137789 | 7.30E-06 | 0.992368 | intergenic_region | RP11-415K20.1:RP11-415K20.2 |
| rs7124448 | 11:86908718_G_A | 11 | 86908718 | A | G | 0.159747 | 0.0836803 | 0.0185174 | 7.30E-06 | 0.995301 | intron_variant | TMEM135 |
| rs72760032 | 15:93325082_A_T | 15 | 93325082 | T | A | 0.110845 | -0.097303 | 0.0217138 | 7.30E-06 | 0.989744 | upstream_gene_variant | CTD-2313J17.5 |
| rs11736085 | 4:2257376_C_T | 4 | 2257376 | T | C | 0.319881 | 0.0656972 | 0.0145327 | 7.40E-06 | 0.989816 | intron_variant | MXD4 |
| rs11368149;rs796753588 | 11:86809184_AT_A | 11 | 86809184 | A | AT | 0.720543 | -0.0677515 | 0.0150483 | 7.40E-06 | 0.996046 | upstream_gene_variant | RP11-648O15.1 |
| rs116940072 | 16:10873026_T_C | 16 | 10873026 | C | T | 0.0311339 | -0.184674 | 0.0417717 | 7.40E-06 | 0.889368 | intron_variant | TVP23A |
| rs111532899 | 15:93324748_C_T | 15 | 93324748 | T | C | 0.0902017 | -0.105723 | 0.0237623 | 7.50E-06 | 0.995006 | upstream_gene_variant | CTD-2313J17.5 |
| rs2044423 | 2:137423197_C_A | 2 | 137423197 | A | C | 0.246221 | 0.0691097 | 0.0156838 | 7.80E-06 | 0.994819 | intergenic_region | AC092786.1:THSD7B |
| rs274656 | 5:6703039_C_T | 5 | 6703039 | T | C | 0.402447 | -0.0620409 | 0.013899 | 7.80E-06 | 0.984646 | intron_variant | CTD-2044J15.1 |
| rs199977904;rs571202663;rs201227415;rs202111775 | 11:86868116_GTT_GTTT | 11 | 86868116 | GTTT | GTT | 0.211024 | 0.0763863 | 0.0170131 | 7.80E-06 | 0.939427 | intron_variant | TMEM135 |
| rs558128997 | 15:77369541_ATG_A | 15 | 77369541 | A | ATG | 0.249307 | -0.0765676 | 0.017136 | 7.80E-06 | 0.835813 | intragenic_variant | TSPAN3 |
| rs6141 | 3:184090266_C_T | 3 | 184090266 | T | C | 0.458959 | -0.0608721 | 0.0136635 | 7.90E-06 | 1 | 3_prime_UTR_variant | THPO |
| rs371613511 | 7:84286410_CATATATATAT_CAT | 7 | 84286410 | CAT | CATATATATAT | 0.0166514 | -0.25789 | 0.0573845 | 8.00E-06 | 0.860569 | intergenic_region | AC003984.1:AC092022.1 |
| rs74332184 | 11:42681068_G_A | 11 | 42681068 | A | G | 0.0919492 | -0.105637 | 0.0234617 | 8.00E-06 | 1 | intergenic_region | CTD-2572N17.1:CTD-2537L20.1 |
| rs75240041 | 11:42685702_T_G | 11 | 42685702 | G | T | 0.138144 | -0.0878537 | 0.0195893 | 8.00E-06 | 0.998376 | intergenic_region | CTD-2572N17.1:CTD-2537L20.1 |
| rs74417733 | 11:42685765_A_G | 11 | 42685765 | G | A | 0.138147 | -0.0878534 | 0.0195897 | 8.00E-06 | 0.998329 | intergenic_region | CTD-2572N17.1:CTD-2537L20.1 |
| rs76740748 | 11:42677692_C_T | 11 | 42677692 | T | C | 0.0917659 | -0.105696 | 0.0234867 | 8.10E-06 | 0.999174 | intergenic_region | CTD-2572N17.1:CTD-2537L20.1 |
| rs3100847 | 11:86806968_A_G | 11 | 86806968 | G | A | 0.720704 | -0.0675357 | 0.0150551 | 8.10E-06 | 0.993921 | intron_variant | TMEM135 |
| rs116175103 | 15:93325131_A_C | 15 | 93325131 | C | A | 0.090076 | -0.105789 | 0.0238472 | 8.10E-06 | 0.989413 | upstream_gene_variant | CTD-2313J17.5 |
| rs202076092 | 10:31763146_A_G | 10 | 31763146 | G | A | 0.0196727 | -0.220201 | 0.0492763 | 8.20E-06 | 0.961639 | intron_variant | ZEB1 |
| rs567805191 | 6:74509581_AT_ATT | 6 | 74509581 | ATT | AT | 0.0116592 | 0.306272 | 0.0684137 | 8.30E-06 | 0.860732 | intron_variant | CD109 |
| rs71900804;rs762326534;rs869219910;rs71865966;rs3083000;rs755480223;rs770214348 | 6:166069746_TCACACACACACACACACACA_TCACACACACACACACA | 6 | 166069746 | TCACACACACACACACA | TCACACACACACACACACACA | 0.0561888 | 0.1462 | 0.0324498 | 8.30E-06 | 0.823857 | intron_variant | PDE10A |
| rs3063946;rs397837811;rs74776844;rs60279889;rs146096458;rs61583158;rs748754013 | 9:112894312_CGTGTGTGT_CGT | 9 | 112894312 | CGT | CGTGTGTGT | 0.293176 | -0.0668381 | 0.0152027 | 8.40E-06 | 0.9561 | intron_variant | PALM2-AKAP2 |
| rs80055637 | 11:42679176_T_A | 11 | 42679176 | A | T | 0.0917171 | -0.105435 | 0.0234844 | 8.40E-06 | 0.999815 | intergenic_region | CTD-2572N17.1:CTD-2537L20.1 |
| rs78915749 | 11:42679601_C_T | 11 | 42679601 | T | C | 0.0917178 | -0.105457 | 0.0234827 | 8.40E-06 | 0.99997 | intergenic_region | CTD-2572N17.1:CTD-2537L20.1 |
| rs7482565 | 11:42679710_G_A | 11 | 42679710 | A | G | 0.0917176 | -0.10546 | 0.0234825 | 8.40E-06 | 0.999988 | intergenic_region | CTD-2572N17.1:CTD-2537L20.1 |
| rs7479899 | 11:42679785_T_A | 11 | 42679785 | A | T | 0.0917176 | -0.10546 | 0.0234825 | 8.40E-06 | 0.999988 | intergenic_region | CTD-2572N17.1:CTD-2537L20.1 |
| rs75910873 | 11:42680144_A_G | 11 | 42680144 | G | A | 0.091718 | -0.105458 | 0.0234823 | 8.40E-06 | 0.999992 | intergenic_region | CTD-2572N17.1:CTD-2537L20.1 |
| rs77117709 | 11:42680177_C_T | 11 | 42680177 | T | C | 0.091718 | -0.105458 | 0.0234823 | 8.40E-06 | 0.999992 | intergenic_region | CTD-2572N17.1:CTD-2537L20.1 |
| rs11234907 | 11:86728082_G_A | 11 | 86728082 | A | G | 0.152059 | 0.0844851 | 0.0188875 | 8.40E-06 | 0.997281 | intergenic_region | RP11-736K20.6:RP11-736K20.1 |
| rs373486107;rs764335370;rs869082570;rs559750063;rs59117394;rs752823745;rs766840351;rs774627174 | 12:98961841_GCACACACACACACACA_GCACACA | 12 | 98961841 | GCACACA | GCACACACACACACACA | 0.105854 | -0.105964 | 0.0242532 | 8.40E-06 | 0.820548 | intergenic_region | TMPO:RP11-481K9.4 |
| rs75860111 | 11:42678437_A_T | 11 | 42678437 | T | A | 0.0917156 | -0.1054 | 0.0234876 | 8.50E-06 | 0.999547 | intergenic_region | CTD-2572N17.1:CTD-2537L20.1 |
| rs76658142 | 11:42678489_G_A | 11 | 42678489 | A | G | 0.0917169 | -0.105401 | 0.0234871 | 8.50E-06 | 0.999567 | intergenic_region | CTD-2572N17.1:CTD-2537L20.1 |
| rs77550543 | 11:42678795_T_G | 11 | 42678795 | G | T | 0.0917162 | -0.105421 | 0.023486 | 8.50E-06 | 0.999682 | intergenic_region | CTD-2572N17.1:CTD-2537L20.1 |
| rs78767668 | 11:42669895_A_C | 11 | 42669895 | C | A | 0.0917609 | -0.105385 | 0.0234924 | 8.60E-06 | 0.998804 | intergenic_region | CTD-2572N17.1:CTD-2537L20.1 |
| rs76243590 | 11:42676795_G_T | 11 | 42676795 | T | G | 0.0917136 | -0.105347 | 0.0234913 | 8.60E-06 | 0.999228 | intergenic_region | CTD-2572N17.1:CTD-2537L20.1 |
| rs77695437 | 11:42677063_G_A | 11 | 42677063 | A | G | 0.091714 | -0.105351 | 0.0234908 | 8.60E-06 | 0.999269 | intergenic_region | CTD-2572N17.1:CTD-2537L20.1 |
| rs75142590 | 11:42678006_T_A | 11 | 42678006 | A | T | 0.0917153 | -0.10538 | 0.023489 | 8.60E-06 | 0.999427 | intergenic_region | CTD-2572N17.1:CTD-2537L20.1 |
| rs78274397 | 11:42678128_T_G | 11 | 42678128 | G | T | 0.0917153 | -0.105382 | 0.0234887 | 8.60E-06 | 0.999449 | intergenic_region | CTD-2572N17.1:CTD-2537L20.1 |
| rs10898576 | 11:86720187_G_A | 11 | 86720187 | A | G | 0.141218 | 0.0901114 | 0.0200573 | 8.60E-06 | 0.946696 | intergenic_region | RP11-736K20.6:RP11-736K20.1 |
| rs28737222 | 15:93297444_C_T | 15 | 93297444 | T | C | 0.0912878 | -0.106903 | 0.0238879 | 8.60E-06 | 0.976518 | intergenic_region | RP11-386M24.4:CTD-2313J17.3 |
| rs200093157;rs538747110 | 11:42673396_TGATGGATG_TGATGGATGGATG | 11 | 42673396 | TGATGGATGGATG | TGATGGATG | 0.0907766 | -0.1058 | 0.0236115 | 8.70E-06 | 0.997423 | intergenic_region | CTD-2572N17.1:CTD-2537L20.1 |
| rs77894542 | 11:42674930_C_A | 11 | 42674930 | A | C | 0.0917122 | -0.105301 | 0.0234936 | 8.70E-06 | 0.999037 | intergenic_region | CTD-2572N17.1:CTD-2537L20.1 |
| rs77987837 | 11:42675573_G_A | 11 | 42675573 | A | G | 0.091712 | -0.10531 | 0.0234934 | 8.70E-06 | 0.999065 | intergenic_region | CTD-2572N17.1:CTD-2537L20.1 |
| rs117819337 | 11:42676210_A_G | 11 | 42676210 | G | A | 0.0917131 | -0.105324 | 0.0234924 | 8.70E-06 | 0.999137 | intergenic_region | CTD-2572N17.1:CTD-2537L20.1 |
| rs117392879 | 11:42676319_C_T | 11 | 42676319 | T | C | 0.0917133 | -0.105333 | 0.0234922 | 8.70E-06 | 0.999156 | intergenic_region | CTD-2572N17.1:CTD-2537L20.1 |
| rs74604247 | 11:42676417_T_C | 11 | 42676417 | C | T | 0.0917133 | -0.105333 | 0.0234921 | 8.70E-06 | 0.99917 | intergenic_region | CTD-2572N17.1:CTD-2537L20.1 |
| rs79582708 | 11:42676455_T_C | 11 | 42676455 | C | T | 0.0917134 | -0.105332 | 0.0234919 | 8.70E-06 | 0.999178 | intergenic_region | CTD-2572N17.1:CTD-2537L20.1 |
| | 11:86720229_CA_C | 11 | 86720229 | C | CA | 0.141198 | 0.0900206 | 0.0200527 | 8.70E-06 | 0.947201 | intergenic_region | RP11-736K20.6:RP11-736K20.1 |
| rs35962056 | 11:86789834_C_CT | 11 | 86789834 | CT | C | 0.719538 | -0.0671214 | 0.0150724 | 8.70E-06 | 0.991371 | intron_variant | TMEM135 |
| | 17:66918390_G_<DEL> | 17 | 66918390 | <DEL> | G | 0.0104259 | 0.405842 | 0.0915354 | 8.70E-06 | 0.532579 | synonymous_variant | ABCA8 |
| rs77081404 | 11:42663601_G_A | 11 | 42663601 | A | G | 0.0917563 | -0.105298 | 0.0234952 | 8.80E-06 | 0.998641 | intergenic_region | CTD-2572N17.1:CTD-2537L20.1 |
| rs79012583 | 11:42664698_A_G | 11 | 42664698 | G | A | 0.0917579 | -0.105306 | 0.0234946 | 8.80E-06 | 0.998664 | intergenic_region | CTD-2572N17.1:CTD-2537L20.1 |
| rs74364515 | 11:42670791_C_T | 11 | 42670791 | T | C | 0.0917111 | -0.105262 | 0.0234958 | 8.80E-06 | 0.998876 | intergenic_region | CTD-2572N17.1:CTD-2537L20.1 |
| rs80128586 | 11:42671076_C_T | 11 | 42671076 | T | C | 0.0921532 | -0.105292 | 0.0234878 | 8.80E-06 | 0.995866 | intergenic_region | CTD-2572N17.1:CTD-2537L20.1 |
| rs79410009 | 11:42671715_T_A | 11 | 42671715 | A | T | 0.0917113 | -0.105273 | 0.0234954 | 8.80E-06 | 0.998909 | intergenic_region | CTD-2572N17.1:CTD-2537L20.1 |
| rs79498532 | 11:42671731_T_C | 11 | 42671731 | C | T | 0.0917113 | -0.105273 | 0.0234954 | 8.80E-06 | 0.99891 | intergenic_region | CTD-2572N17.1:CTD-2537L20.1 |
| rs77696802 | 11:42671772_T_C | 11 | 42671772 | C | T | 0.0917113 | -0.105273 | 0.0234954 | 8.80E-06 | 0.998911 | intergenic_region | CTD-2572N17.1:CTD-2537L20.1 |
| rs76238931 | 11:42672043_C_T | 11 | 42672043 | T | C | 0.0917111 | -0.105274 | 0.0234953 | 8.80E-06 | 0.99892 | intergenic_region | CTD-2572N17.1:CTD-2537L20.1 |
| rs76114873 | 11:42672153_G_A | 11 | 42672153 | A | G | 0.0917107 | -0.10527 | 0.0234952 | 8.80E-06 | 0.998924 | intergenic_region | CTD-2572N17.1:CTD-2537L20.1 |
| rs59420443 | 11:42672596_T_C | 11 | 42672596 | C | T | 0.0917114 | -0.105274 | 0.023495 | 8.80E-06 | 0.998941 | intergenic_region | CTD-2572N17.1:CTD-2537L20.1 |
| rs75870709 | 11:42673805_C_T | 11 | 42673805 | T | C | 0.0917122 | -0.10529 | 0.0234943 | 8.80E-06 | 0.998988 | intergenic_region | CTD-2572N17.1:CTD-2537L20.1 |
| rs77639471 | 11:42673807_A_G | 11 | 42673807 | G | A | 0.0917122 | -0.10529 | 0.0234943 | 8.80E-06 | 0.998988 | intergenic_region | CTD-2572N17.1:CTD-2537L20.1 |
| rs75079007 | 11:42673820_C_T | 11 | 42673820 | T | C | 0.0917122 | -0.10529 | 0.0234943 | 8.80E-06 | 0.998988 | intergenic_region | CTD-2572N17.1:CTD-2537L20.1 |
| rs78786958 | 11:42673968_G_T | 11 | 42673968 | T | G | 0.0917118 | -0.105288 | 0.0234943 | 8.80E-06 | 0.998995 | intergenic_region | CTD-2572N17.1:CTD-2537L20.1 |
| rs74730169 | 11:42674251_G_A | 11 | 42674251 | A | G | 0.0917118 | -0.105294 | 0.0234941 | 8.80E-06 | 0.999008 | intergenic_region | CTD-2572N17.1:CTD-2537L20.1 |
| rs74407944 | 11:42679342_T_A | 11 | 42679342 | A | T | 0.091502 | -0.105469 | 0.0235399 | 8.80E-06 | 0.997395 | intergenic_region | CTD-2572N17.1:CTD-2537L20.1 |
| rs74688908 | 11:42679344_T_C | 11 | 42679344 | C | T | 0.091502 | -0.105469 | 0.0235399 | 8.80E-06 | 0.997397 | intergenic_region | CTD-2572N17.1:CTD-2537L20.1 |
| rs78396780 | 11:42659934_G_C | 11 | 42659934 | C | G | 0.0917467 | -0.105264 | 0.0235002 | 8.90E-06 | 0.998376 | intergenic_region | CTD-2572N17.1:CTD-2537L20.1 |
| rs74984851 | 11:42661343_C_G | 11 | 42661343 | G | C | 0.0917521 | -0.10524 | 0.0234975 | 8.90E-06 | 0.998518 | intergenic_region | CTD-2572N17.1:CTD-2537L20.1 |
| rs75419756 | 11:42668018_T_A | 11 | 42668018 | A | T | 0.0917105 | -0.10522 | 0.0234971 | 8.90E-06 | 0.998782 | intergenic_region | CTD-2572N17.1:CTD-2537L20.1 |
| rs80030350 | 11:42669755_C_T | 11 | 42669755 | T | C | 0.0917107 | -0.105241 | 0.0234964 | 8.90E-06 | 0.998837 | intergenic_region | CTD-2572N17.1:CTD-2537L20.1 |
| rs74359708 | 11:42670236_G_A | 11 | 42670236 | A | G | 0.091633 | -0.105264 | 0.0235051 | 8.90E-06 | 0.998702 | intergenic_region | CTD-2572N17.1:CTD-2537L20.1 |
| rs77320650 | 11:42670246_T_C | 11 | 42670246 | C | T | 0.091633 | -0.105264 | 0.0235051 | 8.90E-06 | 0.998702 | intergenic_region | CTD-2572N17.1:CTD-2537L20.1 |
| rs78916606 | 11:42670351_A_G | 11 | 42670351 | G | A | 0.0917105 | -0.105252 | 0.0234961 | 8.90E-06 | 0.99886 | intergenic_region | CTD-2572N17.1:CTD-2537L20.1 |
| rs79933451 | 11:42670362_T_C | 11 | 42670362 | C | T | 0.0917105 | -0.105252 | 0.0234961 | 8.90E-06 | 0.99886 | intergenic_region | CTD-2572N17.1:CTD-2537L20.1 |
| rs75794084 | 11:42670752_C_A | 11 | 42670752 | A | C | 0.0917105 | -0.105257 | 0.023496 | 8.90E-06 | 0.998875 | intergenic_region | CTD-2572N17.1:CTD-2537L20.1 |
| rs67606735 | 2:119634182_C_T | 2 | 119634182 | T | C | 0.429267 | 0.0617176 | 0.0140489 | 9.00E-06 | 0.953195 | intergenic_region | EN1:MARCO |
| rs12546255 | 8:26997387_C_T | 8 | 26997387 | T | C | 0.014045 | 0.254416 | 0.0578794 | 9.00E-06 | 0.992879 | intergenic_region | RP11-521M14.2:AC090150.1 |
| rs78987888 | 11:42666168_G_A | 11 | 42666168 | A | G | 0.0917096 | -0.105195 | 0.023498 | 9.00E-06 | 0.998729 | intergenic_region | CTD-2572N17.1:CTD-2537L20.1 |
| rs80040200 | 11:42666697_G_A | 11 | 42666697 | A | G | 0.0917096 | -0.1052 | 0.0234979 | 9.00E-06 | 0.998745 | intergenic_region | CTD-2572N17.1:CTD-2537L20.1 |
| rs66498268 | 2:119634199_C_G | 2 | 119634199 | G | C | 0.429258 | 0.0617215 | 0.014051 | 9.10E-06 | 0.952847 | intergenic_region | EN1:MARCO |
| rs187035548 | 3:112994258_C_T | 3 | 112994258 | T | C | 0.020737 | 0.212623 | 0.0477323 | 9.20E-06 | 0.96369 | intron_variant | BOC |
| rs17132259 | 4:2146436_C_T | 4 | 2146436 | T | C | 0.216833 | 0.0727078 | 0.0163827 | 9.20E-06 | 1 | downstream_gene_variant | RP11-317B7.3 |
| rs1466283422 | 7:149734839_CCACCCTCGCGGGGGG_C | 7 | 149734839 | C | CCACCCTCGCGGGGGG | 0.018828 | -0.337158 | 0.0759572 | 9.20E-06 | 0.45007 | intergenic_region | RP11-728K20.2:RP11-545G3.1 |
| rs544568560;rs766640583;rs34755099;rs397848958;rs35193525;rs762808646;rs771746224 | 11:29249454_ATTT_ATTTT | 11 | 29249454 | ATTTT | ATTT | 0.0338828 | 0.18438 | 0.0422378 | 9.20E-06 | 0.792634 | intron_variant | RP11-466I1.1 |
| rs755721733;rs34204350;rs768904197;rs202228550;rs571906738;rs761952192;rs869041217;rs869106091;rs869196555 | 11:86777640_CAAAAA_CAAA | 11 | 86777640 | CAAA | CAAAAA | 0.216294 | 0.0745037 | 0.0166929 | 9.20E-06 | 0.955428 | intron_variant | TMEM135 |
| rs11627664 | 14:103485147_A_G | 14 | 103485147 | G | A | 0.116129 | -0.0952527 | 0.0210802 | 9.20E-06 | 1 | intron_variant | CDC42BPB |
| rs577811837 | 2:123629192_A_C | 2 | 123629192 | C | A | 0.0104675 | -0.318646 | 0.0722549 | 9.30E-06 | 0.852524 | intergenic_region | AC018737.3:AC062020.2 |
| rs4604871 | 11:42663943_C_A | 11 | 42663943 | A | C | 0.0949244 | -0.104382 | 0.0233621 | 9.30E-06 | 0.976381 | intergenic_region | CTD-2572N17.1:CTD-2537L20.1 |
| rs201780102;rs531867659 | 11:42671506_A_AAAG | 11 | 42671506 | AAAG | A | 0.0905249 | -0.105896 | 0.0236755 | 9.40E-06 | 0.993693 | intergenic_region | CTD-2572N17.1:CTD-2537L20.1 |
| rs11234993 | 11:86863728_G_A | 11 | 86863728 | A | G | 0.150542 | 0.0841417 | 0.0189155 | 9.40E-06 | 0.998122 | intron_variant | TMEM135 |
| rs74742296 | 1:22684576_A_AG | 1 | 22684576 | AG | A | 0.407827 | -0.0620317 | 0.0137594 | 9.50E-06 | 0.99429 | intergenic_region | RP11-415K20.1:RP11-415K20.2 |
| rs2282764 | 4:2255063_A_G | 4 | 2255063 | G | A | 0.359506 | 0.0625597 | 0.0140519 | 9.50E-06 | 0.996506 | upstream_gene_variant | MIR4800 |
| rs4521104 | 2:137445095_G_C | 2 | 137445095 | C | G | 0.268989 | 0.066425 | 0.0151972 | 9.60E-06 | 0.999601 | intergenic_region | AC092786.1:THSD7B |
| rs200529050;rs545034750 | 11:42682722_T_TTTGTTG | 11 | 42682722 | TTTGTTG | T | 0.136947 | -0.0874722 | 0.0196566 | 9.60E-06 | 0.997203 | intergenic_region | CTD-2572N17.1:CTD-2537L20.1 |
| rs868537992 | 16:17421218_CTTTTTTTTTTTTT_CTTTTTTTTTTTT | 16 | 17421218 | CTTTTTTTTTTTT | CTTTTTTTTTTTTT | 0.219829 | -0.0826393 | 0.0184343 | 9.60E-06 | 0.791355 | intron_variant | XYLT1 |
| rs867199289 | 6:129521249_GTGTACACGTATATATATGTGTATATATATACGTGTACACATATACACGTGTATATATA_<DEL> | 6 | 129521249 | <DEL> | GTGTACACGTATATATATGTGTATATATATACGTGTACACATATACACGTGTATATATA | 0.176623 | 0.106551 | 0.0246627 | 9.80E-06 | 0.525316 | intron_variant | LAMA2 |

Variants were filtered based on minor allele frequency (≥ 0.01) and INFO score (≥ 0.4).

Abbreviations: BP, base-pair position (GRCh37); CHR, chromosome; EA, effect allele; EAF, effect allele frequency; INFO, imputation INFO score; NEA, non-effect allele; SE, standard error.

**Supplementary Table S4. Discriminative capacities of candidate polygenic score (PGS) models based on their correlation with T-score.**

| Derivation strategy | Tuning parameter | Number of variants | Spearman's ρ |
|---|---|---|---|
| P + T | Pr = 1, $R^2$ = 0.2 | 617,404 | 0.0835 |
| P + T | Pr = 1, $R^2$ = 0.4 | 905,433 | 0.0965 |
| P + T | Pr = 1, $R^2$ = 0.6 | 1,203,986 | 0.1017 |
| P + T | Pr = 1, $R^2$ = 0.8 | 1,603,859 | 0.1087 |
| P + T | Pr = 0.5, $R^2$ = 0.2 | 397,089 | 0.0801 |
| P + T | Pr = 0.5, $R^2$ = 0.4 | 544,129 | 0.0947 |
| P + T | Pr = 0.5, $R^2$ = 0.6 | 686,721 | 0.0986 |
| P + T | Pr = 0.5, $R^2$ = 0.8 | 873,689 | 0.1061 |
| P + T | Pr = 0.05, $R^2$ = 0.2 | 57,102 | 0.0803 |
| P + T | Pr = 0.05, $R^2$ = 0.4 | 68,451 | 0.0836 |
| P + T | Pr = 0.05, $R^2$ = 0.6 | 80,356 | 0.0896 |
| P + T | Pr = 0.05, $R^2$ = 0.8 | 97,676 | 0.0995 |
| P + T | Pr = $5 \times 10^{-4}$, $R^2$ = 0.2 | 917 | 0.0434 |
| P + T | Pr = $5 \times 10^{-4}$, $R^2$ = 0.4 | 981 | 0.0407 |
| P + T | Pr = $5 \times 10^{-4}$, $R^2$ = 0.6 | 1,106 | 0.0531 |
| P + T | Pr = $5 \times 10^{-4}$, $R^2$ = 0.8 | 1,287 | 0.0537 |
| P + T | Pr = $5 \times 10^{-6}$, $R^2$ = 0.2 | 19 | 0.1259 |
| P + T | Pr = $5 \times 10^{-6}$, $R^2$ = 0.4 | 21 | 0.1222 |
| P + T | Pr = $5 \times 10^{-6}$, $R^2$ = 0.6 | 26 | 0.1105 |
| P + T | Pr = $5 \times 10^{-6}$, $R^2$ = 0.8 | 33 | 0.0879 |
| P + T | Pr = $5 \times 10^{-8}$, $R^2$ = 0.2 | 3 | 0.1300 |
| P + T | Pr = $5 \times 10^{-8}$, $R^2$ = 0.4 | 3 | 0.1300 |
| P + T | Pr = $5 \times 10^{-8}$, $R^2$ = 0.6 | 3 | 0.1300 |
| P + T | Pr = $5 \times 10^{-8}$, $R^2$ = 0.8 | 4 | 0.1260 |
| LDpred | ρ = 0.001 | 5,495,042 | 0.1120 |
| LDpred | ρ = 0.003 | 5,495,042 | 0.1132 |
| LDpred | ρ = 0.01 | 5,495,042 | 0.1078 |
| LDpred | ρ = 0.03 | 5,495,042 | 0.1061 |
| LDpred | ρ = 0.1 | 5,495,042 | 0.1069 |
| LDpred | ρ = 0.3 | 5,495,042 | 0.1061 |
| LDpred | ρ = 1 | 5,495,042 | 0.1061 |
| PRS-CS | 1.0000 | 1,012,314 | 0.0717 |
| PRS-CS | 0.0100 | 1,012,314 | 0.1039 |
| PRS-CS | $1 \times 10^{-4}$ | 1,012,314 | 0.1376 |
| PRS-CS | $1 \times 10^{-6}$ | 1,012,314 | 0.1356 |
| PRS-CS | $1 \times 10^{-8}$ | 1,012,314 | 0.0568 |
| PRS-CS | auto | 1,012,314 | 0.1465 |

*Abbreviations: LD* , linkage disequilibrium; *PRS-CS,* polygenic risk score-continuous shrinkage, P+T, pruning and thresholding method.

**Supplementary Table S5. Correlation between PGS and age- and sex-adjusted T-scores in each dataset.**

| | | Estimate | 95%CI | | *P* value | Adjusted $R^2$ |
|---|---|---|---|---|---|---|
| | | | lower | upper | | |
| Model selection cohort | TMM9K-JPANEO | 0.126 | 0.08 | 0.17 | **7.11E-08** | 0.198 |
| Validation cohort I | TMM9K-JPAv2 | 0.090 | 0.07 | 0.11 | **1.19E-15** | 0.237 |
| Validation cohort II | TMM18K-JPANEO | 0.098 | 0.04 | 0.16 | **0.001** | 0.204 |

Bold indicates statistical significance ($P < 0.05$).

Abbreviations: *CI*, confidence interval; *JPANEO*, Japonica array NEO; *JPAv2*, Japonica array version 2; *TMM*, Tohoku Medical Megabank.

**Supplementary Table S6. Characteristics of osteoporosis cases and controls in each validation cohort at the baseline.**

| | Validation cohort I (TMM9K-JPAv2, N=7,609) | | | Validation cohort II (TMM18K-JPANEO, N=1,061) | | |
|---|---|---|---|---|---|---|
| | Control (N=6,323) | Case (N=1,286) | *P* value | Control (N=828) | Case (N=233) | *P* value |
| Normalized PGS [a] | 0.017 ± 1.000 | -0.085 ± 0.998 | **8.72E-04** | 0.026 ± 1.005 | -0.093 ± 0.978 | 0.101 |
| Women, N (%) | 4104 (64.9%) | 963 (74.9%) | **4.67E-12** | 581 (70.2%) | 152 (65.2%) | 0.150 |
| Postmenopause, N (% of women) [b] | 2444 (59.6%) | 851 (88.4%) | **6.64E-64** | 383 (65.9%) | 128 (84.2%) | **1.25E-05** |
| Age (years) [a] | 54.9 ± 13.1 | 64.2 ± 10.8 | **2.20E-140** | 55.4 ± 14.0 | 62.7 ± 9.4 | **9.72E-20** |
| T-score [a] | -1.17 ± 0.99 | -2.87 ± 0.31 | **< 1.00E-300** | -1.26 ± 0.96 | -2.84 ± 0.29 | **1.73E-220** |
| Height (cm) [a] | 159.7 ± 8.3 | 156.1 ± 8.2 | **7.02E-43** | 158.8 ± 7.9 | 158.3 ± 8.0 | 0.398 |
| Weight (kg) [a] | 59.8 ± 11.3 | 56.4 ± 10.7 | **2.42E-24** | 58.6 ± 10.8 | 59.4 ± 12.3 | 0.323 |
| BMI ($kg/m^2$) [a] | 23.4 ± 3.6 | 23.1 ± 3.5 | **0.004** | 23.2 ± 3.6 | 23.6 ± 4.0 | 0.108 |
| Fracture, N (%) [c] | 429 (6.8%) | 121 (9.4%) | **9.24E-04** | 57 (6.9%) | 16 (6.9%) | 0.993 |
| Obesity, N (%) [d] | 1770 (28.0%) | 343 (26.7%) | 0.335 | 226 (27.3%) | 74 (31.8%) | 0.181 |
| Smoking, N (%) [e] | 2455 (38.8%) | 364 (28.3%) | **1.06E-12** | 304 (36.7%) | 81 (34.8%) | 0.584 |
| High alcohol intake, N (%) [f] | 979 (15.5%) | 113 (8.8%) | **4.27E-10** | 119 (14.4%) | 30 (12.9%) | 0.561 |
| Physical inactivity, N (%) [g] | 4081 (64.5%) | 704 (54.7%) | **3.35E-11** | 443 (53.5%) | 111 (47.6%) | 0.113 |

*P* values were derived from *t*-test for continuous variables and chi-square test for categorical variables. Bold indicates statistical significance ($P < 0.05$).

Abbreviations: *BMI*, body mass index; *JPANEO*, Japonica array NEO; *JPAv2*, Japonica array version 2; *PGS*, polygenic score; *TMM*, Tohoku Medical Megabank.

a, Mean ± standard deviation.

b, Postmenopause women who reported that their menstruation had stopped at the baseline survey.

c, Fracture was defined based on self-reported questionnaire data and included fractures of the lumbar spine, femur, wrist, or humerus at the baseline survey.

d, Obesity was defined as a body mass index (BMI) ≥25 kg/m² at the baseline survey.

e, Participants who had smoked more than 100 cigarettes in total were classified as smokers at the baseline survey.

f, Participants who consumed ≥30 g of alcohol daily were defined as “high alcohol intake” at the baseline survey.

g, Participants who engaged in leisure-time exercise less than three times per week were categorized as the “physical inactivity” group at the baseline survey.

**Supplementary Table S7. AUCs for models including PGS and age and sex in each validation cohort.**

| Model | Validation cohort Ⅰ (TMM9K-JPAv2, N=7,609) | | | | | Validation cohort Ⅱ (TMM18K-JPANEO, N=1,061) | | | | |
|---|---|---|---|---|---|---|---|---|---|---|
| | AUC | 95%CI | | DeLong's test *P* value | | AUC | 95%CI | | DeLong's test *P* value | |
| | | lower | upper | vs PGS | vs Age + Sex | | lower | upper | vs PGS | vs Age + Sex |
| PGS | 0.528 | 0.511 | 0.545 | – | – | 0.523 | 0.481 | 0.565 | – | – |
| Age + Sex | 0.726 | 0.711 | 0.741 | **1.64E-62** | – | 0.659 | 0.622 | 0.696 | **4.10E-06** | – |
| PGS + Age + Sex | 0.728 | 0.713 | 0.743 | **2.29E-72** | 0.220 | 0.664 | 0.627 | 0.701 | **1.95E-08** | 0.464 |

Bold indicates statistical significance ($P < 0.05$).

Abbreviations: *AUC* , area under the curve; *CI* , confidence interval; *JPANEO* , Japonica array NEO; *JPAv2* , Japonica array

**Supplementary Table S8. Multivariable logistic regression analyses of factors associated with prevalent osteoporosis at baseline.**

| | Variables | OR | 95%CI | | *P* value | Cochran's Q | $I^2$ (%) | $p_{het}$ |
|---|---|---|---|---|---|---|---|---|
| | | | lower | upper | | | | |
| Validation cohort I (TMM9K-JPAv2, N=7,609) | PGS (normalized) | 0.88 | 0.83 | 0.94 | **1.36E-04** | – | – | – |
| | Age | 1.08 | 1.07 | 1.08 | **2.79E-106** | – | – | – |
| | Female | 2.09 | 1.75 | 2.50 | **5.67E-16** | – | – | – |
| | Obesity [a] | 0.88 | 0.77 | 1.02 | 0.090 | – | – | – |
| | Smoking [b] | 1.06 | 0.90 | 1.25 | 0.465 | – | – | – |
| | High alcohol intake [c] | 0.82 | 0.65 | 1.02 | 0.083 | – | – | – |
| | Physical inactivity [d] | 1.12 | 0.98 | 1.28 | 0.103 | – | – | – |
| Validation cohort II (TMM18K-JPANEO, N=1,061) | PGS (normalized) | 0.87 | 0.75 | 1.02 | 0.081 | – | – | – |
| | Age | 1.06 | 1.04 | 1.07 | **1.38E-11** | – | – | – |
| | Female | 0.97 | 0.65 | 1.46 | 0.884 | – | – | – |
| | Obesity [a] | 1.16 | 0.83 | 1.60 | 0.376 | – | – | – |
| | Smoking [b] | 1.05 | 0.70 | 1.56 | 0.811 | – | – | – |
| | High alcohol intake [c] | 0.82 | 0.50 | 1.34 | 0.439 | – | – | – |
| | Physical inactivity [d] | 1.11 | 0.81 | 1.52 | 0.506 | – | – | – |
| Fixed-effect meta-analysis | PGS (normalized) | 0.88 | 0.83 | 0.94 | **2.77E-05** | 0.02 | 0.0 | 0.878 |
| | Age | 1.07 | 1.07 | 1.08 | **3.03E-115** | 4.26 | 76.5 | **0.039** |
| | Female | 1.84 | 1.57 | 2.17 | **2.02E-13** | 11.56 | 91.4 | **0.001** |
| | Obesity [a] | 0.92 | 0.81 | 1.05 | 0.231 | 2.21 | 54.8 | 0.137 |
| | Smoking [b] | 1.06 | 0.91 | 1.24 | 0.444 | 0.00 | 0.0 | 0.954 |
| | High alcohol intake [c] | 0.82 | 0.66 | 1.01 | 0.058 | 0.00 | 0.0 | 0.977 |
| | Physical inactivity [d] | 1.12 | 0.99 | 1.26 | 0.079 | 0.00 | 0.0 | 0.976 |
| Random-effects meta-analysis | PGS (normalized) | 0.88 | 0.83 | 0.94 | **2.77E-05** | – | – | – |
| | Age | 1.07 | 1.05 | 1.09 | **2.41E-13** | – | – | – |
| | Female | 1.46 | 0.69 | 3.09 | 0.326 | – | – | – |
| | Obesity [a] | 0.97 | 0.75 | 1.25 | 0.818 | – | – | – |
| | Smoking [b] | 1.06 | 0.91 | 1.24 | 0.444 | – | – | – |
| | High alcohol intake [c] | 0.82 | 0.66 | 1.01 | 0.058 | – | – | – |
| | Physical inactivity [d] | 1.12 | 0.99 | 1.26 | 0.079 | – | – | – |

Multivariable logistic regression models were used to estimate ORs for prevalent osteoporosis at baseline, including PGS, age, sex, obesity, smoking, high alcohol intake, and physical inactivity as covariates. Cohort-specific estimates were pooled using a fixed-effect meta-analysis. Cochran's Q, $I^2$, and Phet summarise between-cohort heterogeneity. Bold indicates statistical significance ($P< 0.05$).

Abbreviations: *CI*, confidence interval; *$I^2$*, percentage of variation due to heterogeneity; *JPANEO*, Japonica Array NEO; *JPAv2*, Japonica Array version 2; OR, odds ratio; PGS, polygenic score; *Phet*, p value for heterogeneity; *TMM*, Tohoku Medical Megabank.

a, Obesity was defined as a body mass index (BMI) ≥25 kg/m² at the baseline survey.

b, Participants who had smoked more than 100 cigarettes in total were classified as smokers at the baseline survey.

c, Participants who consumed ≥30 g of alcohol daily were defined as “high alcohol intake” at the baseline survey.

d, Participants who engaged in leisure-time exercise less than three times per week were categorized as the “physical inactivity” group at the baseline survey.

**Supplementary Table S9. Characteristics of participants in the validation cohorts according to PGS quintiles.**

| | Validation dataset I (TMM9K-JPAv2, N=7,609) | | | | | | Validation dataset II (TMM18K-JPANEO, N=1,061) | | | | | |
|---|---|---|---|---|---|---|---|---|---|---|---|---|
| | PGS quintile | | | | | *P* value | PGS quintile | | | | | *P* value |
| | Q1 | Q2 | Q3 | Q4 | Q5 | | Q1 | Q2 | Q3 | Q4 | Q5 | |
| Normalized PGS [a] | -1.41 ± 0.48 | -0.53 ± 0.17 | 0.01 ± 0.14 | 0.53 ± 0.17 | 1.39 ± 0.46 | – | -1.42 ± 0.48 | -0.50 ± 0.18 | 0.03 ± 0.14 | 0.53 ± 0.17 | 1.37 ± 0.52 | – |
| Subjects, N | 1522 | 1522 | 1521 | 1522 | 1522 | – | 213 | 212 | 212 | 212 | 212 | – |
| Women, N (%) | 1,000 (65.7) | 1,028 (67.5) | 1,005 (66.1) | 1,007 (66.2) | 1,027 (67.5) | 0.735 | 155 (72.8) | 138 (65.1) | 150 (70.8) | 142 (67.0) | 148 (69.8) | 0.448 |
| Postmenopause, N (% of women) [b] | 656 (65.6) | 644 (62.6) | 640 (63.7) | 664 (65.9) | 691 (67.3) | 0.445 | 103 (66.5) | 99 (71.7) | 100 (66.7) | 104 (73.2) | 105 (70.9) | 0.798 |
| Baseline Age [a] | 56.5 ± 13.4 | 56.0 ± 13.5 | 56.1 ± 13.1 | 57.1 ± 13.0 | 56.7 ± 13.0 | 0.126 | 56.4 ± 13.5 | 57.6 ± 13.2 | 57.2 ± 12.5 | 56.1 ± 14.2 | 57.6 ± 13.7 | 0.767 |
| Baseline T-score [a] | -1.56 ± 1.09 | -1.52 ± 1.07 | -1.46 ± 1.10 | -1.46 ± 1.13 | -1.32 ± 1.17 | **1.68E-07** | -1.68 ± 1.00 | -1.64 ± 1.03 | -1.66 ± 1.03 | -1.53 ± 1.19 | -1.54 ± 1.14 | 0.839 |
| Height (cm)[a] | 159.2 ± 8.5 | 159.3 ± 8.3 | 159.5 ± 8.3 | 158.7 ± 8.4 | 158.6 ± 8.4 | **0.013** | 159.1 ± 8.4 | 158.8 ± 7.6 | 158.8 ± 7.9 | 158.6 ± 7.9 | 158.4 ± 7.9 | 0.942 |
| Weight (kg) [a] | 59.6 ± 11.4 | 58.9 ± 11.1 | 59.4 ± 11.1 | 59.1 ± 11.4 | 59.0 ± 11.5 | 0.314 | 59.1 ± 11.9 | 59.6 ± 9.9 | 58.8 ± 11.0 | 58.6 ± 12.1 | 57.8 ± 11.0 | 0.260 |
| BMI (kg/m$^2$) [a] | 23.4 ± 3.7 | 23.1 ± 3.4 | 23.3 ± 3.5 | 23.4 ± 3.5 | 23.4 ± 3.7 | 0.188 | 23.3 ± 3.7 | 23.6 ± 3.4 | 23.3 ± 3.7 | 23.2 ± 3.9 | 22.9 ± 3.5 | 0.310 |
| Osteoporosis, N (%) [c] | 289 (19.0) | 273 (17.9) | 240 (15.8) | 263 (17.3) | 221 (14.5) | **0.009** | 49 (23.0) | 46 (21.7) | 49 (23.1) | 45 (21.2) | 44 (20.8) | 0.968 |
| Fracture, N (%) [d] | 126 (8.3) | 97 (6.4) | 111 (7.3) | 101 (6.6) | 115 (7.6) | 0.266 | 9 (4.2) | 22 (10.4) | 15 (7.1) | 8 (3.8) | 19 (9.0) | **0.026** |
| Obesity, N (%) [e] | 451 (29.6) | 385 (25.3) | 399 (26.2) | 432 (28.4) | 446 (29.3) | **0.025** | 57 (26.8) | 65 (30.7) | 61 (28.8) | 59 (27.8) | 58 (27.4) | 0.914 |
| Smoking, N (%) [f] | 578 (38.0) | 575 (37.8) | 572 (37.6) | 570 (37.5) | 524 (34.4) | 0.223 | 69 (32.4) | 81 (38.2) | 78 (36.8) | 85 (40.1) | 72 (34.0) | 0.465 |
| High alcohol intake, N (%) [g] | 225 (14.8) | 218 (14.3) | 225 (14.8) | 221 (14.5) | 203 (13.3) | 0.776 | 22 (10.3) | 29 (13.7) | 34 (16.0) | 34 (16.0) | 30 (14.2) | 0.426 |
| Physical inactivity, N (%) [h] | 943 (62.0) | 947 (62.2) | 1001 (65.8) | 928 (61.0) | 966 (63.5) | 0.060 | 127 (59.6) | 99 (46.7) | 104 (49.1) | 122 (57.6) | 102 (48.1) | **0.018** |

*P* values were derived from Kruskal–Wallis for continuous variables and Pearson's chi-square for categorical variables across quintiles (Q1–Q5) (two-sided). Bold indicates statistical significance ($P < 0.05$).

Abbreviations: *BMI*, body mass index; *JPANEO*, Japonica Array NEO; *JPAv2*, Japonica Array version 2; *PGS,* polygenic score; *TMM*, Tohoku Medical Megabank. Q indicates quintile.

a, Mean ± standard deviation.

b, Postmenopause women who reported that their menstruation had stopped at the baseline survey. Percentages for postmenopause are calculated among women only.

c, Osteoporosis was defined as a QUS T-score of −2.5 SD or lower at the baseline survey.

d, Fracture was defined based on self-reported questionnaire data and included fractures of the lumbar spine, femur, wrist, or humerus at the baseline survey.

e, Obesity was defined as a body mass index (BMI) ≥25 kg/m² at the baseline survey.

f, Participants who had smoked more than 100 cigarettes in total were classified as smokers at the baseline survey.

g, Participants who consumed ≥30 g of alcohol daily were defined as "high alcohol intake" at the baseline survey.

h, Participants who engaged in leisure-time exercise less than three times per week were categorized as the "physical inactivity" group at the baseline survey.

**Supplementary Table S10. Prevalence and ORs of osteoporosis at baseline by age-stratified genetic risk groups based on PGS quintiles.**

| | | Age (year) | Genetic risk [PGS quintile] | Number of subjects (n) | Number of cases (n) | Prevalence (%) | OR | 95%CI lower | 95%CI upper | *P* value | *P* for trend | Cochran's Q | $I^2$ (%) | $p_{het}$ |
|---|---|---|---|---|---|---|---|---|---|---|---|---|---|---|
| Total | Validation cohort I (TMM9K-JPAv2, N=7,609) | <65 | High [Q1] | 1,052 | 131 | 12.5 | 1.03 | 0.83 | 1.28 | 0.772 | | – | – | – |
| | | | Intermediate [Q2 - Q4] | 3,212 | 390 | 12.1 | 1.00 (reference) | – | – | – | **0.013** | – | – | – |
| | | | Low [Q5] | 1,056 | 99 | 9.4 | 0.72 | 0.56 | 0.91 | **0.006** | | – | – | – |
| | | ≥65 | High [Q1] | 470 | 158 | 33.6 | 1.34 | 1.06 | 1.69 | **0.016** | | – | – | – |
| | | | Intermediate [Q2 - Q4] | 1,353 | 386 | 28.5 | 1.00 (reference) | – | – | – | **0.006** | – | – | – |
| | | | Low [Q5] | 466 | 122 | 26.2 | 0.89 | 0.69 | 1.14 | 0.363 | | – | – | – |
| | Validation cohort II (TMM18K-JPANEO, N=1,061) | <65 | High [Q1] | 141 | 25 | 17.7 | 1.08 | 0.63 | 1.79 | 0.772 | | – | – | – |
| | | | Intermediate [Q2 - Q4] | 407 | 68 | 16.7 | 1.00 (reference) | – | – | – | 0.482 | – | – | – |
| | | | Low [Q5] | 137 | 21 | 15.3 | 0.85 | 0.49 | 1.45 | 0.570 | | – | – | – |
| | | ≥65 | High [Q1] | 72 | 24 | 33.3 | 1.09 | 0.61 | 1.91 | 0.767 | | – | – | – |
| | | | Intermediate [Q2 - Q4] | 229 | 72 | 31.4 | 1.00 (reference) | – | – | – | 0.673 | – | – | – |
| | | | Low [Q5] | 75 | 23 | 30.7 | 0.94 | 0.52 | 1.64 | 0.822 | | – | – | – |
| | Fixed-effect meta-analysis | <65 | High [Q1] | – | – | – | 1.04 | 0.85 | 1.27 | 0.706 | | 0.02 | 0.0 | 0.88 |
| | | | Intermediate [Q2 - Q4] | – | – | – | 1.00 (reference) | – | – | – | – | – | – | – |
| | | | Low [Q5] | – | – | – | 0.74 | 0.59 | 0.92 | **0.006** | | 0.32 | 0.0 | 0.57 |
| | | ≥65 | High [Q1] | – | – | – | 1.30 | 1.04 | 1.62 | **0.019** | | 0.43 | 0.0 | 0.51 |
| | | | Intermediate [Q2 - Q4] | – | – | – | 1.00 (reference) | – | – | – | – | – | – | – |
| | | | Low [Q5] | – | – | – | 0.90 | 0.72 | 1.13 | 0.356 | | 0.03 | 0.0 | 0.87 |
| | Random-effects meta-analysis | <65 | High [Q1] | – | – | – | 1.04 | 0.85 | 1.27 | 0.706 | | – | – | – |
| | | | Intermediate [Q2 - Q4] | – | – | – | 1.00 (reference) | – | – | – | – | – | – | – |
| | | | Low [Q5] | – | – | – | 0.74 | 0.59 | 0.92 | **0.006** | | – | – | – |
| | | ≥65 | High [Q1] | – | – | – | 1.30 | 1.04 | 1.62 | **0.019** | | – | – | – |
| | | | Intermediate [Q2 - Q4] | – | – | – | 1.00 (reference) | – | – | – | – | – | – | – |
| | | | Low [Q5] | – | – | – | 0.90 | 0.72 | 1.13 | 0.356 | | – | – | – |
| Men | Validation cohort I (TMM9K-JPAv2, N=7,609) | <65 | High [Q1] | 307 | 29 | 9.4 | 0.99 | 0.63 | 1.53 | 0.977 | | – | – | – |
| | | | Intermediate [Q2 - Q4] | 937 | 91 | 9.7 | 1.00 (reference) | – | – | – | 0.301 | – | – | – |
| | | | Low [Q5] | 308 | 22 | 7.1 | 0.72 | 0.43 | 1.16 | 0.193 | | – | – | – |
| | | ≥65 | High [Q1] | 215 | 43 | 20.0 | 1.13 | 0.75 | 1.67 | 0.556 | | – | – | – |
| | | | Intermediate [Q2 - Q4] | 588 | 109 | 18.5 | 1.00 (reference) | – | – | – | 0.385 | – | – | – |
| | | | Low [Q5] | 187 | 29 | 15.5 | 0.90 | 0.56 | 1.40 | 0.635 | | – | – | – |
| | Validation cohort II (TMM18K-JPANEO, N=1,061) | <65 | High [Q1] | 35 | 7 | 20.0 | 0.94 | 0.34 | 2.37 | 0.893 | | – | – | – |
| | | | Intermediate [Q2 - Q4] | 112 | 23 | 20.5 | 1.00 (reference) | – | – | – | 0.672 | – | – | – |
| | | | Low [Q5] | 32 | 6 | 18.8 | 0.71 | 0.24 | 1.88 | 0.511 | | – | – | – |
| | | ≥65 | High [Q1] | 23 | 7 | 30.4 | 0.97 | 0.34 | 2.55 | 0.952 | | – | – | – |
| | | | Intermediate [Q2 - Q4] | 94 | 29 | 30.9 | 1.00 (reference) | – | – | – | 0.829 | – | – | – |
| | | | Low [Q5] | 32 | 9 | 28.1 | 0.87 | 0.34 | 2.06 | 0.753 | | – | – | – |
| | Fixed-effect meta-analysis | <65 | High [Q1] | – | – | – | 0.98 | 0.66 | 1.47 | 0.936 | | 0.01 | 0.0 | 0.91 |
| | | | Intermediate [Q2 - Q4] | – | – | – | 1.00 (reference) | – | – | – | – | – | – | – |
| | | | Low [Q5] | – | – | – | 0.72 | 0.46 | 1.12 | 0.147 | | 0.00 | 0.0 | 0.97 |
| | | ≥65 | High [Q1] | – | – | – | 1.10 | 0.76 | 1.60 | 0.600 | | 0.07 | 0.0 | 0.78 |
| | | | Intermediate [Q2 - Q4] | – | – | – | 1.00 (reference) | – | – | – | – | – | – | – |
| | | | Low [Q5] | – | – | – | 0.89 | 0.59 | 1.34 | 0.573 | | 0.00 | 0.0 | 0.95 |
| | Random-effects meta-analysis | <65 | High [Q1] | – | – | – | 0.98 | 0.66 | 1.47 | 0.936 | | – | – | – |
| | | | Intermediate [Q2 - Q4] | – | – | – | 1.00 (reference) | – | – | – | – | – | – | – |
| | | | Low [Q5] | – | – | – | 0.72 | 0.46 | 1.12 | 0.147 | | – | – | – |
| | | ≥65 | High [Q1] | – | – | – | 1.10 | 0.76 | 1.60 | 0.600 | | – | – | – |
| | | | Intermediate [Q2 - Q4] | – | – | – | 1.00 (reference) | – | – | – | – | – | – | – |
| | | | Low [Q5] | – | – | – | 0.89 | 0.59 | 1.34 | 0.573 | | – | – | – |
| Women | Validation cohort I (TMM9K-JPAv2, N=7,609) | <65 | High [Q1] | 745 | 102 | 13.7 | 1.04 | 0.81 | 1.33 | 0.763 | | – | – | – |
| | | | Intermediate [Q2 - Q4] | 2,275 | 299 | 13.1 | 1.00 (reference) | – | – | – | **0.022** | – | – | – |
| | | | Low [Q5] | 748 | 77 | 10.3 | 0.71 | 0.54 | 0.92 | **0.013** | | – | – | – |
| | | ≥65 | High [Q1] | 255 | 115 | 45.1 | 1.47 | 1.09 | 1.98 | **0.010** | | – | – | – |
| | | | Intermediate [Q2 - Q4] | 765 | 277 | 36.2 | 1.00 (reference) | – | – | – | **0.006** | – | – | – |
| | | | Low [Q5] | 279 | 93 | 33.3 | 0.89 | 0.66 | 1.19 | 0.439 | | – | – | – |
| | Validation cohort II (TMM18K-JPANEO, N=1,061) | <65 | High [Q1] | 106 | 18 | 17.0 | 1.15 | 0.61 | 2.09 | 0.662 | | – | – | – |
| | | | Intermediate [Q2 - Q4] | 295 | 45 | 15.3 | 1.00 (reference) | – | – | – | 0.586 | – | – | – |
| | | | Low [Q5] | 105 | 15 | 14.3 | 0.93 | 0.47 | 1.74 | 0.823 | | – | – | – |
| | | ≥65 | High [Q1] | 49 | 17 | 34.7 | 1.16 | 0.57 | 2.30 | 0.676 | | – | – | – |
| | | | Intermediate [Q2 - Q4] | 135 | 43 | 31.9 | 1.00 (reference) | – | – | – | 0.705 | – | – | – |
| | | | Low [Q5] | 43 | 14 | 32.6 | 0.98 | 0.46 | 2.04 | 0.968 | | – | – | – |
| | Fixed-effect meta-analysis | <65 | High [Q1] | – | – | – | 1.05 | 0.84 | 1.33 | 0.659 | | 0.08 | 0.0 | 0.77 |
| | | | Intermediate [Q2 - Q4] | – | – | – | 1.00 (reference) | – | – | – | – | – | – | – |
| | | | Low [Q5] | – | – | – | 0.74 | 0.57 | 0.95 | **0.017** | | 0.57 | 0.0 | 0.45 |
| | | ≥65 | High [Q1] | – | – | – | 1.42 | 1.08 | 1.86 | **0.012** | | 0.38 | 0.0 | 0.54 |
| | | | Intermediate [Q2 - Q4] | – | – | – | 1.00 (reference) | – | – | – | – | – | – | – |
| | | | Low [Q5] | – | – | – | 0.90 | 0.68 | 1.19 | 0.464 | | 0.06 | 0.0 | 0.80 |
| | Random-effects meta-analysis | <65 | High [Q1] | – | – | – | 1.05 | 0.84 | 1.33 | 0.659 | | – | – | – |
| | | | Intermediate [Q2 - Q4] | – | – | – | 1.00 (reference) | – | – | – | – | – | – | – |
| | | | Low [Q5] | – | – | – | 0.74 | 0.57 | 0.95 | **0.017** | | – | – | – |
| | | ≥65 | High [Q1] | – | – | – | 1.42 | 1.08 | 1.86 | **0.012** | | – | – | – |
| | | | Intermediate [Q2 - Q4] | – | – | – | 1.00 (reference) | – | – | – | – | – | – | – |
| | | | Low [Q5] | – | – | – | 0.90 | 0.68 | 1.19 | 0.464 | | – | – | – |

Logistic regression models adjusted for sex were used to estimate ORs for prevalent osteoporosis at baseline across six groups defined by age (<65 or ≥65 years) and genetic risk based on PGS quintiles: high (Q1), intermediate (Q2–Q4), and low (Q5). Within each age stratum, the intermediate-risk group (Q2–Q4) served as the reference. Cohort-specific estimates were pooled using fixed-effect and random-effects (DerSimonian–Laird) meta-analyses; fixed-effect estimates were prespecified as primary, with random-effects estimates examined as sensitivity analyses. Cochran's Q, $I^2$, and Phet summarise between-cohort heterogeneity. Bold indicates statistical significance ($P < 0.05$).

Abbreviations: *CI*, confidence interval; $I^2$, percentage of variation due to heterogeneity; *JPANEO*, Japonica Array NEO; *JPAv2*, Japonica Array version 2; *OR*, odds ratio; *Phet*, P value for heterogeneity; *PGS*, polygenic score; *Q*, quintile; *TMM*, Tohoku Medical Megabank.

**Supplementary Table S11. Prevalence and ORs of osteoporosis at baseline by genetic risk groups based on PGS quintiles and modifiable lifestyle factors.**

| Lifestyle factor | | | Genetic risk [quintile] | Number of subjects (n) | Number of cases (n) | Prevalence (%) | OR | 95%CI | | *P* value | *P* for trend | Cochran's Q | $I^2$ (%) | $p_{het}$ |
|---|---|---|---|---|---|---|---|---|---|---|---|---|---|---|
| | | | | | | | | lower | upper | | | | | |
| Obesity [a] | Validation cohort Ⅰ (TMM9K-JPAv2, N=7,609) | No | High [Q1] | 1,071 | 211 | 19.7 | 1.18 | 0.98 | 1.42 | 0.078 | | – | – | – |
| | | | Intermediate [Q2 - Q4] | 3,349 | 576 | 17.2 | 1.00 (reference) | – | – | – | 0.077 | – | – | – |
| | | | Low [Q5] | 1,076 | 156 | 14.5 | 0.78 | 0.64 | 0.95 | **0.014** | | – | – | – |
| | | Yes | High [Q1] | 451 | 78 | 17.3 | 0.97 | 0.74 | 1.28 | 0.853 | | – | – | – |
| | | | Intermediate [Q2 - Q4] | 1,216 | 200 | 16.4 | 0.89 | 0.74 | 1.07 | 0.223 | 0.197 | – | – | – |
| | | | Low [Q5] | 446 | 65 | 14.6 | 0.76 | 0.56 | 1.01 | 0.060 | | – | – | – |
| | Validation cohort Ⅱ (TMM18K-JPANEO, N=1,061) | No | High [Q1] | 156 | 39 | 25.0 | 1.32 | 0.85 | 2.05 | 0.223 | | – | – | – |
| | | | Intermediate [Q2 - Q4] | 451 | 90 | 20.0 | 1.00 (reference) | – | – | – | 0.971 | – | – | – |
| | | | Low [Q5] | 154 | 30 | 19.5 | 0.93 | 0.58 | 1.49 | 0.760 | | – | – | – |
| | | Yes | High [Q1] | 57 | 10 | 17.5 | 0.89 | 0.42 | 1.86 | 0.753 | | – | – | – |
| | | | Intermediate [Q2 - Q4] | 185 | 50 | 27.0 | 1.34 | 0.89 | 2.02 | 0.161 | 0.656 | – | – | – |
| | | | Low [Q5] | 58 | 14 | 24.1 | 1.11 | 0.57 | 2.15 | 0.757 | | – | – | – |
| | Fixed-effect meta-analysis | No | High [Q1] | – | – | – | 1.20 | 1.01 | 1.43 | **0.036** | | 0.20 | 0.0 | 0.66 |
| | | | Intermediate [Q2 - Q4] | – | – | – | 1.00 (reference) | – | – | – | – | – | – | – |
| | | | Low [Q5] | – | – | – | 0.80 | 0.66 | 0.96 | **0.017** | | 0.47 | 0.0 | 0.49 |
| | | Yes | High [Q1] | – | – | – | 0.96 | 0.75 | 1.24 | 0.777 | | 0.05 | 0.0 | 0.82 |
| | | | Intermediate [Q2 - Q4] | – | – | – | 0.96 | 0.81 | 1.13 | 0.596 | – | 3.17 | 68.4 | 0.08 |
| | | | Low [Q5] | – | – | – | 0.80 | 0.62 | 1.05 | 0.109 | | 1.08 | 7.8 | 0.30 |
| | Random-effects meta-analysis | No | High [Q1] | – | – | – | 1.20 | 1.01 | 1.43 | **0.036** | | – | – | – |
| | | | Intermediate [Q2 - Q4] | – | – | – | 1.00 (reference) | – | – | – | – | – | – | – |
| | | | Low [Q5] | – | – | – | 0.80 | 0.66 | 0.96 | **0.017** | | – | – | – |
| | | Yes | High [Q1] | – | – | – | 0.96 | 0.75 | 1.24 | 0.777 | | – | – | – |
| | | | Intermediate [Q2 - Q4] | – | – | – | 1.05 | 0.71 | 1.55 | 0.816 | – | – | – | – |
| | | | Low [Q5] | – | – | – | 0.81 | 0.61 | 1.09 | 0.167 | | – | – | – |
| Smorking [b] | Validation cohort Ⅰ (TMM9K-JPAv2, N=7,609) | No | High [Q1] | 944 | 211 | 22.4 | 1.20 | 1.00 | 1.46 | 0.054 | | – | – | – |
| | | | Intermediate [Q2 - Q4] | 2,848 | 543 | 19.1 | 1.00 (reference) | – | – | – | 0.225 | – | – | – |
| | | | Low [Q5] | 998 | 168 | 16.8 | 0.83 | 0.68 | 1.01 | 0.062 | | – | – | – |
| | | Yes | High [Q1] | 578 | 78 | 13.5 | 1.14 | 0.86 | 1.51 | 0.354 | | – | – | – |
| | | | Intermediate [Q2 - Q4] | 1,717 | 233 | 13.6 | 1.09 | 0.90 | 1.33 | 0.377 | 0.067 | – | – | – |
| | | | Low [Q5] | 524 | 53 | 10.1 | 0.79 | 0.57 | 1.08 | 0.142 | | – | – | – |
| | Validation cohort Ⅱ (TMM18K-JPANEO, N=1,061) | No | High [Q1] | 144 | 32 | 22.2 | 0.97 | 0.61 | 1.55 | 0.895 | | – | – | – |
| | | | Intermediate [Q2 - Q4] | 392 | 90 | 23.0 | 1.00 (reference) | – | – | – | 0.758 | – | – | – |
| | | | Low [Q5] | 140 | 30 | 21.4 | 0.93 | 0.58 | 1.51 | 0.779 | | – | – | – |
| | | Yes | High [Q1] | 69 | 17 | 24.6 | 1.35 | 0.70 | 2.61 | 0.375 | | – | – | – |
| | | | Intermediate [Q2 - Q4] | 244 | 50 | 20.5 | 0.96 | 0.61 | 1.52 | 0.869 | 0.170 | – | – | – |
| | | | Low [Q5] | 72 | 14 | 19.4 | 0.78 | 0.39 | 1.55 | 0.472 | | – | – | – |
| | Fixed-effect meta-analysis | No | High [Q1] | – | – | – | 1.17 | 0.98 | 1.39 | 0.083 | | 0.71 | 0.0 | 0.40 |
| | | | Intermediate [Q2 - Q4] | – | – | – | 1.00 (reference) | – | – | – | – | – | – | – |
| | | | Low [Q5] | – | – | – | 0.84 | 0.70 | 1.01 | 0.067 | | 0.21 | 0.0 | 0.65 |
| | | Yes | High [Q1] | – | – | – | 1.17 | 0.90 | 1.52 | 0.230 | | 0.20 | 0.0 | 0.65 |
| | | | Intermediate [Q2 - Q4] | – | – | – | 1.07 | 0.89 | 1.28 | 0.455 | – | 0.25 | 0.0 | 0.62 |
| | | | Low [Q5] | – | – | – | 0.78 | 0.59 | 1.05 | 0.102 | | 0.00 | 0.0 | 0.97 |
| | Random-effects meta-analysis | No | High [Q1] | – | – | – | 1.17 | 0.98 | 1.39 | 0.083 | | – | – | – |
| | | | Intermediate [Q2 - Q4] | – | – | – | 1.00 (reference) | – | – | – | – | – | – | – |
| | | | Low [Q5] | – | – | – | 0.84 | 0.70 | 1.01 | 0.067 | | – | – | – |
| | | Yes | High [Q1] | – | – | – | 1.17 | 0.90 | 1.52 | 0.230 | | – | – | – |
| | | | Intermediate [Q2 - Q4] | – | – | – | 1.07 | 0.89 | 1.28 | 0.455 | – | – | – | – |
| | | | Low [Q5] | – | – | – | 0.78 | 0.59 | 1.05 | 0.102 | | – | – | – |
| High alcohol intake [c] | Validation cohort Ⅰ (TMM9K-JPAv2, N=7,609) | No | High [Q1] | 1,297 | 263 | 20.3 | 1.17 | 0.99 | 1.38 | 0.063 | | – | – | – |
| | | | Intermediate [Q2 - Q4] | 3,901 | 702 | 18.0 | 1.00 (reference) | – | – | – | 0.134 | – | – | – |
| | | | Low [Q5] | 1,319 | 208 | 15.8 | 0.82 | 0.69 | 0.98 | **0.030** | | – | – | – |
| | | Yes | High [Q1] | 225 | 26 | 11.6 | 0.92 | 0.59 | 1.43 | 0.712 | | – | – | – |
| | | | Intermediate [Q2 - Q4] | 664 | 74 | 11.1 | 0.91 | 0.69 | 1.20 | 0.522 | 0.101 | – | – | – |
| | | | Low [Q5] | 203 | 13 | 6.4 | 0.49 | 0.28 | 0.88 | **0.018** | | – | – | – |
| | Validation cohort Ⅱ (TMM18K-JPANEO, N=1,061) | No | High [Q1] | 191 | 43 | 22.5 | 1.03 | 0.68 | 1.55 | 0.895 | | – | – | – |
| | | | Intermediate [Q2 - Q4] | 539 | 121 | 22.4 | 1.00 (reference) | – | – | – | 0.747 | – | – | – |
| | | | Low [Q5] | 182 | 39 | 21.4 | 0.92 | 0.60 | 1.40 | 0.692 | | – | – | – |
| | | Yes | High [Q1] | 22 | 6 | 27.3 | 1.37 | 0.50 | 3.72 | 0.543 | | – | – | – |
| | | | Intermediate [Q2 - Q4] | 97 | 19 | 19.6 | 0.81 | 0.46 | 1.45 | 0.487 | 0.245 | – | – | – |
| | | | Low [Q5] | 30 | 5 | 16.7 | 0.59 | 0.21 | 1.62 | 0.303 | | – | – | – |
| | Fixed-effect meta-analysis | No | High [Q1] | – | – | – | 1.15 | 0.99 | 1.34 | 0.077 | | 0.34 | 0.0 | 0.56 |
| | | | Intermediate [Q2 - Q4] | – | – | – | 1.00 (reference) | – | – | – | – | – | – | – |
| | | | Low [Q5] | – | – | – | 0.84 | 0.71 | 0.98 | **0.031** | | 0.23 | 0.0 | 0.63 |
| | | Yes | High [Q1] | – | – | – | 0.98 | 0.66 | 1.47 | 0.926 | | 0.50 | 0.0 | 0.48 |
| | | | Intermediate [Q2 - Q4] | – | – | – | 0.89 | 0.70 | 1.15 | 0.381 | – | 0.12 | 0.0 | 0.72 |
| | | | Low [Q5] | – | – | – | 0.51 | 0.31 | 0.85 | **0.010** | | 0.08 | 0.0 | 0.78 |
| | Random-effects meta-analysis | No | High [Q1] | – | – | – | 1.15 | 0.99 | 1.34 | 0.077 | | – | – | – |
| | | | Intermediate [Q2 - Q4] | – | – | – | 1.00 (reference) | – | – | – | – | – | – | – |
| | | | Low [Q5] | – | – | – | 0.84 | 0.71 | 0.98 | **0.031** | | – | – | – |
| | | Yes | High [Q1] | – | – | – | 0.98 | 0.66 | 1.47 | 0.926 | | – | – | – |
| | | | Intermediate [Q2 - Q4] | – | – | – | 0.89 | 0.70 | 1.15 | 0.381 | – | – | – | – |
| | | | Low [Q5] | – | – | – | 0.51 | 0.31 | 0.85 | **0.010** | | – | – | – |
| Physical inactivity [d] | Validation cohort Ⅰ (TMM9K-JPAv2, N=7,609) | No | High [Q1] | 579 | 138 | 23.8 | 1.31 | 1.03 | 1.66 | **0.025** | | – | – | – |
| | | | Intermediate [Q2 - Q4] | 1,689 | 344 | 20.4 | 1.00 (reference) | – | – | – | 0.785 | – | – | – |
| | | | Low [Q5] | 556 | 100 | 18.0 | 0.87 | 0.68 | 1.13 | 0.307 | | – | – | – |
| | | Yes | High [Q1] | 943 | 151 | 16.0 | 1.26 | 1.01 | 1.58 | **0.044** | | – | – | – |
| | | | Intermediate [Q2 - Q4] | 2,876 | 432 | 15.0 | 1.21 | 1.02 | 1.44 | **0.027** | **0.015** | – | – | – |
| | | | Low [Q5] | 966 | 121 | 12.5 | 0.89 | 0.70 | 1.13 | 0.336 | | – | – | – |
| | Validation cohort Ⅱ (TMM18K-JPANEO, N=1,061) | No | High [Q1] | 86 | 27 | 31.4 | 1.67 | 0.97 | 2.88 | 0.064 | | – | – | – |
| | | | Intermediate [Q2 - Q4] | 311 | 68 | 21.9 | 1.00 (reference) | – | – | – | 0.477 | – | – | – |
| | | | Low [Q5] | 110 | 27 | 24.5 | 1.08 | 0.64 | 1.82 | 0.767 | | – | – | – |
| | | Yes | High [Q1] | 127 | 22 | 17.3 | 1.03 | 0.59 | 1.79 | 0.913 | | – | – | – |
| | | | Intermediate [Q2 - Q4] | 325 | 72 | 22.2 | 1.43 | 0.96 | 2.12 | 0.076 | 0.870 | – | – | – |
| | | | Low [Q5] | 102 | 17 | 16.7 | 1.02 | 0.56 | 1.88 | 0.944 | | – | – | – |
| | Fixed-effect meta-analysis | No | High [Q1] | – | – | – | 1.36 | 1.10 | 1.69 | **0.005** | | 0.65 | 0.0 | 0.42 |
| | | | Intermediate [Q2 - Q4] | – | – | – | 1.00 (reference) | – | – | – | – | – | – | – |
| | | | Low [Q5] | – | – | – | 0.91 | 0.72 | 1.15 | 0.433 | | 0.52 | 0.0 | 0.47 |
| | | Yes | High [Q1] | – | – | – | 1.22 | 0.99 | 1.51 | 0.057 | | 0.44 | 0.0 | 0.51 |
| | | | Intermediate [Q2 - Q4] | – | – | – | 1.24 | 1.06 | 1.45 | **0.006** | – | 0.57 | 0.0 | 0.45 |
| | | | Low [Q5] | – | – | – | 0.91 | 0.73 | 1.13 | 0.384 | | 0.17 | 0.0 | 0.68 |
| | Random-effects meta-analysis | No | High [Q1] | – | – | – | 1.36 | 1.10 | 1.69 | **0.005** | | – | – | – |
| | | | Intermediate [Q2 - Q4] | – | – | – | 1.00 (reference) | – | – | – | – | – | – | – |
| | | | Low [Q5] | – | – | – | 0.91 | 0.72 | 1.15 | 0.433 | | – | – | – |
| | | Yes | High [Q1] | – | – | – | 1.22 | 0.99 | 1.51 | 0.057 | | – | – | – |
| | | | Intermediate [Q2 - Q4] | – | – | – | 1.24 | 1.06 | 1.45 | **0.006** | – | – | – | – |
| | | | Low [Q5] | – | – | – | 0.91 | 0.73 | 1.13 | 0.384 | | – | – | – |

Logistic regression models adjusted for age and sex were used to estimate ORs for prevalent osteoporosis at baseline across six groups defined by genetic risk [high (Q1), intermediate (Q2–Q4), and low (Q5)] and each modifiable lifestyle factor (yes/no).
Cohort-specific estimates were pooled using fixed-effect and random-effects (DerSimonian–Laird) meta-analyses; fixed-effect estimates were prespecified as primary, with random-effects estimates examined as sensitivity analyses. Cochran's Q, $I^2$, and Phet summarise between-cohort heterogeneity. Bold indicates statistical significance ($P < 0.05$).
Abbreviations: *CI* , confidence interval; $I^2$, percentage of variation due to heterogeneity; *JPANEO* , Japonica Array NEO; *JPAv2* , Japonica Array version 2; *OR* , odds ratio; *Phet* , P value for heterogeneity; *PGS* , polygenic score; *Q* , quintile; *TMM* , Tohoku Medical Megabank.
a, Obesity was defined as a body mass index (BMI) ≥25 kg/m² at the baseline survey.
b, Participants who had smoked more than 100 cigarettes in total were classified as smokers at the baseline survey.
c, Participants who consumed ≥30 g of alcohol daily were defined as “high alcohol intake” at the baseline survey.
d, Participants who engaged in leisure-time exercise less than three times per week were categorized as the “physical inactivity” group at the baseline survey.

**Supplementary Table S12. Baseline and follow-up characteristics of osteoporosis cases and controls in each follow-up cohort.**

| | Validation cohort I (TMM9K-JPAv2, N=4,811) | | | Validation cohort II (TMM18K-JPANEO, N=1,061) | | |
|---|---|---|---|---|---|---|
| | Control (N=4,369) | Case (N=442) | *P* value | Control (N=465) | Case (N=116) | *P* value |
| Normalized PGS [a] | 0.047 ± 0.989 | -0.194 ± 0.994 | **1.45E-06** | 0.077 ± 0.983 | -0.226 ± 0.997 | **0.004** |
| Women, N (%) | 2,814 (64.4%) | 321 (72.6%) | **5.51E-04** | 311 (66.9%) | 77 (66.4%) | 0.918 |
| Postmenopause, N (% of women) [b] | 1,607 (57.1%) | 262 (81.6%) | **2.25E-17** | 196 (63.0%) | 69 (89.6%) | **7.16E-06** |
| Baseline Age (years) [a] | 54.7 ± 12.3 | 60.5 ± 10.2 | **4.72E-26** | 54.5 ± 13.7 | 62.0 ± 9.3 | **1.66E-11** |
| Follow-up Age (years) [a] | 58.6 ± 12.3 | 64.4 ± 10.3 | **1.09E-25** | 59.1 ± 13.6 | 66.8 ± 9.4 | **9.51E-12** |
| Baseline T-score [a] | -1.09 ± 0.97 | -1.91 ± 0.52 | **1.35E-122** | -1.12 ± 0.95 | -1.79 ± 0.63 | **2.92E-17** |
| Follow-up T-score [a] | -1.17 ± 0.96 | -2.79 ± 0.29 | **< 1.00E-300** | -1.27 ± 0.91 | -2.86 ± 0.29 | **4.43E-126** |
| ΔT-score [a] | -0.08 ± 0.77 | -0.88 ± 0.64 | **8.81E-92** | -0.15 ± 0.92 | -1.07 ± 0.71 | **2.89E-25** |
| Annual ΔT-score [a] | -0.02 ± 0.24 | -0.27 ± 0.22 | **3.49E-81** | -0.04 ± 0.27 | -0.31 ± 0.21 | **9.75E-24** |
| Baseline Height (cm)[a] | 159.9 ± 8.0 | 157.7 ± 8.2 | **1.06E-07** | 159.6 ± 8.0 | 157.7 ± 8.4 | **0.036** |
| Follow-up Height (cm)[a] | 159.6 ± 8.1 | 157.2 ± 8.2 | **2.82E-09** | 159.2 ± 8.0 | 157.2 ± 8.4 | **0.026** |
| Baseline Weight (kg) [a] | 59.9 ± 11.1 | 58.9 ± 11.7 | 0.086 | 58.8 ± 10.7 | 59.0 ± 10.8 | 0.869 |
| Follow-up Weight (kg) [a] | 60.4 ± 11.2 | 59.1 ± 11.7 | **0.032** | 58.7 ± 10.9 | 58.3 ± 11.1 | 0.709 |
| Baseline BMI ($kg/m^2$) [a] | 23.4 ± 3.5 | 23.6 ± 3.7 | 0.190 | 23.0 ± 3.5 | 23.6 ± 3.3 | 0.103 |
| Follow-up BMI ($kg/m^2$) [a] | 23.6 ± 3.6 | 23.8 ± 3.7 | 0.225 | 23.1 ± 3.6 | 23.5 ± 3.4 | 0.323 |
| Fracture, N (%) [c] | 90 (2.1%) | 14 (3.2%) | 0.127 | 8 (1.7%) | 3 (2.6%) | 0.540 |
| Obesity, N (%) [d] | 1,355 (31.0%) | 148 (33.5%) | 0.602 | 119 (25.6%) | 45 (38.8%) | **0.040** |
| Smoking, N (%) [e] | 1,628 (37.3%) | 138 (31.2%) | **0.009** | 167 (35.9%) | 40 (34.5%) | 0.851 |
| High alcohol intake, N (%) [f] | 596 (13.6%) | 39 (8.8%) | **0.014** | 67 (14.4%) | 17 (14.7%) | 0.762 |
| Physical inactivity, N (%) [g] | 2,645 (60.5%) | 248 (56.1%) | **0.003** | 265 (57.0%) | 49 (42.2%) | **0.019** |

ΔT-score was calculated as follow-up T-score minus baseline T-score. The annual change in T-score (annual ΔT-score) was obtained by dividing ΔT-score by the follow-up period (years). *P* values were derived from *t*-tests for continuous variables and Pearson's chi-square tests for categorical variables. Bold indicates statistical significance ($P < 0.05$).

Abbreviations: *BMI*, body mass index; *JPANEO*, Japonica array NEO; *JPAv2*, Japonica array version 2; *PGS*, polygenic score; *TMM*, Tohoku Medical Megabank.

a, Mean ± standard deviation.

b, Postmenopause women who reported that their menstruation had stopped at the baseline survey.

c, Fracture was defined based on self-reported questionnaire data and included fractures of the lumbar spine, femur, wrist, or humerus at the follow-up survey.

d, Obesity was defined as a body mass index (BMI) ≥25 $kg/m^2$ at the baseline survey.

e, Participants who had smoked more than 100 cigarettes in total were classified as smokers at the baseline survey.

f, Participants who consumed ≥30 g of alcohol daily were defined as "high alcohol intake" at the baseline survey.

g, Participants who engaged in leisure-time exercise less than three times per week were categorized as the "physical inactivity" group at the baseline survey.

**Supplementary Table S13. Interaction between age and PGS in relation to prevalent osteoporosis at baseline.**

| | Variable | OR | 95%CI lower | 95%CI upper | *P* value | Cochran's Q | $I^2$ (%) | $p_{het}$ |
|---|---|---|---|---|---|---|---|---|
| Validation cohort I (TMM9K-JPAv2, N=7,609) | Age | 1.07 | 1.07 | 1.08 | **1.19E-116** | – | – | – |
| | Sex | 2.15 | 1.86 | 2.49 | **3.13E-25** | – | – | – |
| | PGS (normalized) | 0.69 | 0.47 | 1.02 | 0.062 | – | – | – |
| | Age x PGS (normalized) | 1.00 | 1.00 | 1.01 | 0.206 | – | – | – |
| Validation cohort II (TMM18K-JPANEO, N=1,061) | Age | 1.06 | 1.04 | 1.07 | **3.55E-12** | – | – | – |
| | Sex | 0.98 | 0.72 | 1.36 | 0.921 | – | – | – |
| | PGS (normalized) | 0.71 | 0.30 | 1.74 | 0.456 | – | – | – |
| | Age x PGS (normalized) | 1.00 | 0.99 | 1.02 | 0.661 | – | – | – |
| Fixed-effect meta-analysis | Age | 1.07 | 1.07 | 1.08 | **3.89E-126** | 4.44 | 77.5 | 0.04 |
| | Sex | 1.88 | 1.65 | 2.15 | **4.493E-21** | 18.92 | 94.7 | 0.00 |
| | PGS (normalized) | 0.70 | 0.49 | 0.99 | **0.044** | 0.00 | 0.0 | 0.95 |
| | Age x PGS (normalized) | 1.00 | 1.00 | 1.01 | 0.181 | 0.01 | 0.0 | 0.93 |
| Random-effects meta-analysis | Age | 1.07 | 1.05 | 1.09 | **1.67E-13** | – | – | – |
| | Sex | 1.47 | 0.69 | 3.17 | 0.320 | – | – | – |
| | PGS (normalized) | 0.70 | 0.49 | 0.99 | **0.044** | – | – | – |
| | Age x PGS (normalized) | 1.00 | 1.00 | 1.01 | 0.181 | – | – | – |

Multivariable logistic regression models including age, sex, normalized PGS, and an age × PGS interaction term were fitted in each validation cohort to estimate ORs for prevalent osteoporosis at baseline. Cohort-specific estimates were pooled using fixed-effect and random-effects (DerSimonian–Laird) meta-analyses; fixed-effect estimates were prespecified as primary, with random-effects estimates examined as sensitivity analyses. Cochran's Q, $I^2$, and Phet summarise between-cohort heterogeneity. Bold indicates statistical significance ($P < 0.05$).

Abbreviations: *CI*, confidence interval; *$I^2$*, percentage of variation due to heterogeneity; *JPANEO*, Japonica Array NEO; *JPAv2*, Japonica Array version 2; *OR*, odds ratio; *Phet*, P value for heterogeneity; *PGS*, polygenic score; *TMM*, Tohoku Medical Megabank.

**Supplementary Table S14. Differences in the age–T-score slope by PGS-based genetic risk group.**

| | Genetic risk [quintile] | ΔSlope vs Intermediate | 95%CI | | *P* value | Cochran's Q | $I^2$ (%) | $p_{het}$ |
|---|---|---|---|---|---|---|---|---|
| | | | lower | upper | | | | |
| Validation cohort I (TMM9K-JPAv2, N=7,609) | High [Q1] | 0.003 | -0.001 | 0.008 | 0.117 | – | – | – |
| | Low [Q5] | -0.002 | -0.006 | 0.003 | 0.487 | – | – | – |
| Validation cohort II (TMM18K-JPANEO, N=1,061) | High [Q1] | 0.002 | -0.009 | 0.013 | 0.688 | – | – | – |
| | Low [Q5] | 0.004 | -0.007 | 0.015 | 0.503 | – | – | – |
| Fixed-effect meta analysis | High [Q1] | 0.003 | -0.001 | 0.007 | 0.108 | 0.033 | 0.0 | 0.856 |
| | Low [Q5] | -0.001 | -0.005 | 0.003 | 0.687 | 0.769 | 0.0 | 0.381 |
| Random-effects meta analysis | High [Q1] | 0.003 | -0.001 | 0.007 | 0.108 | – | – | – |
| | Low [Q5] | -0.001 | -0.005 | 0.003 | 0.687 | – | – | – |

Differences (Δ) in the age–T-score regression slope were estimated for the high- (Q1) and low-risk (Q5) groups relative to the intermediate-risk group (Q2–Q4, reference) in each validation cohort. Cohort-specific estimates were pooled using fixed-effect and random-effects (DerSimonian–Laird) meta-analyses; fixed-effect estimates were prespecified as primary, with random-effects estimates examined as sensitivity analyses. Cochran's Q, $I^2$, and Phet summarise between-cohort heterogeneity. Bold indicates statistical significance ($P < 0.05$).

Abbreviations: *CI*, confidence interval; *$I^2$*, percentage of variation due to heterogeneity; *JPANEO*, Japonica Array NEO; *JPAv2*, Japonica Array version 2; *Phet*, P value for heterogeneity; *PGS*, polygenic score; *Q,* quintile; *TMM*, Tohoku Medical Megabank.

**Supplementary Table S15. Differences in predicted T-score at age 20 for high- and low-genetic risk groups versus the intermediate-risk group (Q2–Q4), extrapolated from linear regression.**

| | Genetic risk [quintile] | Predicted T-score at age 20 | | Differences in predicted T-score at age 20 vs Intermediate | 95%CI | | *P* value | Cochran's Q | $I^2$ (%) | $p_{het}$ |
|---|---|---|---|---|---|---|---|---|---|---|
| | | Mean | SE | | lower | upper | | | | |
| Validation cohort I (TMM9K-JPAv2, N=7,609) | High [Q1] | -0.21 | 0.04 | -0.08 | -0.14 | -0.02 | **0.006** | – | – | – |
| | Intermediate [Q2 - Q4] | -0.13 | 0.04 | – | – | – | – | – | – | – |
| | Low [Q5] | 0.04 | 0.04 | 0.17 | 0.11 | 0.23 | **1.00E-08** | – | – | – |
| Validation cohort II (TMM18K-JPANEO, N=1,061) | High [Q1] | -0.38 | 0.10 | -0.10 | -0.25 | 0.06 | 0.214 | – | – | – |
| | Intermediate [Q2 - Q4] | -0.29 | 0.09 | – | – | – | – | – | – | – |
| | Low [Q5] | -0.20 | 0.11 | 0.09 | -0.06 | 0.24 | 0.222 | – | – | – |
| Fixed-effect meta analysis | High [Q1] | – | – | -0.08 | -0.14 | -0.03 | **0.003** | 0.03 | 0.0 | 0.86 |
| | Low [Q5] | – | – | 0.16 | 0.11 | 0.21 | **7.27E-09** | 0.84 | 0.0 | 0.36 |
| Random-effects meta analysis | High [Q1] | – | – | -0.08 | -0.14 | -0.03 | **0.003** | – | – | – |
| | Low [Q5] | – | – | 0.16 | 0.11 | 0.21 | **7.27E-09** | – | – | – |

Predicted T-scores at age 20 were obtained by extrapolating linear regression models of T-score on age within each PGS-based genetic risk group. The table shows mean predicted T-scores at age 20 and differences (Δ) for the high-risk (Q1) and low-risk (Q5) groups compared with the intermediate-risk group (Q2–Q4, reference) in each validation cohort. Cohort-specific estimates were pooled using fixed-effect and random-effects (DerSimonian–Laird) meta-analyses; fixed-effect estimates were prespecified as primary, with random-effects estimates examined as sensitivity analyses. Bold indicates statistical significance ($P < 0.05$).

Abbreviations: CI, confidence interval; $I^2$, percentage of variation due to heterogeneity; JPANEO, Japonica Array NEO; JPAv2, Japonica Array version 2; Phet, p value for heterogeneity; PGS, polygenic score; SE, standard error; TMM, Tohoku Medical Megabank.
